# Strengthening specialized capacity in Greek public services through the co-development and implementation of psychotherapeutic protocols for child and adolescent mental health conditions

**DOI:** 10.64898/2026.04.30.26352158

**Authors:** Kalliopi Triantafyllou, Anastasia Koumoula, Elianna Konialis, Panos Papoulias, Dimitra Moustaka, Vasiliki Eirini Karagiorga, Julia Luiza Schafer, Andre Simioni, Lauro Estivalete Marchionatti, Kenneth Schuster, Jill Emanuele, Caio Borba Casella, Heather Bernstein, Angela Breidenstine, Emma C. Woodward, Christina Tsoukala, Katherina Aggeli, Georgia Kaklamani, Panagiota Balikou, Christina Varveri, Eirini Karyotaki, Takis Koulouvaris, Nikolaos Scarmeas, Sarah Burke, Peter Szatmari, Pim Cuijpers, Ronald Rapee, Christos Vlachos, Sophia Parousi, Fani Vasilopoulou, Aspasia Serdari, Lilian Athanasopoulou, Nikolaos Zilikis, Vaios Dafoulis, Maria Basta, Konstantinos Kotsis, Katherina Papanikolaou, Harold Koplewicz, Giovanni A. Salum

## Abstract

**Introduction:** Despite evidence supporting child and adolescent cognitive behavioural therapy (CBT) globally, interventions remain largely unavailable within public systems. This gap requires implementation models integrating development, training, and scalability research within real-world settings and changes in management structures. Here we developed and implemented manualised psychotherapeutic protocols through a national capacity-building initiative in Greece.

**Methods:** We designed a structured implementation pathway conducted through the Child and Adolescent Mental Health Initiative (CAMHI), a public–private partnership involving clinical and academic institutions across Greece: (1) pre-implementation research through national reviews and national surveys of health professionals; (2) co-development and iterative pilot implementation of evidence-based interventions through supervised practice within the National Health System; and (3) dissemination research supporting scalability and institutionalisation within public structures.

**Results:** Pre-implementation research identified gaps in the availability of clinical protocols in Greek services; a survey of 120 psychologists and psychiatrists indicated the need for psychotherapeutic training. Three evidence-based protocols were co-developed: CBT for anxiety (6–12 years), CBT for depression (12–17 years), and behavioural parent training (BPT) for disruptive behaviour (4–14 years). During a three-stage pilot, 45 clinicians delivered interventions to 140 cases (anxiety n=52; depression n=25; BPT n=63); 117 (83·5%) completed treatment. Significant symptom reductions were observed for anxiety (d=−2·92; RCADS-25), depression (d=−1·79, RCADS-25), and disruptive behaviour (d=−2·3, SNAP-IV), with 63%, 38% and 44% showing reliable improvement at the treatment endpoint. A train-the-trainer model is under implementation for national scale-up. Institutionalisation includes integration into child and adolescent psychiatry curricula. Sustainability safeguards were established through Law 5015/2023, with the Ministry of Health assuming operations by 2027.

**Discussion:** Pilot results demonstrate the feasibility of evidence-based psychotherapeutic interventions embedded within real-world child and adolescent services in Greece. Integrated implementation approaches provide a viable pathway for developing, refining, and scaling clinical manuals within public health provision.

## INTRODUCTION

The adoption of clinical protocols is a cornerstone of high-quality mental health services, including intensive psychosocial therapies that effectively improve treatment outcomes within specialized settings.^1–4^ Despite this strong evidence base, clinical protocols remain underused, insufficiently implemented, and rarely scaled in routine practice across the majority of national health systems, rendering these treatments largely inaccessible to patients.^4–6^ These shortcomings are especially pronounced in child and adolescent services, an age group with the highest prevalence of emerging mental health conditions yet the most limited access to timely, developmentally appropriate, and specialized care.^7–10^

Several factors contribute to this gap in real-world uptake. First, many treatment manuals and protocols are not accessible: only an estimated 25% of protocols used in published clinical trials are publicly available,^11^ and a substantial proportion remain behind paywalls, under commercial licenses, or described in research publications without being disseminated to the professional community.^5^ Second, limited accessibility is compounded by the predominance of protocols developed in high-income, English-speaking contexts, which are seldom translated or culturally adapted for diverse linguistic and service environments.^12,13^ Third, apart from notable exceptions,^14^ treatment manuals and protocols are often poorly aligned with existing care pathways and service structures, limiting awareness and creating substantial barriers to uptake.^3^ Protocols tend to be developed through top-down approaches that lack meaningful engagement of frontline clinicians, service users, and system stakeholders.^15^ This contributes to manuals being perceived by professionals as impractical, overly complex, or incompatible with the capacity, resources, and workflows of real-world services.^3^

This research-to-practice gap requires scientific approaches addressing the processes, contexts, and actors involved in the uptake of evidence-based practices in public services.^4^ Apart from the Children and Young People’s Improving Access to Psychological Therapies (CYP-IAPT) Program in England,^16^ empirical literature remains otherwise scant on how short-term, evidence-based interventions can be implemented, sustained, and scaled within real-world systems. To address this gap, the 2024 Lancet Psychiatry Commission on Transforming Mental Health Implementation Research set forth an an integrated model embedding the development, evaluation, and training of protocols within local service structures.^15^ By bringing together researchers and real-world providers, this approach aims to generate protocols that are culturally appropriate, responsive to service needs, and feasible for routine practice, thereby facilitating their adoption, dissemination, and sustainability. Importantly, it also shifts the focus of empirical assessment from demonstrating efficacy under highly-controlled conditions to establishing effectiveness and widespread implementation within the complexities and constraints of real-world settings.^17^

Here, we report the co-development and implementation of child and adolescent mental health care protocols in Greece, with the aim of strengthening specialized capacity across public services in the country. This work forms part of the Child and Adolescent Mental Health Initiative (CAMHI), a nationwide capacity building programme launched in 2021 through a grant from the Stavros Niarchos Foundation (SNF). This article presents pre-implementation research informing protocol development, co-design of clinical manuals by local and international experts, and pilot implementation within the National Health System (NHS). It also outlines scale-up and sustainment plans developed with the Ministry of Health to integrate the protocols into routine national care.

## METHODS

Our conceptual design operationalizes the integrated model of implementation science in which interventions are developed and assessed directly within the real-world systems where they are intended to be scaled (see the 2024 Lancet Psychiatry Commission on Transforming Mental Health Implementation Research).^15^ In contrast to translational approaches positioning implementation as an endpoint after efficacy research, this framework advances a coordinated continuum that integrates pre-intervention, intervention, and dissemination research. It emphasises bottom-up, context-sensitive development; partnership building with health systems and communities; co-creation between researchers and frontline clinicians implementers; embedding training and supervision capacity within implementation; and systematic assessment employing hybrid designs to evaluate outcomes in naturalistic settings rather than highly controlled trials.

This program is embedded within the organisational structure of CAMHI, a public–private partnership with the Greek state operating through an international collaboration between a local implementation partner and the Child Mind Institute (CMI, a non-profit based in the USA). A national network of hubs has been established in five regional centers (Athens, Thessaloniki, Ioannina, Alexandroupoli, and Heraklion) in partnership with academic and clinical institutions (see <u>Figure 1</u>). Each CAMHI hub is staffed by multidisciplinary professionals responsible for programme implementation. These hubs serve as primary points for outreach to public health services and participant recruitment.

**Figure 1.**
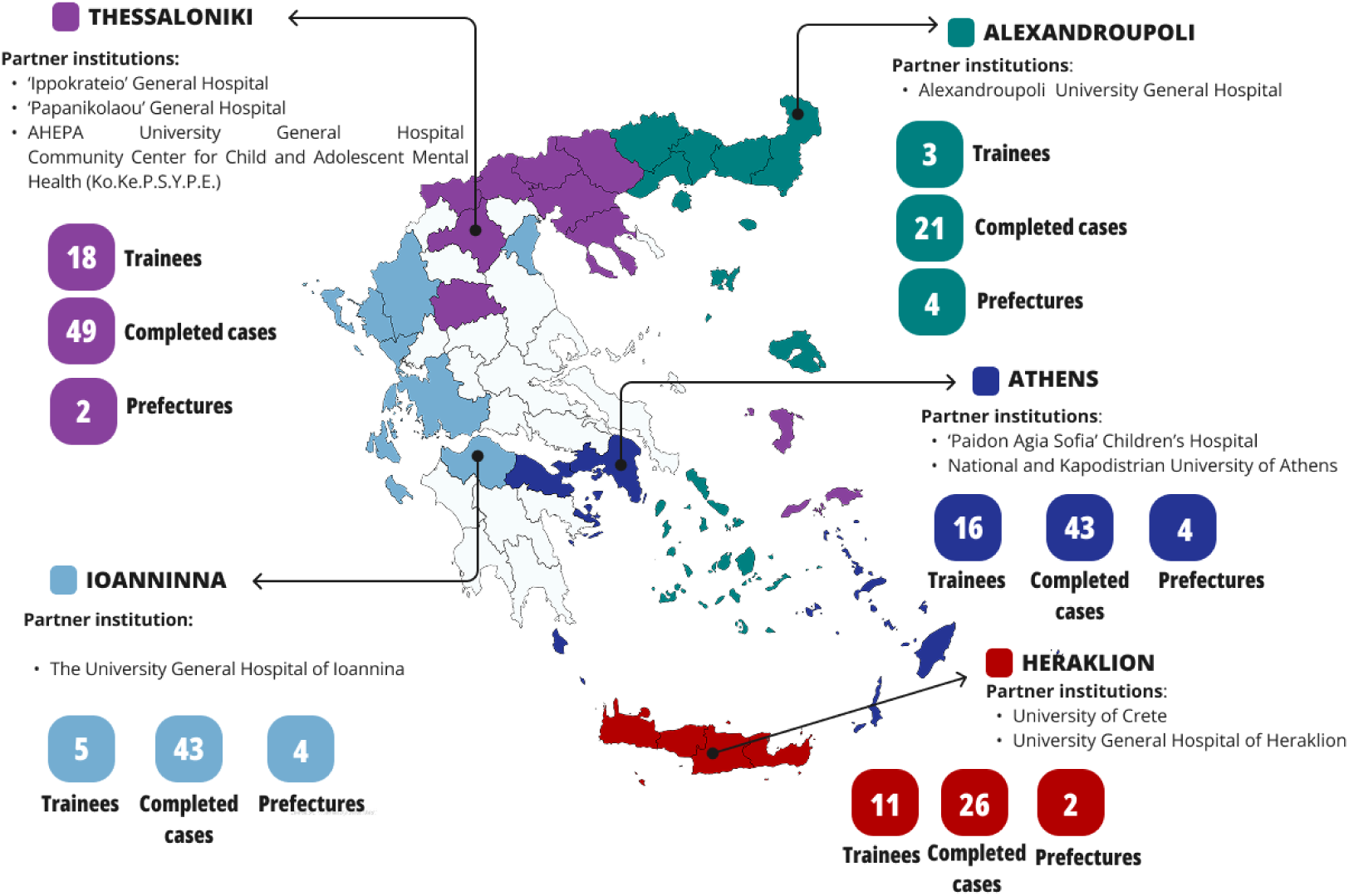
National network of implementation

### Phase 1. Pre-intervention research

#### 1.1. Examining mental health care provision and scientific resources in Greece

A comprehensive review of mental health and psychosocial care provision for children and adolescents in Greece was conducted. This review, included the organisation of the national health system, availability and affordability of care, service structure and coordination, standards of practice, workforce capacity and human resources (fully reported elsewhere).^18^ In brief, we reviewed scientific publications, governmental and institutional reports, legislative and regulatory documents, practice guidelines, and service documentation. Workforce data were obtained from official statistical sources, including professional association registries and national census data. To map service distribution and capacity, field data were collected directly from public facilities providing mental health care. In a complementary fashion, we assessed Greece’s macro-level performance using data from the Global Mental Health Countdown 2030 monitoring platform.^19^ We analyzed 39 national-level indicators related to mental health burden and service provision.

We also conducted a national systematic review of interventions related to child and adolescent mental health in Greece.^20^ Here, we draw on this resource to identify evidence-based manuals available in the country, as well as to map high-burden conditions. The review searched PubMed, Web of Science, PsycINFO, Google Scholar, and IATPOTEK from inception to December 16, 2021, and included peer-reviewed articles, book chapters, theses, and dissertations reporting data on children and adolescents up to 18 years of age in Greece. It included 34 intervention studies that employed experimental designs or reported the translation and/or adaptation of protocols for mental health promotion, prevention, or treatment.

#### 1.2. National survey with mental health professionals

We drew on our 2022-2023 national survey examining child and adolescent mental health needs in Greece according to the perspectives of 1,201 children/adolescents, 1,756 caregivers, 404 school educators, and 475 health professionals working primarily with individuals under 18 years (refer to prior work for detailed methodology).^21^ Here, we performed a subsample analysis of data from 120 mental health specialists (60 psychologists, 60 psychiatrists, and 40 child and adolescent psychiatrists). These participants were recruited through a proprietary market research panel using random sampling within each specialty and regional quotas matched to national census distributions. Data was collected online with developed questionnaires tailored to each speciality assessing: (1) main challenges encountered in clinical work; (2) professional practices, including the use of psychotherapeutic techniques, clinical approaches, and evidence-based interventions; (3) background formation; and (4) training interests.

### Phase 2. Pilot implementation

Protocols and training resources were developed by a cross-cultural team comprising local experts in Greece and international specialists from CMI, adapting clinical guidelines to the local cultural context (anxiety manual was led by EW; depression manual by HB and JE; and BPT manual by AB). Next, the program was assessed and iteratively refined with small cohorts of participants, establishing efficacy, acceptability, and feasibility prior to dissemination. This early developmental stage allowed for substantial modifications to core program components, including session structure, content and materials, supervision practices, training curricula, and delivery procedures.

Three successive pilots (I, II, and III) were conducted to enable a reassessment of incorporated changes with progressively larger samples. Trainees were mental health professionals affiliated with CAMHI hubs and partner NHS clinics, delivering the clinical protocols to one or a few participants within the routine service settings where they practice. There was an open invitation to these professionals through the director of each clinic. Supervision was provided by four CAMHI-associated experts in CBT, all qualified psychotherapy trainers, who led the training and systematically collected open feedback to support iterative refinement of materials and procedures.

Patient recruitment involved a structured assessment of candidates identified within participating clinics. Participants completed the self-reported Revised Children’s Anxiety and Depression Scale-47 (RCADS-47), alongside caregiver-reported ratings on the Swanson, Nolan, and Pelham Scale (SNAP-IV). For the anxiety protocol, children aged 6–12 years were eligible if RCADS-47 anxiety subscale T-scores were ≥65 (at occasions, we also included adolescents or children with lower scores based on professional judgment). For the depression protocol, adolescents aged 12–17 years with RCADS-47 depression T-scores ≥65 were eligible (in borderline-age cases with both anxiety and depressive symptoms above threshold, protocol allocation was determined at clinical discretion). For BPT, children aged 4–12 years were eligible if SNAP-IV scores indicated at least moderate severity on either the hyperactivity/impulsivity (≥18) or oppositional defiance (≥14) subscales.

For monitoring patient outcomes, we drew on a set of clinical instruments that were adapted to Greek language and psychometrically assessed in our national survey with 1,201 children/adolescents and 1,756 caregivers,^21–24^ This provided self- and/or caregiver-rated versions of the Pediatric Symptom Checklist (PSC), the RCADS-25, the SNAP-IV, the Parent Behavior Inventory (PBI), and the Experience of Service Questionnaire (ESQ). All caregivers, adolescents, and children aged ≥8 years completed questionnaires at baseline and at each weekly session.

Participants’ satisfaction was assessed for all participants using a brief 5-point Likert scale. We also developed questionnaires for trainees and supervisors to evaluate clinical protocols, sessions, materials, training delivery, knowledge acquisition, and development of professional competencies.

### Phase 3. Dissemination

#### 3.1. Implementation scale-up across CAMHI network (ongoing)

In this phase, training was expanded following a stepped approach to reach a larger pool of mental health professionals affiliated with CAMHI network, partner institutions, and independent NHS services. The emphasis was on replicating training procedures with fidelity while restricting modifications to the core structure of clinical protocols. We developed standardized protocols and shared operation benchmarks to ensure consistent practices across multiple implementation sites, alongside evaluating effectiveness in larger samples.

The Train-the-Trainer model was further developed during this phase, building specialized capacity to support national dissemination. CAMHI internal mental health professionals were prepared to assume trainer-supervisor roles, and eligibility criteria were defined for independent candidates applying to this preparation. Qualification requirements were designed to grant certification to new supervisors.

#### 3.2 National dissemination and sustainability (planned)

Developed in collaboration with public governance bodies, this phase focused on dissemination and institutional integration within the entire national network of public child and adolescent mental health services. CAMHI hubs provided technical and operational support to expand training to independent services and build the institutional scaffolding required to ensure sustainable program dissemination. Finally, we outlined a structured transition plan toward full uptake of operations by the Greek state. CAMHI provided intensive early support while building technical capacity to sustain future public ownership of operations.

##### Statistical analysis

This section details statistical procedures for analysing pilot implementation outcomes (Phase 2). We pooled data from Pilots I, II, and III to estimate intervention effects in a larger sample. Because the number of sessions varied across pilots, data were harmonized into common timepoints including baseline (first session), middle session, and end session. All analyses were conducted using R software (version 4.5.2).

For protocol adherence, survival curves were estimated to examine treatment dropout across sessions using the *survminer* package.^25^ Primary clinical measures were defined as RCADS-47 self-reported anxiety subscale (for anxiety treatment), RCADS-47 self-reported depression subscale (for depression treatment), and SNAP-IV caregiver-report oppositionality subscale (for disruptive behavior treatment). Secondary instruments included RCADS-47 (caregiver-report), PSC-17 (self- and caregiver-report), and the PBI. ^22,23^ The primary outcome was treatment response, defined as a ≥50% reduction in symptom severity from the beginning of treatment (raw score). Secondary outcomes included remission (defined as post-treatment scores below a T-score threshold of 55) and the Reliable Change Index (RCI).^26^ For all clinical measures, standardized T-scores were calculated based on normative references generated in national Greek samples (fully reported in dedicated validation work).^22,23^

Clinical outcomes were analysed using linear mixed-effects models to account for repeated measurements nested within individuals. Time was modelled as a fixed effect, and a random intercept was included to capture individual-level variability across sessions. This approach allows for robust estimation of change over time while accommodating unbalanced data and missing observations. Models were implemented using the *lme4* package.^27^ Estimated marginal means were derived from the fitted models, and contrasts between timepoints were computed using the *emmeans* package.^28^ Effect sizes were also derived from the mixed-effects models.

The Reliable Change Index (RCI) was estimated following the Jacobson–Truax combined method using the *clinicalsignificance* package.^26^ This approach combines two criteria: (1) whether the magnitude of change exceeds measurement error (|RCI| > 1.96), indicating reliable change; and (2) whether post-treatment scores fall within the functional population. The Reliability parameter was based on omega coefficients derived from the national Greek sample.^22,23^ Functional parameters were defined using T-score norms (mean = 50, SD = 10). The cutoff separating clinical and functional ranges was calculated using the “C” criterion, which corresponds to the weighted average of the clinical and functional means, with weights based on the opposite group’s standard deviations. This cross-weighting reduces the influence of more variable distributions and improves classification accuracy. Based on these criteria, participants were classified as recovered, improved, unchanged, deteriorated, or harmed.^26^

##### Ethical approval and data management

We obtained ethical approval from local boards across all study centers: the Scientific Council of “Agia Sofia” Children’s General Hospital (27761/04.12.2023; 20381/12.08.2024) in Attica; the Scientific Councils of Hippokrateio Hospital (52297/17.11.2023) and Papanikolaou Hospital (925/28.11.2023; 1218/10.09.2024) in Macedonia; the administrative Board of the University General Hospital of Ioannina (36/19.12.2023); the Scientific Council of the University General Hospital of Heraklion (36515/31.10.2023); the Administrative Board of the University General Hospital of Alexandroupolis (54805/09.11.2023; 55388/04.11.2024).

All families provided written informed consent and/or assent for the voluntary participation of children, adolescents, and caregivers. Trainees and supervisors also signed informed consent. Data were collected and managed using Kobo, with security measures in compliance with the General Data Protection Regulation (GDPR) and national policies established by the European Parliament and the Council of the European Union.^29^ All datasets were de-identified through the use of participant codes, and access was strictly restricted to a limited number of authorized professionals. The Thinkific platform served as a secure virtual training environment, hosting asynchronous lectures and online supervision sessions.

## RESULTS

Results are presented across three implementation stages. First, we report pre-intervention research examining mental health service capacity and professional training needs in Greece. Next, we describe the co-development and pilot implementation of three 16-session manualised treatment protocols delivered through a supervised clinician training programme. Finally, we outline the ongoing dissemination phase and national scale-up strategy across the CAMHI network. All manuals and supporting materials (including patient worksheets, therapist workbooks, and supervisor guides) remain freely available for professionals participating in advanced training.

### Phase 1: Pre-intervention research

#### 1.1. Examining mental health care provision and scientific resources in Greece

The National Health Care Service (ESY) is the health system designed to provide universal coverage in Greece through a mixed model characterized by public governance with substantial private-sector participation. The Ministry of Health centralizes governance at national level, with a regionalization framework involving seven Regional Health Authorities (YPEs) and municipal administrations. Mental health care is formally included as a core service, and recent policies have increasingly prioritized child and adolescent mental health, promoting regionalization and community-based care through outpatient services, day centres, inpatient units, and school-based psychoeducational facilities. However, implementation of these structures remains limited. Public mental health infrastructure is scarce and unevenly distributed, and coordination across services is weak, with no integrated stepped-care model or clearly defined patient pathways. These challenges push many families toward private-sector care with high out-of-pocket costs. Consequently, mental health coverage, accessibility, and affordability remain limited and unequal. Rates of minimally adequate treatment for major depressive disorder range between 20% and 29%, with particularly pronounced access gaps for children and adolescents.^19^

Specialist workforce capacity to deliver high-intensity care is adequate in aggregate (406 child and adolescent psychiatrists; about 21 per 100 000 individuals under 19 years). However, clinicians predominantly practice in the private sector, and public services are concentrated in a few urban centers. This geographic inequity is reflected in inpatient infrastructure: while the national average is 3 beds per 100,000 individuals under age 18, regional rates fluctuate from 6.79 in the 1st Health Region (Attica) to zero in the 5th Health Region (Thessaly and Central Greece). Clinical protocols are largely lacking, as existing manuals are narrow in scope and poorly disseminated.^30^ Professional practices are highly variable and often determined by individual preference or local tradition rather than evidence-based guidelines, and gaps in psychotherapeutic competencies are not uncommon. Yet, the actual standards of care remain largely unknown due to an absence of service monitoring mechanisms.

A review of scientific literature also reveals a shortage of evidence-based resources to support implementation. We could not identify an intervention ready to scale, as the few reports applying manualised CBT or other high-intensity psychotherapies were limited in scope, often delivered in group formats, and generally lacked cross-cultural validation and integration within public services.^31–33^

#### 1.2. National survey with mental health professionals (Sep 22 - Jan 23)

<u>Supplementary Table 1</u> describes the sample of 60 psychologists, 60 general psychiatrists, and 40 child and adolescent psychiatrists who participated in the survey. In routine practice, regular provision of psychotherapy was reported by 75% of psychologists and 83% of child and adolescent psychiatrists, compared with only 6% of general psychiatrists. <u>Supplementary Figure 1</u> shows the mental health conditions encountered in practice, with internalising symptoms of anxiety and depression, and behavioural difficulties being the most common across specialists.

Among participants, only 58% of psychologists reported feeling adequately qualified to manage child and adolescent mental health conditions, despite 85% having received some training in this area. Among physicians, training in psychotherapy for children and adolescents was reported by 38% of general and 71% of child and adolescent psychiatrists. <u>Supplementary Figure 2</u> summarizes background training across clinical competences. Considering psychology bachelors, 41–43% of respondents reported cognitive, psychodynamic, family-based, and behavioural therapies as absent or only superficially covered. As for residency programmes in child and adolescent psychiatry, about 27–32% reported insufficient training in these areas.

<u>Supplementary Figure 3</u> displays training interests, indicating overall receptiveness to the implementation of manualised therapies. Psychologists expressed the lowest interest: for training in disruptive behaviour disorders, 23% would definitely and 30% would maybe undertake training; for CBT targeting anxiety and depressive disorders, 23% would definitely and 20% would maybe undertake training. Child and adolescent psychiatrists showed the highest interest overall, with 62% reporting they would definitely undertake parent training (22% would maybe) and 45% reporting they would definitely undertake CBT training for anxiety and depression (22% would maybe).

### Phase 2: Pilot Implementation (Oct 23 to Nov 25)

We co-developed three high-intensity, manualized treatment protocols targeting distinct clinical populations: CBT for childhood anxiety, CBT for adolescent depression, and behavioural parent training for children with disruptive behaviour disorders (see <u>Table 1</u> for session structure and materials). The protocols were delivered through a supervised clinical training programme. First, clinicians completed a 26-hour asynchronous module covering psychotherapy foundations and CBT principles (<u>Supplementary Table 2</u>). Trainees then selected one or more protocols and delivered treatment under weekly two-hour group supervision led by certified supervisors.

**Table 1.**
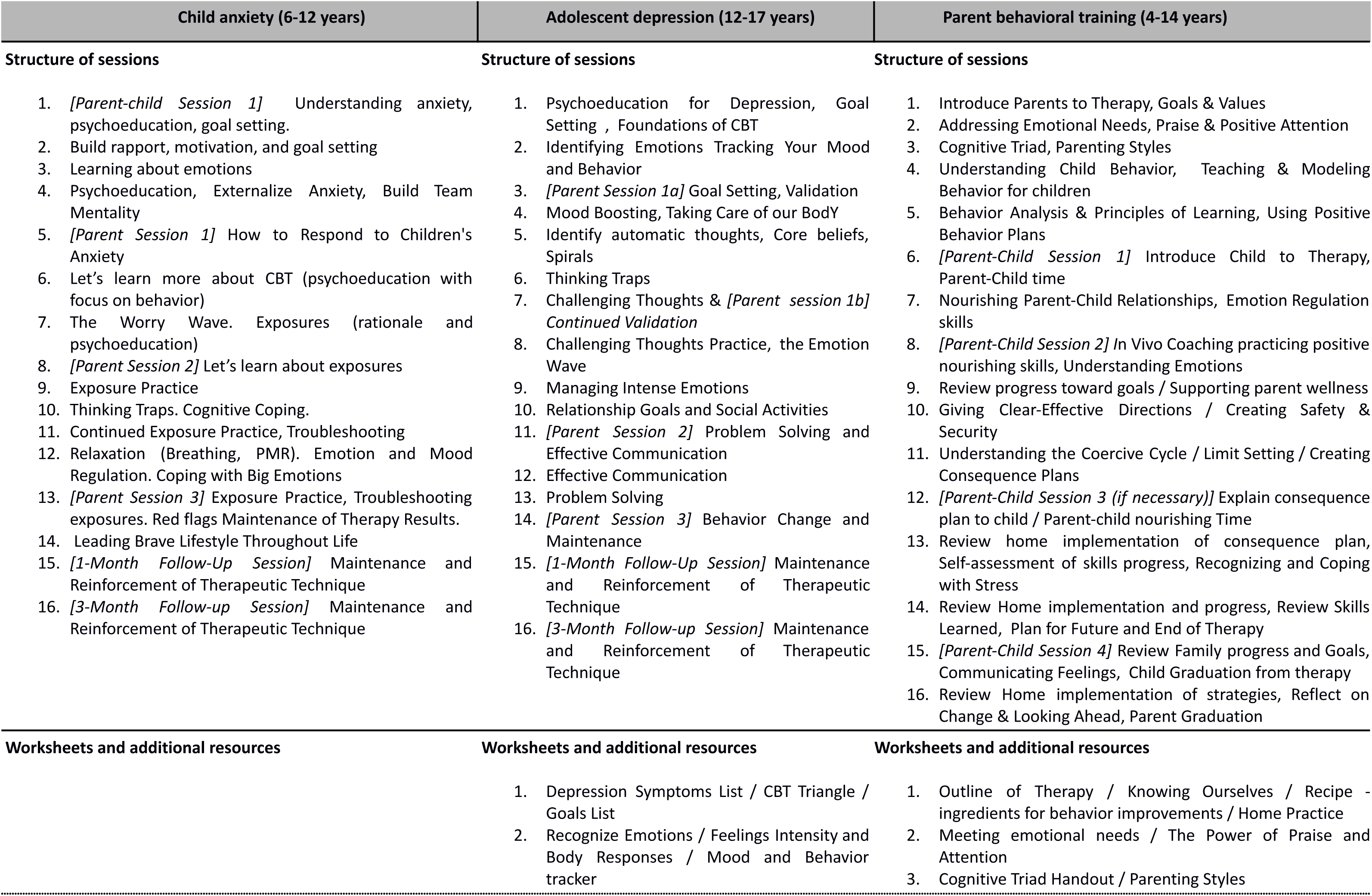
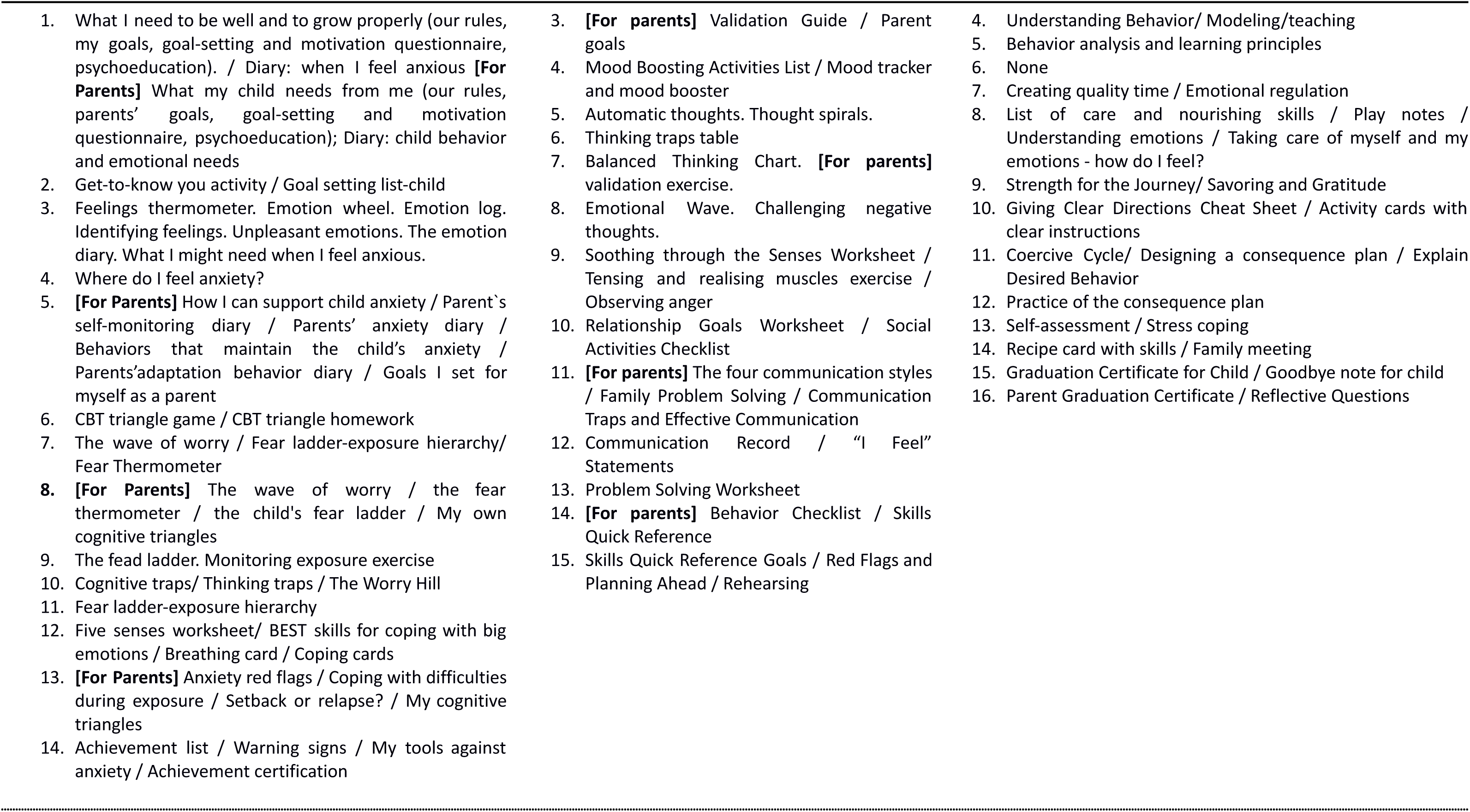
Clinical protocols: session structure and supporting worksheets.

A three-stage pilot evaluated and refined the protocols and training programme. <u>Supplementary Table 3</u> summarizes iterative refinements to the manuals. Pilot I led to the most substantial alterations in session structure and spacing, whereas Pilot II validated these data-driven refinements and resulted only in minor textual adjustments. Pilot III subsequently scaled delivery to a larger sample for ascertaining effectiveness.

<u>Table 2</u> describes participant characteristics across pilot stages. In total, supervised training delivered interventions to 140 clinical cases across the following protocols: anxiety treatment (N = 52), depression treatment (N = 25), and BPT (N = 63). Completion rates ranged from 86.5% in anxiety protocol (n = 45 completers) to 80% in depression (n = 20). <u>Figure 2</u> shows session-level dropouts across protocols, with no specific sessions associated with disproportionate attrition.

**Figure 2.**
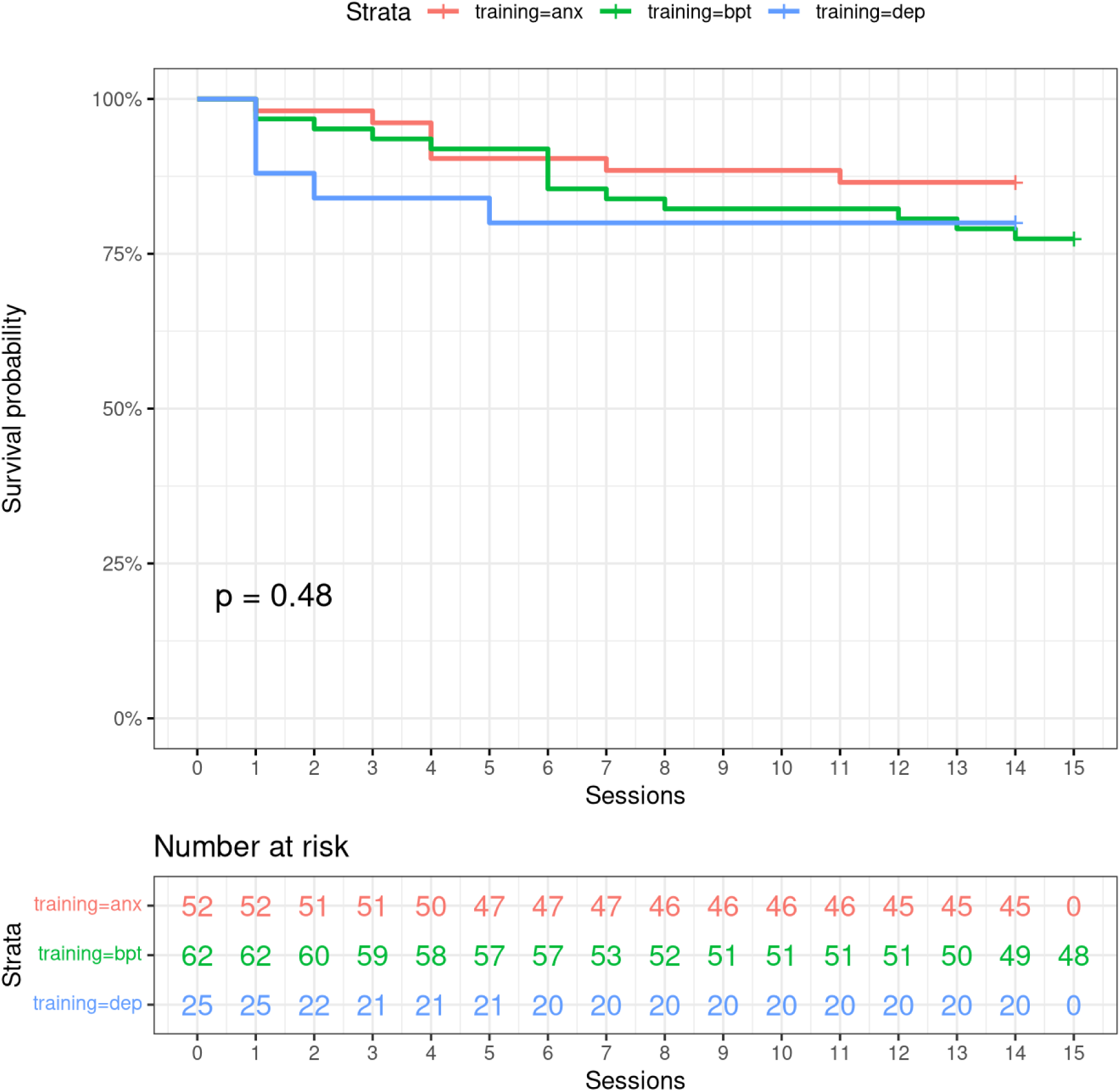
Dropout rates across protocol sessions

**Table 2.** Participants description.

|  | All protocols | Anxiety treatment<sup>1</sup> | Depression treatment<sup>1</sup> | Behavioural Parent Training<sup>2</sup> |
| --- | --- | --- | --- | --- |
| **Trainees (N)\*** | **45** | **18** | **16** | **31** |
| Pilot 1 | 16 | 5 | 4 | 7 |
| Pilot 2 | 32 | 9 | 10 | 18 |
| Pilot 3 | 28 | 12 | 9 | 17 |
| **Participants (N)** | **140** | **52** | **25** | **63** |
| Completion rates (%<br>n) | 83,5% (117) | 86,5% (45) | 80% (20) | 82,5% (52) |
| Pilot 1 (N / completed) | 22 (20) | 6 (5) | 4 (4) | 12 (11) |
| Pilot 2 | 52 (43) | 16 (14) | 10 (8) | 26 (21) |
| Pilot 3 | 66 (55) | 30 (27) | 11 (8) | 25 (20) |
| **Age** |  |  |  |  |
| Mean (SD) | 10.33 (3.72) | 10.5 (2.0) | 15.21 (1.83) | 7.61 (2.47) |
| Range | 4.00-17.55 | 5.88-13.24 | 11.44-17.55 | 4.00-14.34 |
| **Gender** |  |  |  |  |
| Male | 77 (55,3%) | 21 (40%) | 6 (25%) | 50 (81%) |
| Female | 62 (44,6%) | 31 (60%) | 19 (76%) | 12 (19%) |
| **Caregiver's age: mean (SD)** |  | 42 (10) | 50.6 (5.8) | 41 (6.1) |
| **Caregiver's gender** |  |  |  |  |
| Male | 12 (8.6%) | 5 (10%) | 4 (16%) | 3 (5%) |
| Female | 127 (91%) | 47 (90%) | 21 (84%) | 59 (95%) |
| **School grade** |  |  |  |  |
| **Pre-school (4-6y)** | **19 (13,7%)** | **0 (0%)** | **0 (0%)** | **19 (31%)** |
| **Primary school** | **82 (59%)** | **41 (79%)** | **3 (12%)** | **38 (60%)** |
| 1<sup>st</sup> grade | 14 (10%) | 6 (12%) | 0 (0%) | 8 (13%) |
| 2<sup>nd</sup> grade | 19 (14%) | 4 (7.7%) | 0 (0%) | 15 (24%) |
| 3<sup>rd</sup> grade | 6 (4.3%) | 1 (1.9%) | 0 (0%) | 5 (8.1%) |
| 4<sup>th</sup> grade | 17 (12.2%) | 10 (19%) | 0 (0%) | 7 (11%) |
| 5<sup>th</sup> grade | 9 (6.5%) | 7 (13%) | 0 (0%) | 2 (3.2%) |
| 6<sup>th</sup> grade | 17 (12.2%) | 13 (25%) | 3 (12%) | 1 (1.6%) |
| **Secondary school<sup>3</sup>** | **38 (27.3%)** | **11 (21%)** | **22 (88%)** | **5 (8%)** |
| 1<sup>st</sup> grade | 18 (12.9%) | 11 (21%) | 4 (16%) | 3 (4.8%) |
| 2<sup>nd</sup> grade | 12 (8.6%) | 0 (0%) | 10 (40%) | 2 (3.2%) |
| 3<sup>rd</sup> grade | 8 (5.8%) | 0 (0%) | 8 (33,5%) | 0 (0%) |
| **Hub Center** |  |  |  |  |
| Alexandroupoli | 14 (10%) | 3 (5.8%) | 3 (12%) | 8 (13%) |
| Athens | 32 (23%) | 9 (17%) | 9 (36%) | 14 (23%) |
| Heraklion | 22 (16%) | 7 (13%) | 3 (12%) | 12 (19%) |
| Ioannina | 35 (25%) | 20 (38%) | 6 (24%) | 9 (15%) |
| Thessaloniki | 36 (26%) | 13 (25%) | 4 (16%) | 19 (31%) |

<sup>1</sup>Data based on self-report by children or adolescents. <sup>2</sup>Data based on caregiver-report <sup>3</sup>Secondary school aggregates Gymnasium, Lyceum and EPAL.
\* Some trainees participated in more than one pilot and protocol. We report the number of unique trainees as total.

<u>Table 3</u> summarises patient-reported clinical outcomes. <u>Figure 3</u> and <u>Supplementary Figure 4</u> display symptom trajectories across protocol sessions (see estimated changes in <u>Supplementary Figure 5</u>). <u>Supplementary Table 4</u> details RCI and corresponding estimations of recovery, improvement, and worsening rates.

**Figure 3.**
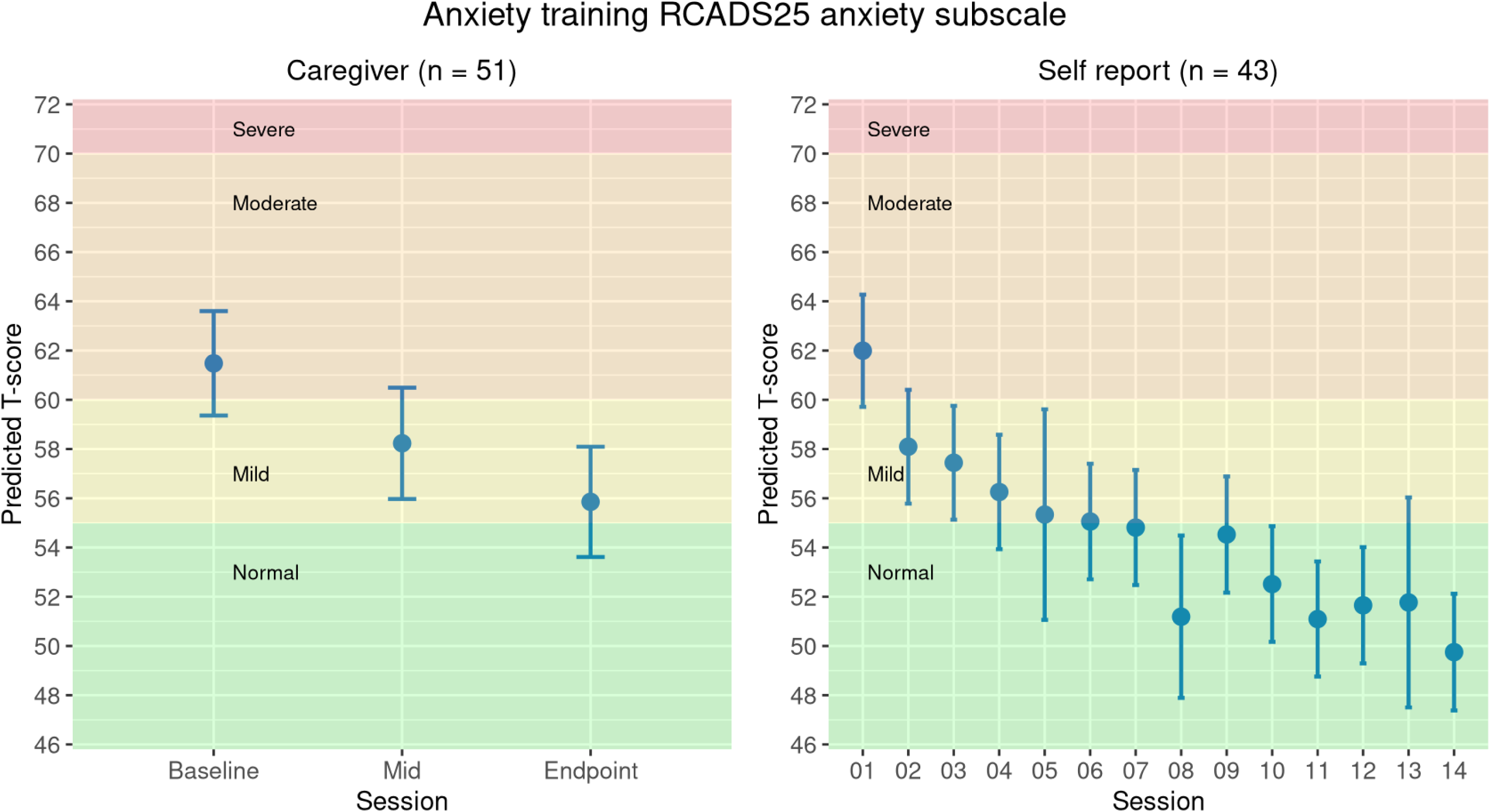
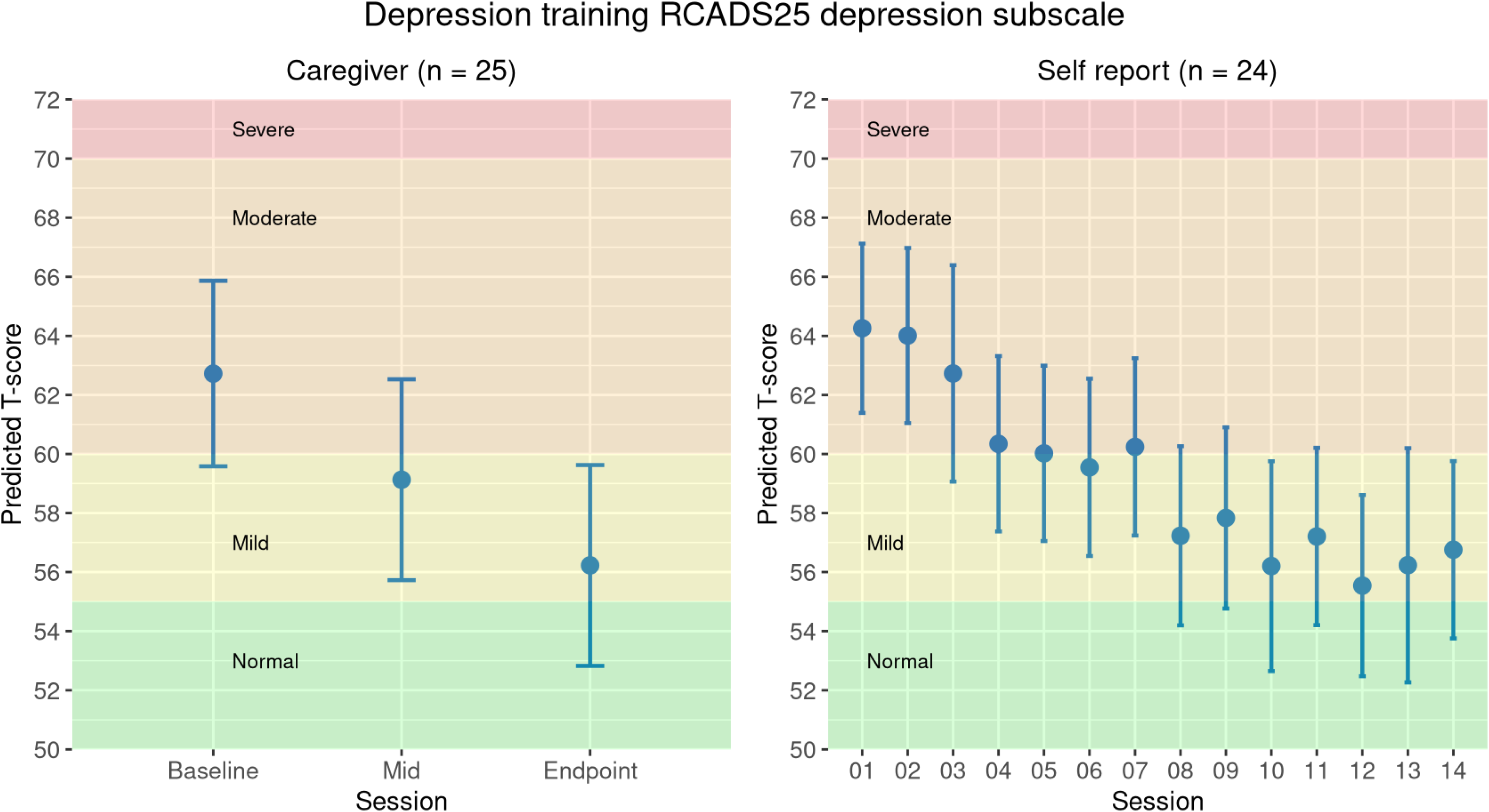
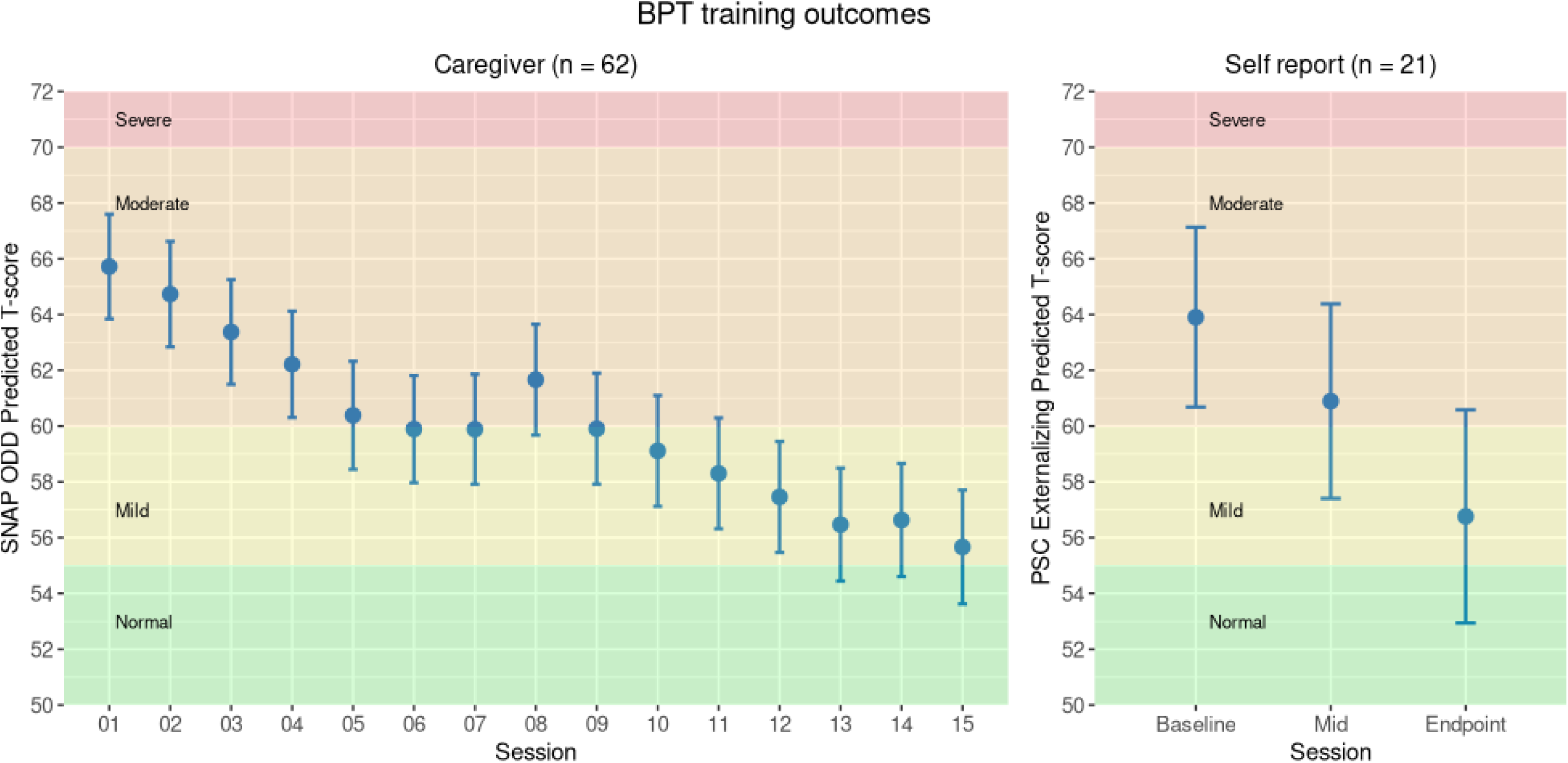
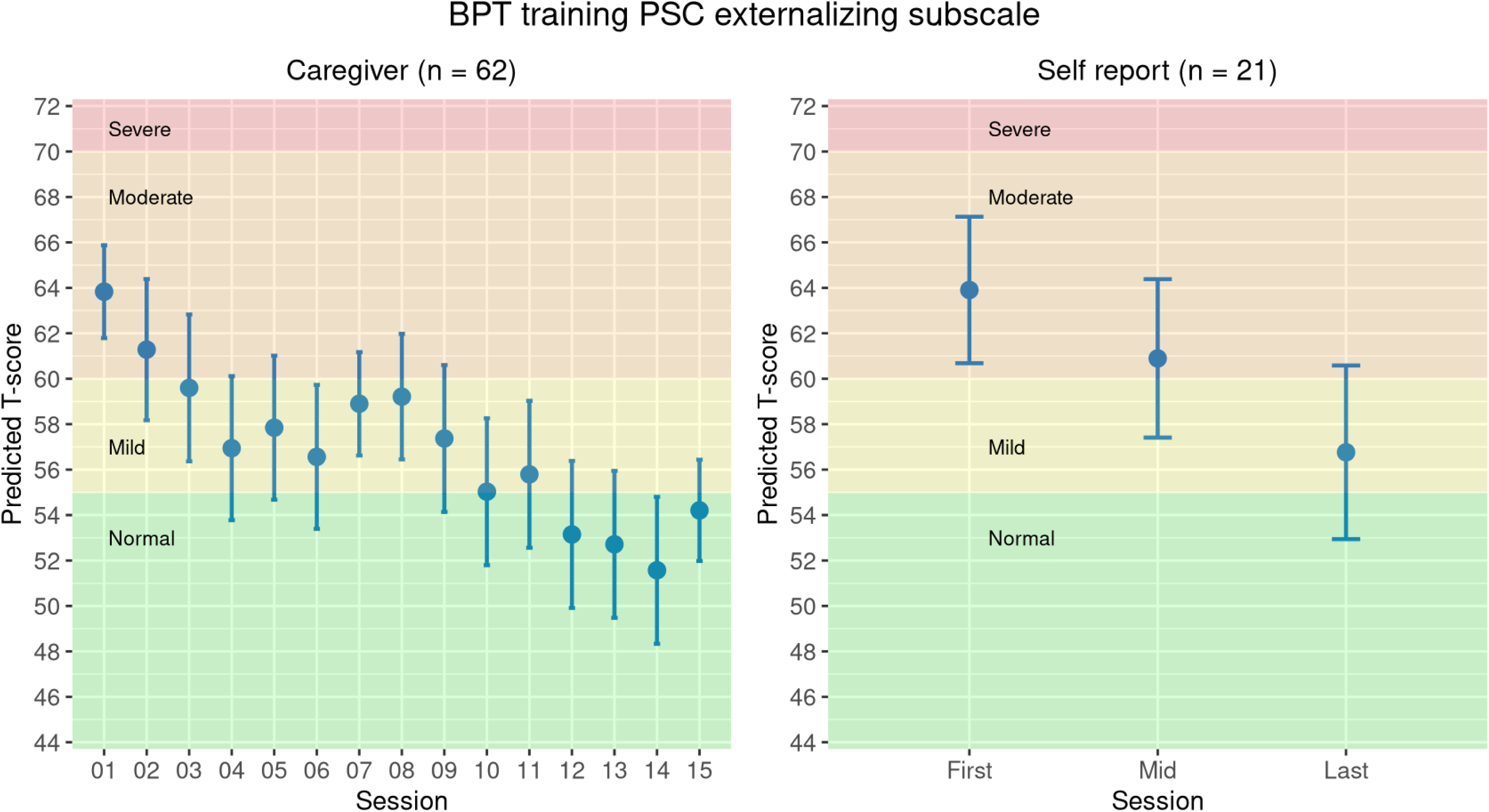
Clinical scores across protocol sessions

**Table 3.**
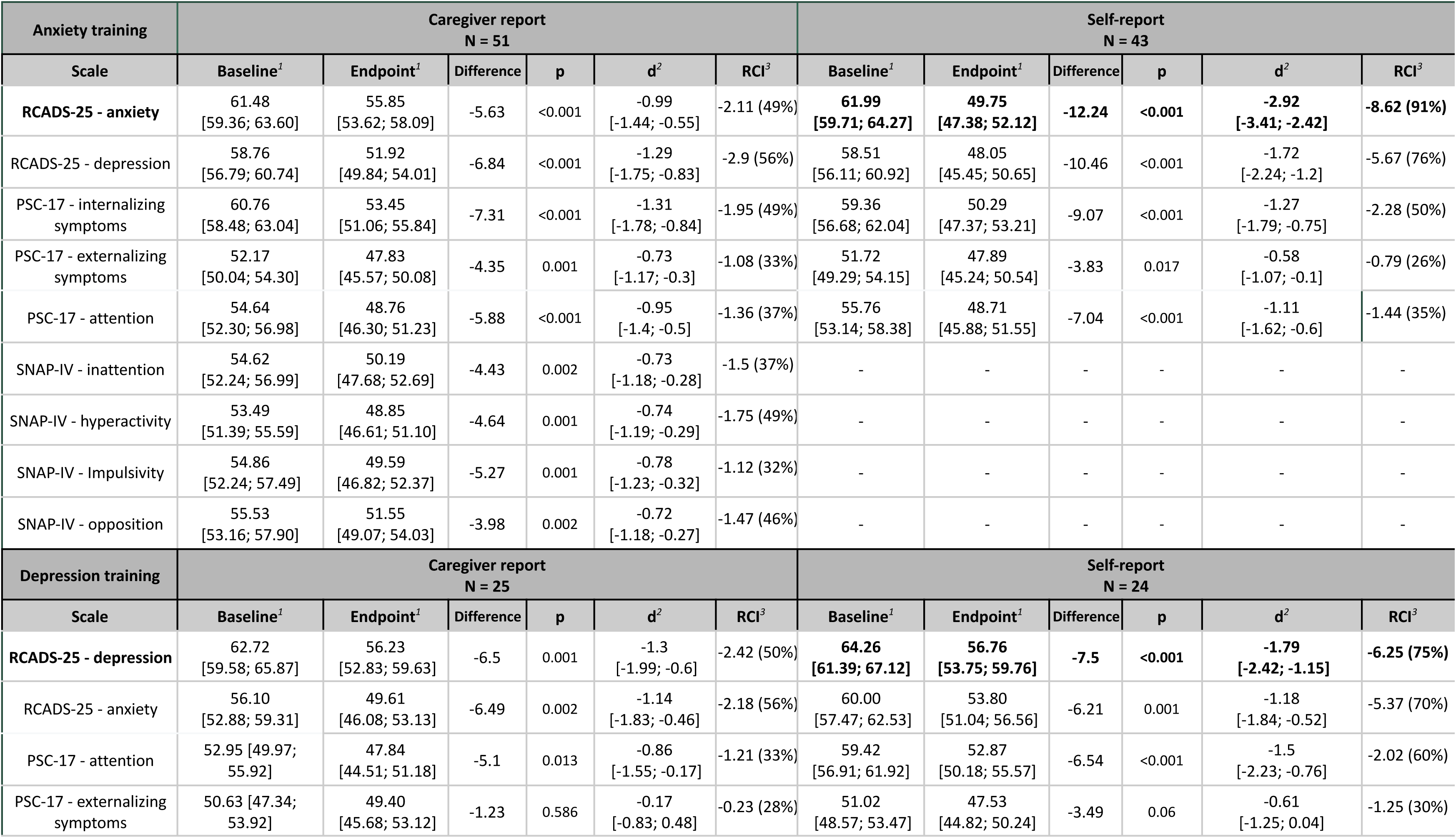
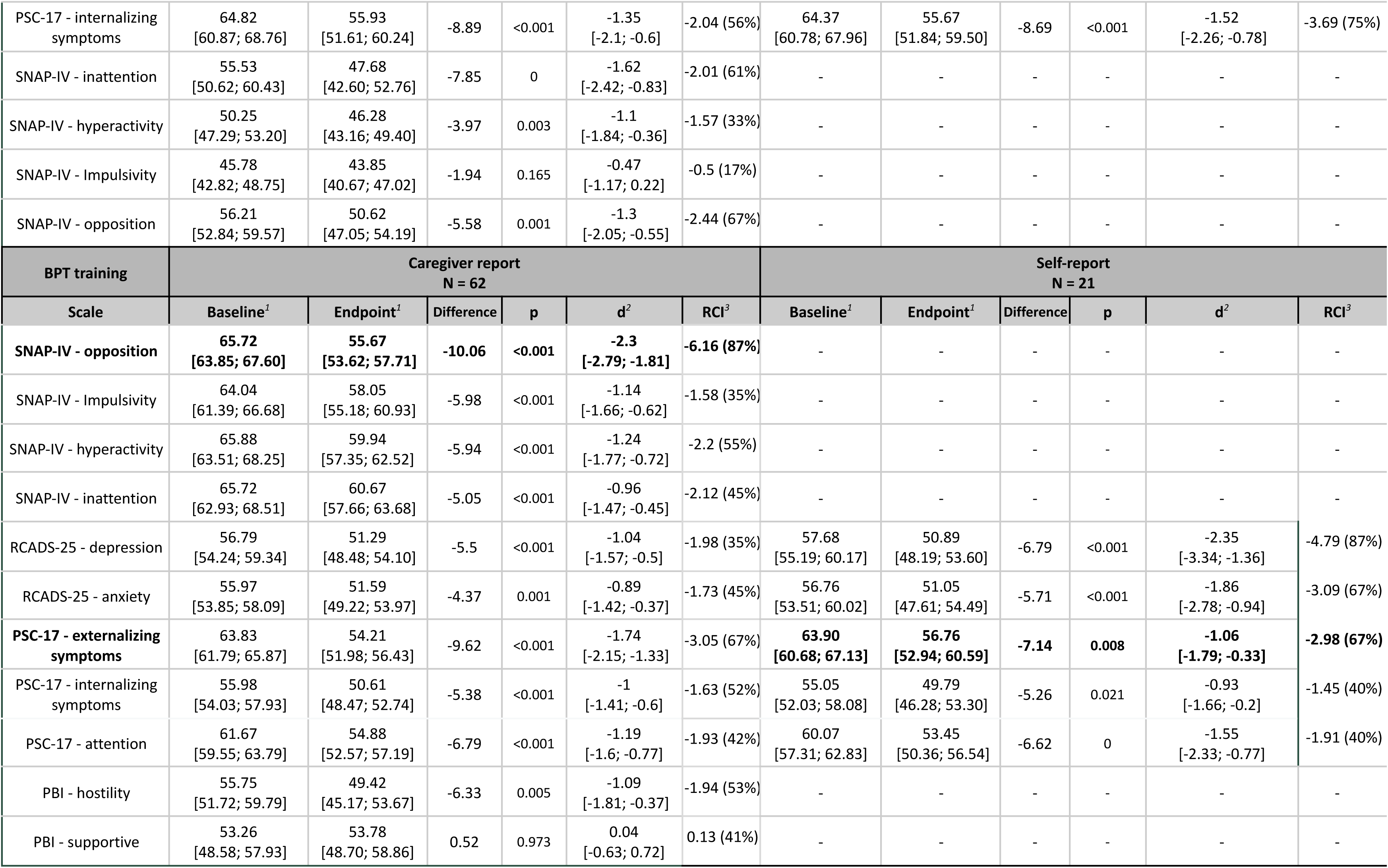
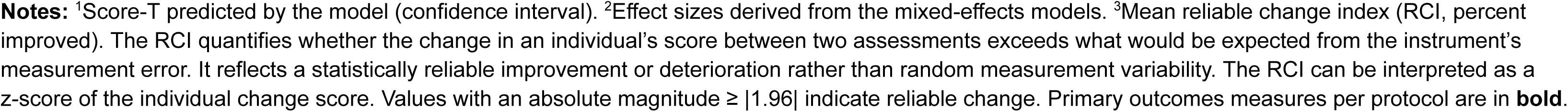
Pilot outcomes: clinical scores per treatment protocol Notes:

Overall, all protocols were associated with significant clinical improvements across primary and secondary symptoms. Large effect sizes were observed for the main outcomes in anxiety (Cohen’s d = −2.92, CI [−3.41, −2.42], p<0.001, RCADS-47 anxiety self-report subscale), depression (d = −1.79 CI [-2.42; -1.15], p<0.001, RCADS-47 self-report depression subscale), and BPT (d = −2.3 CI [-2.79; -1.81], p<0.001; SNAP-IV oppositional caregiver-report subscale). Treatment response was observed in 63% of anxiety cases (27/43), 38% of depression cases (9/24), and 44% of BPT cases (19/43), while remission occurred in 53% (23/43), 21% (5/24), and 33% (14/42), respectively.

Supplementary Table 5 details responses to the Experiences of Service Questionnaire (ESQ) items, and <u>Supplementary Figure 6</u> summarizes satisfaction rates. Overall, participants were highly satisfied across interventions according to both self- and caregiver-report. Only a small proportion of children in the BPT protocol reported being not satisfied (7%). ESQ responses further corroborated the positive appraisal of the interventions, including treatment received, facilities, training structure, logistics, and the extent to which services addressed participants’ needs.

Regarding professionals, 45 clinicians across 14 municipalities completed training, including 26 psychologists, 6 child and adolescent psychiatrists, and 13 child and adolescent psychiatry residents. There were no dropouts during protocol delivery, but 4 enrolled professionals discontinued at prior synchronous training. <u>Supplementary Figure 7</u> shows completion rates of manual items. <u>Supplementary Table 6</u> presents trainee evaluations of each protocol session in terms of materials, content, and the supervision environment; <u>Supplementary Table 7</u> shows supervisor assessment of trainees’ achievement on the therapy session’s goals. <u>Supplementary Table 8</u>, <u>Supplementary Table 9</u>, and <u>Supplementary Table 10</u> report difficulty ratings for each individual manual item across different protocol versions.

In general, sessions were positively appraised in most cases with regard to time allocation, session content and worksheets, cultural alignment, and clarity of manual instructions for applying therapeutic techniques. Supervision was also highly rated, particularly regarding the didactic guidance provided by supervisors and the collaborative learning environment. In open feedback, clinicians further emphasised the benefits of group supervision and the structured nature of the protocols. Therapists particularly valued the practical, goal-oriented strategies, which promoted active patient engagement and facilitated structured delivery of care.

### Phase 3. Dissemination

#### 3.1. Implementation scale-up across CAMHI network (Sept 25 - *ongoing*)

Beginning with the launch of the asynchronous module in September 2025, this phase is scheduled for completion in May 2026, targeting to deliver care to 65 families and qualify 30 therapists with clinical supervision initially provided by four collaborators. Professionals are trained in two protocols and are assigned five participants per protocol for a 16-week course; upon completion, trainees may extend supervised practice to three additional participants or undertake full training in a third protocol. Patient recruitment is ongoing through affiliated services across CAMHI hubs, with 74 patients enrolled and 9 drop outs to date. A total of 23 psychologists, 4 child and adolescent psychiatrists, and 3 residents in child and adolescent psychiatry have initiated training. There are 12 trainees in the anxiety programme, 9 in the depression programme, and 21 in BPT. Another 4 professionals enrolled in the program but discontinued during the synchronous training.

Official certification will be granted per protocol and is currently under refinement. Preliminary requirements for therapist certification involve completion of a 15-hour theoretical curriculum, protocol delivery to five participants (80 hours of supervised clinical practice plus 32 hours of supervision), and a satisfactory performance review. For supervisor candidates, certification will require protocol delivery to ten participants (160 hours of clinical practice plus 40 hours of supervision), mentoring with senior trainers (10 hours of observed supervision sessions), and implementation of all protocols with at least one patient each.

#### 3.2. National dissemination and sustainability

We established partnerships with the Ministry of Health, the seven Regional Health Authorities, and the National Mental Health Services Network (EDYPSY) to enable reach across public services and coordinate national rollout with a staged approach. An initial transition phase is running from February 19 to August 2026, qualifying a supervisory workforce and accreditation practices across scenarios. During this phase, 10 hub members have assumed supervisory roles under biweekly mentorship from four master trainers (2 supervisor-trainees in anxiety protocols, 3 in depression, and 5 in BPT). They provide supervision to 24 independent professionals (5 trainees in anxiety, 6 in depression, and 13 in BPT) that deliver care within their local services (outside CAMHI network). These 10 CAMHI members also deliver clinical protocols without supervision to assess feasibility and fidelity across more diverse service settings. This stage targets to deliver care to 70 cases, supporting the transition of these supervisors into train-the-trainer roles, and facilitating future accreditation of independent professionals to expand supervisory workforce. At present, the Deputy Minister is reviewing the protocols and assessing timelines for subsequent phases of dissemination.

Institutionalisation mechanisms are being defined through ongoing discussions with the Ministry of Health. The leading sustainability strategy is integration of training into national child and adolescent psychiatry residency curricula. A formal proposal to this effect has been approved by the Central Health Council (KESY), with the potential to train approximately 50 residents every five years. In addition, the program is being considered for incorporation into a Continuing Education Center of a Greek university to implement training to psychologists and other professionals.

The programme will be adopted by the Greek state to support long-term uptake, with handover of operations planned by 2028 upon completion of grant funding. A key regulatory safeguard was established under Law 5015/2023, which formalises the CAMHI public–private partnership and provides the basis for sustained implementation by the Hellenic state.

## DISCUSSION

We describe the development and early implementation of manualised treatment protocols for child anxiety, adolescent depression, and disruptive behaviour embedded within the Greek public health system. Following an integrated implementation pathway from national needs assessment and protocol co-development to pilot-to-scale delivery, the manuals were introduced and refined across real-world settings. Multi-centre pilot implementation with 45 trainee clinicians delivering treatment to 140 clinical cases showed consistent adherence to the treatment manuals, significant improvements in core symptoms, and positive appraisal by clinicians and participants. A train-the-trainer model is now being developed to support national scale-up and integrate training into medical residency programme curricula. For long-term sustainability, program operations will be fully transferred to the Greek state in 2028.

To our knowledge, this represents the first structured operationalisation of the agenda proposed by The Lancet Psychiatry Commission on transforming mental health implementation research. Whereas previous reports have mostly referred to this framework conceptually or implemented isolated elements of it,^34–40^ we outline a comprehensive pathway for embedding pre-intervention, intervention, and dissemination research within a coordinated implementation platform operating directly within routine service settings. Pre-intervention research drew on national survey data and literature reviews to assess population needs, service capacity, responsiveness to local priorities, and contextual fit, while generating evidence-based tools such as patient-reported outcome measures to support evaluation, monitoring, and accountability. Next, a national network of research–implementation teams were composed of clinicians and investigators, co-developing treatment protocols tailored to diverse regional contexts. Pilot work enabled evaluation of intervention under routine clinical conditions alongside iterative data-driven refinement of the protocols. By integrating research, workforce training and supervision, and service delivery within the same platform, the programme strengthened institutional and workforce capacity to support future scale-up. Future iterations should aim to engage a broader range of therapeutic traditions in protocol development and strengthen integration with existing training centres for psychologists and other psychotherapy professionals to enhance buy-in, dissemination, and long-term sustainability. To enhance equity, the programme should expand cultural adaptation to better reach vulnerable and underserved populations, including refugees, Roma, Pomak, and other minority groups.

We discuss some lessons that emerged from this implementation process. First, the findings underscore that sustained financing and coordinated, multi-sectoral partnerships are central to enabling large-scale mental health system reform. While short-term, fragmented funding cycles often constrain programme development, the SNF 10-year grant supported the establishment of a long-term collaborative ecosystem that integrated research, workforce training, and implementation infrastructure. This platform fostered sustained exchange between local and international experts, operating through a comprehensive national network of partners that extended beyond major urban centres, thereby underpinning the co-development of evidence-based, locally responsive protocols tailored to diverse regional contexts. A formal public–private partnership with the Greek state established the governance mechanisms necessary to integrate the programme within the public health system; specifically, formal endorsement by the Ministry of Health proved critical in addressing regulatory barriers related to clinical responsibility. It further enabled incorporation of training into official medical residency curricula, as well as enactment of legal safeguards on sustainability. Finally, we highlight that early investment in robust documentation, measurement-based care, and structured training and supervision was essential to accountability, ensuring that the interventions were not only fit for purpose but also assessable and capable of demonstrating benefit in real-world settings.

Several limitations should be acknowledged. First, the treatment protocols were not adapted from a single manual but integrated multiple evidence-based sources without systematically documenting individual contributions. Second, as an uncontrolled evaluation, improvements in clinical outcomes cannot be definitively attributed to the interventions. In this sense, we found large effect sizes that are expectedly higher than those reported in controlled trials of similar manualised CBT interventions.^41^ Third, early pilot phases involved smaller samples, and variation in session numbers across stages required harmonisation of data for outcome analyses. Finally, the initial implementers consisted of volunteers who may have been particularly motivated, whilst supervision was provided by highly-qualified professionals, potentially limiting generalisability to the broader workforce during large-scale implementation. In the ongoing dissemination phase, multicentre evaluation will be essential to assess effectiveness, scalability, and system-level impact under routine service conditions nationwide.

This initiative represents a landmark national application of an integrated implementation science model, offering a scalable blueprint for strengthening specialized workforce capacity and bridging the research-to-practice gap. By embedding research, training, and service delivery within a continuous implementation pathway, the programme co-developed, tested, and scaled interventions within routine public system settings in Greece. This approach produced evidence-based treatment protocols that were clinically effective, culturally responsive, aligned with existing health system structures, and feasible for national scale-up and long-term sustainability. Future efforts should build on this model to strengthen capacity across diverse service settings, refine implementation practices, and expand access to evidence-based mental health care.

## Data Availability

All data produced in the present study are available upon reasonable request to the authors

