## Supplementary for "Strengthening specialized capacity in Greek public services through the co-development and implementation of psychotherapeutic protocols for child and adolescent mental health conditions"

Supplementary Table 1 - National survey assessing professional needs: sample description 2
Supplementary Table 2 - Overview of theoretical curriculum 4
Supplementary Table 3 - Refinement of intervention protocols across pilot implementation 5
Supplementary Table 4 - Reliable Change Index (RCI): percentage of recovery, improvement, and worsening rates 7
Supplementary Table 5 - Experience of Service Use Questionnaire 9
Supplementary Table 6 - Protocol assessment by trainees 11
Supplementary Table 7 - Supervisor assessment: trainees`achievement of the therapy session's goals 13
Supplementary Table 8: Anxiety treatment protocol: session-level item completion and difficulty (trainee) 15
Supplementary Table 9: Depression treatment protocol: session-level item completion and difficulty (trainee) 24
Supplementary Table 10: BPT treatment protocol: session-level item completion and difficulty (trainee) 31
Supplementary Figure 1 - National survey with professionals: clinical challenges 41
Supplementary Figure 2 - National survey with professionals: background training 42
Supplementary Figure 3 - National survey assessing needs with professionals: training interests 43
Supplementary Figure 4. Clinical scores across protocol sessions 45
Supplementary Figure 5. Estimated percentage change on clinical scores across protocol sessions 49
Supplementary Figure 6 - Pilot implementation: satisfaction rates 50
Supplementary Figure 7 - Completion rate of manual items 53

Supplementary Table 1 - National survey assessing professional needs: sample description

| | Psychologists<br>N = 60 | Psychiatrists<br>N = 60 | Child psychiatrists<br>N = 40 |
|---|---|---|---|
| Age | | | |
| Mean (SD) | 47 (9) | 51 (9) | 55 (8) |
| Min, Max | 30, 67 | 37, 80 | 37, 72 |
| Gender | | | |
| Female | 46 (77%) | 16 (27%) | 26 (65%) |
| Male | 14 (23%) | 44 (73%) | 14 (35%) |
| Residence: Health Region (HR) | | | |
| 1st Health Region of Attica | 40 (67%) | 43 (72%) | 28 (70%) |
| 3rd Health Region of Macedonia | 14 (23%) | 12 (20%) | 8 (20%) |
| 4th Health Region of Macedonia and Thrace | 1 (1.7%) | 1 (1.7%) | 1 (2.5%) |
| 6th Health Region of Peloponnese, Ionian islands, Epirus and Western Greece | 2 (3.3%) | 2 (3.3%) | 1 (2.5%) |
| 7th Health Region of Crete | 3 (5.0%) | 2 (3.3%) | 2 (5.0%) |
| What is the funding provision of the service(s) you work at? | | | |
| Public | 26 (43%) | 25 (42%) | 18 (45%) |
| Private | 34 (57%) | 38 (63%) | 23 (58%) |
| Non-governmental organization (NGOs) | 2 (3.3%) | 0 (0%) | 0 (0%) |
| What type of service(s) do you work at? | | | |
| Primary care unit (rural clinic, PEDY, Medical Center etc) | 2 (3.3%) | 5 (8.3%) | 2 (5.0%) |
| Community center (Mental Health Center, Medicopedagogical center- Community Center for Mental Health of Children and Adolescents, Branches of the "Hellenic Center for Mental Health and Research-EKEPSYE", private community mental health center etc) | 5 (8.3%) | 5 (8.3%) | 6 (15%) |
| Support mobile team (offering mental health services) | 3 (5.0%) | 1 (1.7%) | 0 (0%) |
| Hospital not linked to the university – Outpatient | 4 (6.7%) | 7 (12%) | 2 (5.0%) |

| | | | |
|---|---|---|---|
| Hospital not linked to the university – Inpatient | 8 (13%) | 9 (15%) | 2 (5.0%) |
| University hospital – Outpatient | 4 (6.7%) | 3 (5.0%) | 6 (15%) |
| University hospital – Inpatient | 1 (1.7%) | 5 (8.3%) | 5 (13%) |
| Private practice | 32 (53%) | 38 (63%) | 21 (53%) |
| NGO | 4 (6.7%) | 0 (0%) | 0 (0%) |
| Do you provide any type of psychotherapy in your clinical practice? | | | |
| No | 11 (18%) | 40 (67%) | 6 (15%) |
| Yes, eventually | 4 (6.7%) | 16 (27%) | 1 (2.5%) |
| Yes, frequently | 13 (22%) | 3 (5.0%) | 18 (45%) |
| Yes, on a daily basis | 32 (53%) | 1 (1.7%) | 15 (38%) |
| Highest level of education | | | |
| University | 5 (8.3%) | 41 (68%) | 20 (50%) |
| Master's degree | 44 (73%) | 11 (18%) | 13 (33%) |
| Doctorate | 11 (18%) | 8 (13%) | 7 (18%) |

#### Supplementary Table 2 - Overview of theoretical curriculum

**Introductory online lectures: fundamentals of psychotherapy**
Before clinical practice, all trainees complete asynchronous training modules

**Section A. Psychotherapy Principles**

1. Principles, patient-centered care, and intervention impact assessment
2. Basics skills of effective therapists: establishing trust, maintaining boundaries, and following confidentiality rules
3. Working with children and adolescents, building a therapeutic relationship, and managing challenging behaviors
4. Working with parents, including them in therapy and providing feedback
5. Family system theory and conducting family therapy sessions
6. Child protection, identification of abuse and neglect, and reporting under circumstances.
7. Assessing suicidality, including effective interviewing strategies, evaluating risk levels, and protective factors
8. Trauma signs recognition, diagnostic criteria, and treatment options

**Section B. Cognitive Behaviour Therapy Principle**

1. Fundamentals of CBT, the cognitive triangle and cognitive distortion
2. Behavioral frameworks in psychotherapy, reinforcement and punishment concepts
3. Initial assessment needs for therapy selection and CBT case formulation
4. Measurement-based care and clinical case examples.
5. Engaging patients to treatment using basic motivational interviewing principles and dialectic behavioral therapy commitment strategies
6. Manage willful or challenging behaviors in treatment with parents or patients
7. Mindfulness definitions, reasons to practice it, and ways it benefits treatment, with the use of clinical examples

**Applied concepts for each clinical protocol**
Additional theoretical themes are delivered during supervision sessions

**CHILD ANXIETY**

1. Epidemiology & phenomenology of anxiety
2. Evidence of effective interventions for child anxiety
3. Anxiety CBT basic principles
4. Anxiety CBT specific techniques and active components (eg. exposures)
5. Putting theory into practice: how to use this manual

**ADOLESCENT DEPRESSION**

1. Epidemiology & phenomenology of Depression
2. Evidence of effective interventions for Adolescent depression
3. Depression CBT basic principles
4. Depression CBT specific techniques and active components eg. mood boosters
5. Putting theory into practice: how to use this manual

**PARENT BEHAVIOUR**

1. Epidemiology & phenomenology of behavioral problems
2. Evidence of effective interventions for BPT
3. BPT basic principles
4. BPT specific techniques and active components (e.g., Managing a Behavioral Chart)
5. Putting theory into practice: how to use this manual

Supplementary Table 3 - Refinement of intervention protocols across pilot implementation

| | Anxiety (6–12y) | Depression (12–17y) | BPT (4–14y) |
|---|---|---|---|
| **Pilot I (Oct 23 – Jul 24)** | **Format**: 17 sessions (5 with parents)<br>**Duration**: 17 weeks<br>**Participants**: 5 completed cases + 1 drop out (5 trainees) | **Format**: 16 sessions (3 with parents)<br>**Duration**: 16 weeks<br>**Participants**: 4 completed cases (4 trainees) | **Format**: 16 sessions (4 parent-child sessions)<br>**Duration**: 16 weeks<br>**Participants**: 11 completed cases + 1 drop out (7 trainees) |
| Refinements | • Reduced long sessions<br>• Removal of repetitive session on exposure<br>• First session: child with the parents instead of two first sessions with the parents<br>• Moved parent sessions to initial weeks<br>• Parent sessions in the same week as regular sessions<br>• More emphasis on parental contribution (not only via adjustment, but also via unrelenting standards and modeling)<br>• Additional content on core emotional needs and anxiety (1st session)<br>• Adjusted workbooks for slightly younger children<br>• More examples and experiential ideas | • Removal of mindfulness and self-compassion sessions<br>• Moved parent sessions to initial weeks<br>• Parent sessions in the same week as regular sessions<br>• Workbook with clearer examples and minor corrections | • Removal of repetitive session on consequence plan<br>• Quality time session earlier in the treatment<br>• Adapted workbook to age, severity, and comorbidities |
| **Pilot II (Sep 24 – Mar 25)** | **Format**: 14 sessions (3 with parents, 1 parent-child)<br>**Duration**: 11 weeks<br>**Participants**: 14 completed cases + 2 drop outs ( 9 trainees) | **Format**: 14 sessions (3 with parents)<br>**Duration**: 11 weeks<br>**Participants**: 8 completed cases + 2 drop outs (10 trainees) | **Format**: 15 sessions (4 parent - child)<br>**Duration**: 15 weeks<br>**Participants**: 21 completed cases + 5 drop outs (18 trainees) |
| Refinements | • Minor textual adjustments<br>• Session structure and spacing considered adequate | • Minor textual adjustments<br>• Session structure and spacing considered adequate | • Minor textual adjustments<br>• Session structure and spacing considered adequate |
| **Pilot III (Feb 25 – Nov 25)** | **Format**: 14 sessions (3 with parents, 1 parent-child)<br>**Duration**: 11 weeks<br>**Participants**: 27 completed cases + 3 drop outs (12 trainees) | **Format**: 14 sessions (3 with parents)<br>**Duration**: 11 weeks<br>**Participants**: 8 completed cases + 3 drop outs (9 trainees) | **Format**: 15 sessions (4 parent - child)<br>**Duration**: 15 weeks<br>**Participants**: 20 completed cases + 5 drop outs (17 trainees) |

| | | | |
|---|---|---|---|
| Refinements | • More emphasis and exercises on parent’s self observation<br>• Details added considering the therapeutic relationship and the ways of communicating with the child<br>•More space for cognitive work (part of the 11th session)<br>• Minor changes in the workbook content<br>• Addition of 1-month and 3-month follow-up sessions | • Added an extra adolescent session with a brief 30-min session with parents session immediately after<br>• Minor adjustments to workbooks<br>• Addition of 1-month and 3-month follow-up sessions | • Revisions of content and order of protocols adding another session:<br>● Addition of Session 2 on emotional needs<br>● Reorganization of content (e.g., placing parents’ values and goals in Session 1)<br>● Modification of examples for cultural relevance<br>● Addition of functional behavior analysis<br>● Addition of examples for the cognitive triad and positive behavior plans |
| **Full implementation** | **Format**: 16 sessions (3 with parents, 1 parent-child)<br>**Duration**: 11 weeks + *follow-up at 1 and 3 months*<br>**Participants**: 21 cases + 4 drop outs (12 trainees) | **Format**: 16 sessions (3 with parents)<br>**Duration**: 11 weeks + *follow-up at 1 and 3 months*<br>**Participants**: 13 cases + 1 drop outs (9 trainees) | **Format**: 16 sessions (4 parent-child)<br>**Duration**: 15 weeks<br>**Participants**: 31 cases + 4 drop outs (21 trainees) |

Supplementary Table 4 - Reliable Change Index (RCI): percentage of recovery, improvement, and worsening rates

| **Anxiety training** | **Caregiver report N = 51** | | | | | | **Self-report N = 43** | | | | | |
|---|---|---|---|---|---|---|---|---|---|---|---|---|
| **Scale** | **n[1]** | **Recovered[2]** | **Improved[2]** | **Unchanged[2]** | **Deteriorate[2]** | **Harmed[2]** | **n[1]** | **Recovered[2]** | **Improved[2]** | **Unchanged[2]** | **Deteriorated[2]** | **Harmed[2]** |
| RCADS-25 - depression | 41 | 11 (27%) | 12 (29%) | 15 (37%) | 2 (5%) | 1 (2%) | 34 | 17 (50%) | 9 (26%) | 8 (24%) | 0 (0%) | 0 (0%) |
| **RCADS-25 - anxiety** | 41 | 11 (27%) | 9 (22%) | 18 (44%) | 3 (7%) | 0 (0%) | **35** | **25 (71%)** | **7 (20%)** | **3 (9%)** | **0 (0%)** | **0 (0%)** |
| SNAP-IV - inattention | 41 | 9 (22%) | 6 (15%) | 21 (51%) | 2 (5%) | 3 (7%) | - | - | - | - | - | - |
| SNAP-IV - hyperactivity | 41 | 13 (32%) | 7 (17%) | 17 (41%) | 1 (2%) | 3 (7%) | - | - | - | - | - | - |
| SNAP-IV - Impulsivity | 41 | 8 (20%) | 5 (12%) | 23 (56%) | 3 (7%) | 2 (5%) | - | - | - | - | - | - |
| SNAP-IV - opposition | 41 | 10 (24%) | 9 (22%) | 16 (39%) | 5 (12%) | 1 (2%) | - | - | - | - | - | - |
| PSC-17 - attention | 43 | 9 (21%) | 7 (16%) | 26 (60%) | 0 (0%) | 1 (2%) | 34 | 10 (29%) | 2 (6%) | 22 (65%) | 0 (0%) | 0 (0%) |
| PSC-17 - internalizing symptoms | 43 | 12 (28%) | 9 (21%) | 22 (51%) | 0 (0%) | 0 (0%) | 34 | 12 (35%) | 5 (15%) | 14 (41%) | 1 (3%) | 2 (6%) |
| PSC-17 - externalizing symptoms | 43 | 9 (21%) | 5 (12%) | 25 (58%) | 1 (2%) | 3 (7%) | 34 | 6 (18%) | 3 (9%) | 22 (65%) | 0 (0%) | 3 (9%) |
| **Depression training** | **Caregiver report N = 25** | | | | | | **Self-report N = 24** | | | | | |
| **Scale** | **n[1]** | **Recovered** | **Improved** | **Unchanged** | **Deteriorated** | **Harmed** | **n[1]** | **Recovered** | **Improved** | **Unchanged** | **Deteriorated** | **Harmed** |
| **RCADS-25 - depression** | 18 | 7 (39%) | 2 (11%) | 8 (44%) | 1 (6%) | 0 (0%) | **20** | **11 (55%)** | **4 (20%)** | **4 (20%)** | **0 (0%)** | **1 (5%)** |
| RCADS-25 - anxiety | 18 | 5 (28%) | 5 (28%) | 7 (39%) | 1 (6%) | 0 (0%) | 20 | 10 (50%) | 4 (20%) | 3 (15%) | 2 (10%) | 1 (5%) |
| SNAP-IV - inattention | 18 | 3 (17%) | 8 (44%) | 7 (39%) | 0 (0%) | 0 (0%) | - | - | - | - | - | - |
| SNAP-IV - hyperactivity | 18 | 3 (17%) | 3 (17%) | 12 (67%) | 0 (0%) | 0 (0%) | - | - | - | - | - | - |
| SNAP-IV - Impulsivity | 18 | 2 (11%) | 1 (6%) | 15 (83%) | 0 (0%) | 0 (0%) | - | - | - | - | - | - |
| SNAP-IV - opposition | 18 | 4 (22%) | 8 (44%) | 5 (28%) | 1 (6%) | 0 (0%) | - | - | - | - | - | - |
| PSC-17 - attention | 18 | 4 (22%) | 2 (11%) | 12 (67%) | 0 (0%) | 0 (0%) | 20 | 8 (40%) | 4 (20%) | 8 (40%) | 0 (0%) | 0 (0%) |
| PSC-17 - internalizing symptoms | 18 | 5 (28%) | 5 (28%) | 7 (39%) | 1 (6%) | 0 (0%) | 20 | 11 (55%) | 4 (20%) | 4 (20%) | 0 (0%) | 1 (5%) |
| PSC-17 - externalizing symptoms | 18 | 4 (22%) | 1 (6%) | 10 (56%) | 0 (0%) | 3 (17%) | 20 | 5 (25%) | 1 (5%) | 12 (60%) | 0 (0%) | 2 (10%) |
| **BPT training** | **Caregiver report N = 62** | | | | | | **Self-report N = 21** | | | | | |

| Scale | n[1] | Recovered | Improved | Unchanged | Deteriorated | Harmed | n[1] | Recovered | Improved | Unchanged | Deteriorated | Harmed |
|---|---|---|---|---|---|---|---|---|---|---|---|---|
| RCADS-25 - depression | 31 | 7 (23%) | 4 (13%) | 20 (65%) | 0 (0%) | 0 (0%) | 15 | 6 (40%) | 7 (47%) | 2 (13%) | 0 (0%) | 0 (0%) |
| RCADS-25 - anxiety | 31 | 5 (16%) | 9 (29%) | 14 (45%) | 0 (0%) | 3 (10%) | 15 | 6 (40%) | 4 (27%) | 4 (27%) | 1 (7%) | 0 (0%) |
| SNAP-IV - inattention | 31 | 9 (29%) | 5 (16%) | 15 (48%) | 2 (6%) | 0 (0%) | - | - | - | - | - | - |
| SNAP-IV - hyperactivity | 31 | 11 (35%) | 6 (19%) | 12 (39%) | 2 (6%) | 0 (0%) | - | - | - | - | - | - |
| SNAP-IV - Impulsivity | 31 | 6 (19%) | 5 (16%) | 19 (61%) | 0 (0%) | 1 (3%) | - | - | - | - | - | - |
| **SNAP-IV - opposition** | **31** | **19 (61%)** | **8 (26%)** | **3 (10%)** | **1 (3%)** | **0 (0%)** | - | - | - | - | - | - |
| PSC-17 - attention | 48 | 14 (29%) | 6 (12%) | 26 (54%) | 1 (2%) | 1 (2%) | 15 | 3 (20%) | 3 (20%) | 9 (60%) | 0 (0%) | 0 (0%) |
| PSC-17 - internalizing symptoms | 48 | 12 (25%) | 13 (27%) | 20 (42%) | 1 (2%) | 2 (4%) | 15 | 5 (33%) | 1 (7%) | 8 (53%) | 1 (7%) | 0 (0%) |
| PSC-17 - externalizing symptoms | 48 | 21 (44%) | 11 (23%) | 13 (27%) | 1 (2%) | 2 (4%) | **15** | **9 (60%)** | **1 (7%)** | **5 (33%)** | **0 (0%)** | **0 (0%)** |
| PBI - hostility | 17 | 3 (18%) | 6 (35%) | 7 (41%) | 1 (6%) | 0 (0%) | - | - | - | - | - | - |
| PBI - supportive | 17 | 3 (18%) | 4 (24%) | 5 (29%) | 3 (18%) | 2 (12%) | - | - | - | - | - | - |

**Notes:** Clinical significance was evaluated using the Jacobson–Truax combined method (cutoff C). Reliable change was determined with the Reliable Change Index (RCI), accounting for measurement error based on instrument reliability. The clinical cutoff separating clinical and functional populations was calculated from the means and standard deviations of both groups, and participants were classified according to reliable change and whether their post-treatment score crossed this cutoff.

[1]Participants with data in baseline and endpoint. [2]**Recovered**, the individual showed a reliable change in the beneficial direction and changed from the clinical to the functional population. **Improved:** the individual showed a reliable change in the beneficial direction but did not change populations. **Unchanged:** the individual showed no reliable change. **Deteriorated:** the individual showed a reliable change in the disadvantageous direction but did not change populations. **Harmed:** the individual showed a reliable change in the disadvantageous direction and switched from the functional to the clinical population

#### Supplementary Table 5 - Experience of Service Use Questionnaire

| Experience of service use questionnaire* | Caregiver | | | | Self report | | | |
|---|---|---|---|---|---|---|---|---|
| | N | Certainly true | Partly true | Not true | N | Certainly true | Partly true | Not true |
| **Anxiety treatment protocol** | | | | | | | | |
| I feel that the people who have seen me/my child listened to me/my child | 44 | 43 (98%) | 1 (2.3%) | 0 (0%) | 31 | 28 (90%) | 3 (9.7%) | 0 (0%) |
| It was easy to talk to the people who have seen me/my child | 44 | 43 (98%) | 1 (2.3%) | 0 (0%) | 34 | 26 (76%) | 6 (18%) | 2 (5.9%) |
| I was treated well by the people who have seen me/my child | 44 | 43 (98%) | 1 (2.3%) | 0 (0%) | 34 | 31 (91%) | 3 (8.8%) | 0 (0%) |
| My views and worries were taken seriously | 44 | 42 (95%) | 2 (4.5%) | 0 (0%) | 27 | 25 (93%) | 0 (0%) | 2 (7.4%) |
| I feel the people at that service know how to help with the problem I went for | 44 | 42 (95%) | 2 (4.5%) | 0 (0%) | 33 | 30 (91%) | 3 (9.1%) | 0 (0%) |
| I have been given enough explanation about the help available at that service | 44 | 41 (93%) | 3 (6.8%) | 0 (0%) | 34 | 27 (79%) | 7 (21%) | 0 (0%) |
| I feel that the people who have seen me/my child are working together to help with the problem(s) | 43 | 41 (95%) | 2 (4.7%) | 0 (0%) | 28 | 26 (93%) | 2 (7.1%) | 0 (0%) |
| The facilities at that service were comfortable (e.g. waiting area) | 44 | 34 (77%) | 9 (20%) | 1 (2.3%) | 33 | 22 (67%) | 11 (33%) | 0 (0%) |
| The appointments were usually at a convenient time (e.g. didn't interfere with work, school) | 44 | 37 (84%) | 6 (14%) | 1 (2.3%) | 33 | 22 (67%) | 10 (30%) | 1 (3.0%) |
| It was quite easy to get to the place where the appointments were | 44 | 39 (89%) | 5 (11%) | 0 (0%) | 33 | 21 (64%) | 10 (30%) | 2 (6.1%) |
| If a friend needed similar help, I would recommend that he or she go to that service | 44 | 43 (98%) | 1 (2.3%) | 0 (0%) | 33 | 29 (88%) | 3 (9.1%) | 1 (3.0%) |
| Overall, the help I have received at that service was good | 44 | 43 (98%) | 1 (2.3%) | 0 (0%) | 32 | 32 (100%) | 0 (0%) | 0 (0%) |
| **Depression treatment protocol** | | | | | | | | |
| I feel that the people who have seen me/my child listened to me/my child | 19 | 19 (100%) | 0 (0%) | 0 (0%) | 17 | 16 (94%) | 1 (5.9%) | 0 (0%) |
| It was easy to talk to the people who have seen me/my child | 19 | 18 (95%) | 1 (5.3%) | 0 (0%) | 19 | 11 (58%) | 7 (37%) | 1 (5.3%) |
| I was treated well by the people who have seen me/my child | 19 | 19 (100%) | 0 (0%) | 0 (0%) | 18 | 16 (89%) | 1 (5.6%) | 1 (5.6%) |
| My views and worries were taken seriously | 19 | 18 (95%) | 1 (5.3%) | 0 (0%) | 19 | 16 (84%) | 1 (5.3%) | 2 (11%) |
| I feel the people at that service know how to help with the problem I went for | 19 | 18 (95%) | 1 (5.3%) | 0 (0%) | 17 | 12 (71%) | 4 (24%) | 1 (5.9%) |
| I have been given enough explanation about the help available at that service | 19 | 19 (100%) | 0 (0%) | 0 (0%) | 18 | 13 (72%) | 4 (22%) | 1 (5.6%) |
| I feel that the people who have seen me/my child are working together to help with the problem(s) | 19 | 19 (100%) | 0 (0%) | 0 (0%) | 15 | 12 (80%) | 2 (13%) | 1 (6.7%) |
| The facilities at that service were comfortable (e.g. waiting area) | 19 | 16 (84%) | 3 (16%) | 0 (0%) | 18 | 10 (56%) | 6 (33%) | 2 (11%) |
| The appointments were usually at a convenient time (e.g. didn't interfere with work, school) | 19 | 16 (84%) | 1 (5.3%) | 2 (11%) | 18 | 11 (61%) | 6 (33%) | 1 (5.6%) |

| Experience of service use questionnaire* | Caregiver | | | | Self report | | | |
|---|---|---|---|---|---|---|---|---|
| | **N** | **Certainly true** | **Partly true** | **Not true** | **N** | **Certainly true** | **Partly true** | **Not true** |
| It was quite easy to get to the place where the appointments were | 19 | 17 (89%) | 2 (11%) | 0 (0%) | 18 | 13 (72%) | 3 (17%) | 2 (11%) |
| If a friend needed similar help, I would recommend that he or she go to that service | 19 | 18 (95%) | 1 (5.3%) | 0 (0%) | 18 | 13 (72%) | 5 (28%) | 0 (0%) |
| Overall, the help I have received at that service was good | 19 | 19 (100%) | 0 (0%) | 0 (0%) | 17 | 16 (94%) | 0 (0%) | 1 (5.9%) |
| **Behavioural parent training protocol** | **N** | **Certainly true** | **Partly true** | **Not true** | **N** | **Certainly true** | **Partly true** | **Not true** |
| I feel that the people who have seen me/my child listened to me/my child | 48 | 48 (100%) | 0 (0%) | 0 (0%) | 8 | 5 (63%) | 1 (13%) | 2 (25%) |
| It was easy to talk to the people who have seen me/my child | 48 | 47 (98%) | 1 (2.1%) | 0 (0%) | 9 | 4 (44%) | 4 (44%) | 1 (11%) |
| I was treated well by the people who have seen me/my child | 48 | 48 (100%) | 0 (0%) | 0 (0%) | 9 | 7 (78%) | 1 (11%) | 1 (11%) |
| My views and worries were taken seriously | 48 | 48 (100%) | 0 (0%) | 0 (0%) | 8 | 7 (88%) | 1 (13%) | 0 (0%) |
| I feel the people at that service know how to help with the problem I went for | 48 | 48 (100%) | 0 (0%) | 0 (0%) | 9 | 4 (44%) | 4 (44%) | 1 (11%) |
| I have been given enough explanation about the help available at that service | 48 | 46 (96%) | 2 (4.2%) | 0 (0%) | 9 | 6 (67%) | 3 (33%) | 0 (0%) |
| I feel that the people who have seen me/my child are working together to help with the problem(s) | 48 | 43 (90%) | 5 (10%) | 0 (0%) | 9 | 4 (44%) | 3 (33%) | 2 (22%) |
| The facilities at that service were comfortable (e.g. waiting area) | 48 | 36 (75%) | 12 (25%) | 0 (0%) | 9 | 5 (56%) | 4 (44%) | 0 (0%) |
| The appointments were usually at a convenient time (e.g. didn't interfere with work, school) | 48 | 44 (92%) | 3 (6.3%) | 1 (2.1%) | 9 | 3 (33%) | 4 (44%) | 2 (22%) |
| It was quite easy to get to the place where the appointments were | 48 | 37 (77%) | 9 (19%) | 2 (4.2%) | 8 | 6 (75%) | 1 (13%) | 1 (13%) |
| If a friend needed similar help, I would recommend that he or she go to that service | 48 | 48 (100%) | 0 (0%) | 0 (0%) | 9 | 6 (67%) | 0 (0%) | 3 (33%) |
| Overall, the help I have received at that service was good | 48 | 47 (98%) | 1 (2.1%) | 0 (0%) | 7 | 5 (71%) | 1 (14%) | 1 (14%) |

**Note:** The wording used specifically for this table was adapted from the original questionnaires presented to participants, which employ age-specific phrasing to ensure that items are appropriate and comprehensible for the target developmental stage.

### Supplementary Table 6 - Protocol assessment by trainees

| **ANXIETY TRAINING** | Average | Per session | | | | | | | | | | | | | |
|---|---|---|---|---|---|---|---|---|---|---|---|---|---|---|---|
| | % satisfied OR very satisfied | S1 | S2 | S3 | S4 | S5 | S6 | S7 | S8 | S9 | S10 | S11 | S12 | S13 | S14 |
| **Material & Content** | | N=20 | N=20 | N=20 | N=20 | N=19 | N=19 | N=18 | N=18 | N=18 | N=18 | N=18 | N=18 | N=17 | N=17 |
| Time available | 88% | 70% | 95% | 95% | 85% | 68% | 95% | 94% | 89% | 94% | 83% | 89% | 89% | 88% | 94% |
| Understandable | 98% | 90% | 100% | 95% | 95% | 95% | 100% | 100% | 100% | 100% | 100% | 100% | 100% | 100% | 100% |
| Cultural alignment | 98% | 95% | 100% | 100% | 100% | 100% | 100% | 100% | 100% | 94% | 100% | 94% | 94% | 100% | 100% |
| Clear instructions | 98% | 90% | 100% | 100% | 100% | 95% | 100% | 100% | 100% | 100% | 100% | 100% | 94% | 100% | 100% |
| **Training & supervision evaluation** | | | | | | | | | | | | | | | |
| Understandable instruction | 95% | 95% | 95% | 95% | 95% | 95% | 100% | 94% | 94% | 94% | 94% | 94% | 94% | 94% | 94% |
| Helped me effectively apply techniques | 95% | 95% | 95% | 95% | 95% | 95% | 100% | 94% | 94% | 94% | 94% | 94% | 94% | 94% | 94% |
| Offered supportive and constructive feedback | 95% | 95% | 95% | 95% | 95% | 95% | 100% | 94% | 94% | 94% | 94% | 94% | 94% | 94% | 94% |
| Created an environment of cooperation and trust | 95% | 95% | 95% | 95% | 95% | 95% | 100% | 94% | 94% | 94% | 94% | 94% | 94% | 94% | 94% |
| Encouraged me to make independent decisions | 95% | 95% | 95% | 95% | 95% | 95% | 100% | 94% | 94% | 94% | 94% | 94% | 94% | 94% | 94% |

| **DEPRESSION TRAINING** | Average | Per session | | | | | | | | | | | | | |
|---|---|---|---|---|---|---|---|---|---|---|---|---|---|---|---|
| | % satisfied OR very satisfied | S1 | S2 | S3 | S4 | S5 | S6 | S7 | S8 | S9 | S10 | S11 | S12 | S13 | S14 |
| **Material & Content** | | N=16 | N=17 | N=17 | N=17 | N=17 | N=17 | N=17 | N=17 | N=17 | N=17 | N=17 | N=17 | N=17 | N=17 |
| Time available | 84% | 81% | 65% | 82% | 94% | 82% | 82% | 76% | 76% | 82% | 82% | 88% | 88% | 88% | 100% |
| Understandable | 94% | 100% | 94% | 94% | 94% | 94% | 100% | 88% | 76% | 88% | 94% | 100% | 94% | 94% | 100% |
| Cultural alignment | 97% | 100% | 88% | 88% | 100% | 94% | 100% | 100% | 94% | 94% | 100% | 100% | 100% | 100% | 100% |
| Clear instructions | 89% | 88% | 94% | 94% | 94% | 82% | 88% | 82% | 76% | 82% | 94% | 88% | 94% | 88% | 100% |
| **Training & supervision evaluation** | | | | | | | | | | | | | | | |
| Understandable instruction | 98% | 94% | 100% | 100% | 100% | 100% | 94% | 94% | 94% | 100% | 100% | 100% | 100% | 100% | 100% |
| Helped me effectively apply techniques | 97% | 100% | 94% | 100% | 100% | 100% | 94% | 88% | 94% | 94% | 100% | 100% | 100% | 100% | 100% |
| Offered supportive and constructive feedback | 98% | 100% | 100% | 100% | 100% | 100% | 88% | 88% | 100% | 100% | 100% | 100% | 100% | 100% | 100% |
| Created an environment of cooperation and trust | 100% | 100% | 100% | 100% | 100% | 100% | 100% | 94% | 100% | 100% | 100% | 100% | 100% | 100% | 100% |
| Encouraged me to make independent decisions | 97% | 88% | 100% | 100% | 100% | 100% | 94% | 94% | 88% | 100% | 100% | 100% | 100% | 94% | 94% |

| BEHAVIOUR PARENT TRAINING PROTOCOL | Average | Per session | | | | | | | | | | | | | | |
|---|---|---|---|---|---|---|---|---|---|---|---|---|---|---|---|---|
| | % satisfied OR very satisfied | S1<br>N=33 | S2<br>N=32 | S3<br>N=32 | S4<br>N=32 | S5<br>N=30 | S6<br>N=31 | S7<br>N=30 | S8<br>N=30 | S9<br>N=30 | S10<br>N=30 | S11<br>N=30 | S12<br>N=30 | S13<br>N=30 | S14<br>N=30 | S15<br>N=30 |
| **Material & Content** | | | | | | | | | | | | | | | | |
| Time available | 84% | 64% | 59% | 81% | 78% | 93% | 84% | 80% | 87% | 77% | 90% | 90% | 93% | 93% | 97% | 97% |
| Understandable | 96% | 94% | 100% | 97% | 100% | 93% | 97% | 93% | 93% | 97% | 97% | 90% | 97% | 97% | 97% | 97% |
| Cultural alignment | 91% | 88% | 94% | 91% | 91% | 93% | 87% | 90% | 87% | 93% | 93% | 90% | 97% | 93% | 93% | 87% |
| Clear instructions | 95% | 94% | 94% | 97% | 97% | 90% | 94% | 93% | 97% | 93% | 97% | 90% | 97% | 97% | 97% | 97% |
| **Training & supervision evaluation** | | | | | | | | | | | | | | | | |
| Understandable instruction | 99% | 100% | 100% | 100% | 100% | 100% | 97% | 100% | 100% | 97% | 100% | 97% | 100% | 100% | 97% | 100% |
| Helped me effectively apply techniques | 99% | 100% | 97% | 100% | 100% | 93% | 97% | 100% | 97% | 97% | 100% | 100% | 100% | 100% | 100% | 100% |
| Offered supportive and constructive feedback | 100% | 100% | 100% | 100% | 100% | 100% | 100% | 100% | 100% | 100% | 100% | 100% | 100% | 100% | 100% | 100% |
| Created an environment of cooperation and trust | 100% | 100% | 100% | 100% | 100% | 100% | 100% | 100% | 100% | 100% | 100% | 100% | 100% | 100% | 100% | 100% |
| Encouraged me to make independent decisions | 93% | 97% | 97% | 94% | 94% | 93% | 90% | 90% | 93% | 90% | 93% | 93% | 93% | 93% | 93% | 93% |

### Supplementary Table 7 - Supervisor assessment: trainees`achievement of the therapy session's goals

| | Average | Per session | | | | | | | | | | | | | |
|---|---|---|---|---|---|---|---|---|---|---|---|---|---|---|---|
| **ANXIETY TRAINING** | % satisfied OR very satisfied | S1 N=26 | S2 N=28 | S3 N=27 | S4 N=27 | S5 N=26 | S6 N=25 | S7 N=25 | S8 N=26 | S9 N=27 | S10 N=23 | S11 N=26 | S12 N=25 | S13 N=23 | S14 N=23 |
| Adherence to session goals | 86% | 90% | 96% | 96% | 96% | 69% | 96% | 86% | 77% | 89% | 87% | 80% | 80% | 70% | 91% |
| Effective session time management | 92% | 83% | 93% | 96% | 93% | 85% | 100% | 90% | 92% | 93% | 96% | 90% | 90% | 91% | 91% |
| Homework review with clear feedback | 83% | | | 85% | 96% | 73% | 92% | 78% | 80% | 80% | 80% | 70% | 76% | 70% | 96% |
| Efficient homework assignment | 94% | 92% | 96% | 100% | 100% | 92% | 100% | 90% | 92% | 93% | 96% | 92% | 88% | 91% | |
| Proper use of session worksheets | 97% | 96% | 100% | 96% | 96% | 96% | 100% | 90% | 96% | 96% | 100% | 97% | 92% | 96% | 100% |
| Adaptation to patient and family needs | 97% | 100% | 100% | 100% | 100% | 85% | 100% | 100% | 92% | 96% | 96% | 100% | 96% | 96% | 100% |

| **DEPRESSION TRAINING** | Average | Per session | | | | | | | | | | | | | | |
|---|---|---|---|---|---|---|---|---|---|---|---|---|---|---|---|---|
| **Material & Content** | % satisfied OR very satisfied | S1 N=25 | S2 N=22 | S3 N=22 | S4 N=20 | S5 N=21 | S6 N=20 | S7 N=20 | S8 N=20 | S9 N=20 | S10 N=19 | S11 N=20 | S12 N=20 | S13 N=19 | S14 N=20 | S15 N=16 |
| Adherence to session goals | 97% | 92% | 100% | 95% | 95% | 100% | 95% | 100% | 95% | 100% | 100% | 100% | 95% | 100% | 95% | 100% |
| Effective session time management | 97% | 88% | 95% | 95% | 95% | 100% | 100% | 100% | 100% | 95% | 95% | 100% | 95% | 100% | 100% | 100% |
| Homework review with clear feedback | 97% | | 91% | | 100% | 100% | 95% | 100% | 95% | 100% | 93% | 100% | 95% | 100% | 95% | |
| Efficient homework assignment | 96% | 96% | 91% | | 95% | 100% | 95% | 100% | 95% | 95% | 100% | 100% | 95% | 100% | 94% | |
| Proper use of session worksheets | 99% | 100% | 100% | 100% | 95% | 100% | 95% | 100% | 100% | 100% | 100% | 100% | 100% | 100% | 95% | 100% |
| Adaptation to patient and family needs | 99% | 100% | 100% | 100% | 95% | 100% | 100% | 100% | 100% | 100% | 100% | 100% | 95% | 100% | 100% | 100% |

| | Average | Per session | | | | | | | | | | | | | | |
|---|---|---|---|---|---|---|---|---|---|---|---|---|---|---|---|---|
| **BEHAVIOUR PARENT TRAINING PROTOCOL** | % satisfied OR very satisfied | S1 N=51 | S2 N=49 | S3 N=61 | S4 N=51 | S5 N=47 | S6 N=49 | S7 N=49 | S8 N=49 | S9 N=49 | S10 N=49 | S11 N=49 | S12 N=49 | S13 N=48 | S14 N=49 | S15 N=49 |
| Time available | 84% | 64% | 59% | 81% | 78% | 93% | 84% | 80% | 87% | 77% | 90% | 90% | 93% | 93% | 97% | 97% |
| Understandable | 96% | 94% | 100% | 97% | 100% | 93% | 97% | 93% | 93% | 97% | 97% | 90% | 97% | 97% | 97% | 97% |
| Cultural alignment | 91% | 88% | 94% | 91% | 91% | 93% | 87% | 90% | 87% | 93% | 93% | 90% | 97% | 93% | 93% | 87% |
| Clear instructions | 95% | 94% | 94% | 97% | 97% | 90% | 94% | 93% | 97% | 93% | 97% | 90% | 97% | 97% | 97% | 97% |
| **Training & supervision evaluation** | | | | | | | | | | | | | | | | |

| | | | | | | | | | | | | | | | | |
|---|---|---|---|---|---|---|---|---|---|---|---|---|---|---|---|---|
| Understandable instruction | 99% | 100% | 100% | 100% | 100% | 100% | 97% | 100% | 100% | 97% | 100% | 97% | 100% | 100% | 97% | 100% |
| Helped me effectively apply techniques | 99% | 100% | 97% | 100% | 100% | 93% | 97% | 100% | 97% | 97% | 100% | 100% | 100% | 100% | 100% | 100% |
| Offered supportive and constructive feedback | 100% | 100% | 100% | 100% | 100% | 100% | 100% | 100% | 100% | 100% | 100% | 100% | 100% | 100% | 100% | 100% |
| Created an environment of cooperation and trust | 100% | 100% | 100% | 100% | 100% | 100% | 100% | 100% | 100% | 100% | 100% | 100% | 100% | 100% | 100% | 100% |
| Encouraged me to make independent decisions | 93% | 97% | 97% | 94% | 94% | 93% | 90% | 90% | 93% | 90% | 93% | 93% | 93% | 93% | 93% | 93% |

Supplementary Table 8: Anxiety treatment protocol: session-level item completion and difficulty (trainee)

| ANXIETY TREATMENT PROTOCOL: PILOT 1 | Item completion n (%) | Very easy | Easy | Neutral | Difficult | Very difficulty |
|---|---|---|---|---|---|---|
| Session 1 | | | | | | |
| G1a. I introduced myself and praised parents for engaging in their child's treatment | 5 (100%) | 5 (100%) | 0 (0%) | 0 (0%) | 0 (0%) | 0 (0%) |
| G1b. I oriented parents to the steps of today's session, in advance, following the manual's script | 5 (100%) | 2 (40%) | 3 (60%) | 0 (0%) | 0 (0%) | 0 (0%) |
| G2a. I explained the scope of this treatment (i.e., including duration, frequency, parent participation, confidentiality rules and boundaries of interventions). | 5 (100%) | 4 (80%) | 1 (20%) | 0 (0%) | 0 (0%) | 0 (0%) |
| G2b. I explained the concept of the negative reinforcement cycle using the "Negative Reinforcement Cycle graphic" in their workbook. | 5 (100%) | 2 (40%) | 2 (40%) | 1 (20%) | 0 (0%) | 0 (0%) |
| G3. I worked with the parents to define two reasonable, measurable and specific goals for their child's treatment, using the "Parent Goal Setting" worksheet | 5 (100%) | 2 (40%) | 2 (40%) | 1 (20%) | 0 (0%) | 0 (0%) |
| G4. I explained the rationale and how implementing the CBT model will elevate the probability of the child's anxiety symptoms to decrease. | 5 (100%) | 2 (40%) | 2 (40%) | 1 (20%) | 0 (0%) | 0 (0%) |
| G5. I asked for the parents feedback in the end, including which parts were more difficult for them to understand, which parts they enjoyed and/or found more helpful. | 5 (100%) | 2 (40%) | 2 (40%) | 1 (20%) | 0 (0%) | 0 (0%) |
| Session 2 | | | | | | |
| G1. I introduced the child to the therapy process by explaining the purpose of our sessions, the nature of this therapy, and by outlining the structure for our future meetings. | 5 (100%) | 4 (80%) | 1 (20%) | 0 (0%) | 0 (0%) | 0 (0%) |
| G2. I used the "All About Me" activity worksheet and/or involved the child in playing one of the games outlined in the manual to foster a therapeutic connection. | 5 (100%) | 4 (80%) | 1 (20%) | 0 (0%) | 0 (0%) | 0 (0%) |
| G3. I drew upon a shared experience from the child's past (i.e. learning to swim, etc.), to convey that consistent practice and perseverance with initially challenging tasks lead to improvement over time. | 5 (100%) | 2 (40%) | 3 (60%) | 0 (0%) | 0 (0%) | 0 (0%) |
| G4. I engaged the child in a discussion about their goals for treatment using the Goals Worksheet | 5 (100%) | 2 (40%) | 3 (60%) | 0 (0%) | 0 (0%) | 0 (0%) |
| Session 3 | | | | | | |
| G1a. I provided a brief review of last week's home assignment and encouraged the child to collaboratively address any incomplete aspects during today's session (i.e., "All About Me" worksheet) | 5 (100%) | 3 (60%) | 2 (40%) | 0 (0%) | 0 (0%) | 0 (0%) |
| G1b. I discussed with the child about six fundamental emotions (joy, anger, sadness, disgust, fear, excitement) and elucidated their roles in our daily experiences. | 5 (100%) | 3 (60%) | 2 (40%) | 0 (0%) | 0 (0%) | 0 (0%) |

| | | | | | | |
|---|---|---|---|---|---|---|
| G2a. I supported the child in articulating accurate descriptions of their emotions to enhance their understanding of associated feelings, employing the Emotion Wheel illustration | 5 (100%) | 3 (60%) | 2 (40%) | 0 (0%) | 0 (0%) | 0 (0%) |
| G2b. I employed the Emotions Thermometer worksheet to help the child practice assessing the intensity of their emotions | 5 (100%) | 4 (80%) | 1 (20%) | 0 (0%) | 0 (0%) | 0 (0%) |
| G3. I helped the child in recognizing specific intense and unhelpful emotions that are influencing their life, making situations more challenging, utilizing the Emotion Wheel worksheet once again | 5 (100%) | 3 (60%) | 2 (40%) | 0 (0%) | 0 (0%) | 0 (0%) |
| G4. I introduced the Emotion Log worksheet and conveyed to the child that, throughout therapy, they would acquire tools and coping skills to effectively manage significant emotions on a daily basis | 5 (100%) | 3 (60%) | 2 (40%) | 0 (0%) | 0 (0%) | 0 (0%) |
| Session 4 | | | | | | |
| G1a. I provided a brief review of last week's home assignment and encouraged the child to collaboratively address any incomplete aspects during today's session (i.e., Emotion Log, Identifying Feelings Worksheets) | 5 (100%) | 3 (60%) | 2 (40%) | 0 (0%) | 0 (0%) | 0 (0%) |
| G1b. I offered psychoeducation on anxiety, elucidating its adaptive and beneficial role in protecting us. Simultaneously, I highlighted how it can occasionally induce distress by signaling danger even in situations where everything is genuinely fine. | 5 (100%) | 4 (80%) | 0 (0%) | 1 (20%) | 0 (0%) | 0 (0%) |
| G2. I helped the child in comprehending that the overarching objective of this therapy is not to eliminate anxiety but rather to enhance the child's skills to effectively manage and cope with it. | 5 (100%) | 4 (80%) | 1 (20%) | 0 (0%) | 0 (0%) | 0 (0%) |
| G3. I engaged in a conversation with the child regarding their experience of anxiety and assisted them in comprehending its physical manifestations by utilizing the "Where I Feel Anxiety" worksheet | 5 (100%) | 3 (60%) | 2 (40%) | 0 (0%) | 0 (0%) | 0 (0%) |
| G4. I discussed with the child about how certain thoughts and emotions, like fear, can lead to physical responses such as a fast heartbeat, sweating, shaking, evoking instinctual behaviors associated with fight, flight, or freeze. | 5 (100%) | 4 (80%) | 1 (20%) | 0 (0%) | 0 (0%) | 0 (0%) |
| G5. I assisted the child in comprehending and implementing the concept of externalizing anxiety, providing relevant examples and collaboratively coming up with a nickname to distinguish the anxiety from themselves | 5 (100%) | 5 (100%) | 0 (0%) | 0 (0%) | 0 (0%) | 0 (0%) |
| G6. I outlined the purpose of building a collaborative team (clinician, child, family) to collectively fight against the child's anxiety, rather than fighting against the child, emphasizing empowerment and confidence-building. | 5 (100%) | 3 (60%) | 1 (20%) | 1 (20%) | 0 (0%) | 0 (0%) |
| G7. I emphasized to the child that our joint efforts will gradually combat their anxiety in a way that feels manageable for them, promoting long-term well-being | 5 (100%) | 2 (40%) | 3 (60%) | 0 (0%) | 0 (0%) | 0 (0%) |
| Session 5 | | | | | | |
| G1a. I provided a brief review of last parents' meeting discussion and first home assignment for parents (i.e., Parent Accommodation Log worksheet) | 5 (100%) | 2 (40%) | 2 (40%) | 1 (20%) | 0 (0%) | 0 (0%) |
| G1b. I discussed with the parents about effective behavioral strategies, including techniques like differential attention, removing accommodations, and responding thoughtfully to reassurance-seeking questions, utilizing The Power of your Attention worksheet | 5 (100%) | 2 (40%) | 1 (20%) | 2 (40%) | 0 (0%) | 0 (0%) |

| | | | | | | |
|---|---|---|---|---|---|---|
| G2. I supported parents in consistently applying targeted strategies at home by working together to create a priority list for their child's behaviors and offering very specific examples (e.g., checking the child's closet only once at bedtime) | 5 (100%) | 1 (20%) | 4 (80%) | 0 (0%) | 0 (0%) | 0 (0%) |
| G3. I assisted parents to address potential obstacles in implementing identified behavior changes at home and discussed any concerns they had about successfully carrying out these changes. | 5 (100%) | 1 (20%) | 3 (60%) | 1 (20%) | 0 (0%) | 0 (0%) |
| Session 6 | | | | | | |
| G1a. I provided a brief review of last week's home assignment and encouraged the child to collaboratively address any incomplete aspects during today's session (i.e., Where I Feel Anxiety worksheet and finding a name for their anxiety) | 5 (100%) | 3 (60%) | 2 (40%) | 0 (0%) | 0 (0%) | 0 (0%) |
| G1b. I assisted the child in grasping the connection between thoughts, feelings, and behaviors by utilizing the CBT Triangle and the CBT Matching Game worksheets | 5 (100%) | 3 (60%) | 2 (40%) | 0 (0%) | 0 (0%) | 0 (0%) |
| G2. I discussed with the child that to alter the trajectory of our thoughts, feelings, and behaviors, the initial step is to modify our actions, utilizing again the CBT Triangle worksheet | 5 (100%) | 4 (80%) | 1 (20%) | 0 (0%) | 0 (0%) | 0 (0%) |
| G3. I helped the child recognize and comprehend behaviors that reinforce anxiety (like accommodation, reassurance-seeking, avoidance, control, and other safety behaviors) or, alternatively, confront anxiety in the long run. | 5 (100%) | 4 (80%) | 1 (20%) | 0 (0%) | 0 (0%) | 0 (0%) |
| G4. Together with the child, we generated a list of behaviors that either perpetuate or challenge anxiety, utilizing the My Behaviors and Anxiety Worksheet collaboratively. | 5 (100%) | 4 (80%) | 0 (0%) | 1 (20%) | 0 (0%) | 0 (0%) |
| Session 7 | | | | | | |
| G1. I provided a brief review of the last home assignment (i.e., The Power of Your Attention) and addressed potential challenges in implementing initial parenting changes. | 5 (100%) | 2 (40%) | 2 (40%) | 1 (20%) | 0 (0%) | 0 (0%) |
| G2. I helped parents grasp behavioral exposure practices using the Worry Wave Worksheet and the Fear Ladder | 5 (100%) | 2 (40%) | 2 (40%) | 1 (20%) | 0 (0%) | 0 (0%) |
| G3. We reviewed how parents can model and teach bravery to children and the importance of parents modeling brave behaviors. | 5 (100%) | 2 (40%) | 1 (20%) | 2 (40%) | 0 (0%) | 0 (0%) |
| G4. We discussed whether a reward system would be helpful for the child in facing their anxiety and which reward system would be appropriate for the family. | 5 (100%) | 2 (40%) | 1 (20%) | 2 (40%) | 0 (0%) | 0 (0%) |
| Session 8 | | | | | | |
| G1. I provided a brief review of last week's home assignment and encouraged the child to collaboratively address any incomplete aspects during today's session (i.e., The Worry Wave Worksheet, Fear Thermometer Worksheet, and Fear Ladder Worksheet) | 5 (100%) | 3 (60%) | 2 (40%) | 0 (0%) | 0 (0%) | 0 (0%) |
| G2. I reoriented the child to behavioral-based work and created a fear ladder to work with the child in the office using the Fear Ladder Worksheet | 5 (100%) | 2 (40%) | 2 (40%) | 1 (20%) | 0 (0%) | 0 (0%) |
| G3. I let the child choose a low rated practice to start their first exposure in vivo in the office using the SUDS scale | 5 (100%) | 2 (40%) | 1 (20%) | 2 (40%) | 0 (0%) | 0 (0%) |

| | | | | | | |
|---|---|---|---|---|---|---|
| G4a. We troubleshoot any challenges that the child might have encountered during the exposure practice using the tips from the manual. | 5 (100%) | 2 (40%) | 1 (20%) | 2 (40%) | 0 (0%) | 0 (0%) |
| G4b. I helped the child get ready to continue their exposure therapy at home by involving their parents. Together, we planned for a homework assignment (i.e., Exposure Tracking Worksheet). | 5 (100%) | 2 (40%) | 0 (0%) | 3 (60%) | 0 (0%) | 0 (0%) |
| Session 9 | | | | | | |
| G1. I provided a brief review of last week's home assignment and encouraged the child to collaboratively address any incomplete aspects during today's session (i.e., Exposure Tracking Worksheet) | 5 (100%) | 3 (60%) | 2 (40%) | 0 (0%) | 0 (0%) | 0 (0%) |
| G2. I introduced to the child cognitive coping skills, as a way to fight back against anxiety. | 5 (100%) | 2 (40%) | 3 (60%) | 0 (0%) | 0 (0%) | 0 (0%) |
| G3. I promoted disengagement with the thoughts and worked with the child to empower them to challenge and manage their thoughts effectively. | 5 (100%) | 2 (40%) | 3 (60%) | 0 (0%) | 0 (0%) | 0 (0%) |
| G4. I introduced the child to the concept of thinking traps to prepare them for challenging their thoughts using the Thinking Traps Poster and Thinking Traps Worksheet. | 5 (100%) | 2 (40%) | 2 (40%) | 0 (0%) | 1 (20%) | 0 (0%) |
| G5. I introduced the child to the idea that anxiety can give children maladaptive thoughts. | 5 (100%) | 3 (60%) | 1 (20%) | 1 (20%) | 0 (0%) | 0 (0%) |
| G6. We worked together with the child to challenge maladaptive thoughts. | 5 (100%) | 2 (40%) | 3 (60%) | 0 (0%) | 0 (0%) | 0 (0%) |
| G7. We discussed ways that the child can replace an unhelpful thought with a more realistic one. | 5 (100%) | 2 (40%) | 3 (60%) | 0 (0%) | 0 (0%) | 0 (0%) |
| G8. I helped the child to build motivation to challenge anxious thoughts, practicing supportive self statements and using examples. | 5 (100%) | 2 (40%) | 3 (60%) | 0 (0%) | 0 (0%) | 0 (0%) |
| G9. I put it all together based on the 4 C's: Catch, Challenge, Cultivate, Change. | 5 (100%) | 2 (40%) | 1 (20%) | 2 (40%) | 0 (0%) | 0 (0%) |
| Session 10 | | | | | | |
| G1. I provided a brief review of last week's home assignment and encouraged the child to collaboratively address any incomplete aspects during today's session (i.e., Thinking Traps Home Assignment). | 5 (100%) | 3 (60%) | 2 (40%) | 0 (0%) | 0 (0%) | 0 (0%) |
| G2. I reviewed the child's exposure practice and set continued exposure goals | 5 (100%) | 2 (40%) | 2 (40%) | 1 (20%) | 0 (0%) | 0 (0%) |
| G3. We revisited the hierarchy from session 9, re-rated potential exposure, and gave tips for increasing the difficulty of exposure practices. | 5 (100%) | 2 (40%) | 2 (40%) | 1 (20%) | 0 (0%) | 0 (0%) |
| Session 11 | | | | | | |
| G1. I provided a brief review of last week's home assignment and encouraged the child to collaboratively address any incomplete aspects during today's session (i.e., Exposure Tracker Worksheet). | 5 (100%) | 4 (80%) | 1 (20%) | 0 (0%) | 0 (0%) | 0 (0%) |
| G2. We reviewed the instructions and guidance from session 11, and actively conducted in vivo exposure exercises with the child, using the Fear Ladder. | 5 (100%) | 3 (60%) | 2 (40%) | 0 (0%) | 0 (0%) | 0 (0%) |
| G3. We addressed challenges that have arisen by referencing the tips and strategies provided in sessions 9 and 11. | 5 (100%) | 3 (60%) | 0 (0%) | 2 (40%) | 0 (0%) | 0 (0%) |
| Session 12 | | | | | | |
| G1. I provided a brief review of last week's home assignment and encouraged the child to collaboratively address any incomplete aspects during today's session (i.e., Exposure Tracker Worksheet). | 5 (100%) | 3 (60%) | 2 (40%) | 0 (0%) | 0 (0%) | 0 (0%) |

| | | | | | | |
|---|---|---|---|---|---|---|
| G2. We reviewed the Emotion Wheel and talked with the child about BIG emotions using the Feelings Thermometer | 5 (100%) | 3 (60%) | 2 (40%) | 0 (0%) | 0 (0%) | 0 (0%) |
| G3. I explained to the child the connection between the brain and body, while we also briefly reviewed the Coping Cards Worksheet and the skills outlined on this worksheet. | 5 (100%) | 3 (60%) | 1 (20%) | 1 (20%) | 0 (0%) | 0 (0%) |
| G4. I had the child practice deep breathing in session using the Breathing Cards worksheet. | 5 (100%) | 4 (80%) | 1 (20%) | 0 (0%) | 0 (0%) | 0 (0%) |
| G5. I prepared the child for the upcoming end of treatment and let them express their emotions and thoughts about it. | 5 (100%) | 3 (60%) | 1 (20%) | 1 (20%) | 0 (0%) | 0 (0%) |
| Session 13 | | | | | | |
| G1. I focused on reviewing the child's and family's experience and practice with exposure so far. If applicable, I evaluated the reward system implemented during this process. | 4 (100%) | 1 (25%) | 1 (25%) | 2 (50%) | 0 (0%) | 0 (0%) |
| G2. We created a fear ladder for the child's next exposures based on recent experiences. With the parents, I set goals for the child's progress and outlined the steps to reach those goals. | 3 (75%) | 1 (33%) | 1 (33%) | 0 (0%) | 1 (33%) | 0 (0%) |
| G3. I discussed potential barriers to implementation and gave troubleshooting tips for exposure. | 4 (100%) | 1 (25%) | 2 (50%) | 0 (0%) | 1 (25%) | 0 (0%) |
| G4. I followed up on the previous session focused on building a brave lifestyle, gauging the parents' implementation progress. We revisited the key principles of leading a brave lifestyle if necessary. | 4 (100%) | 1 (25%) | 1 (25%) | 2 (50%) | 0 (0%) | 0 (0%) |
| G5. I prepared the family for the end of treatment and helped them develop a plan for maintaining the progress made in therapy. | 4 (100%) | 2 (50%) | 1 (25%) | 0 (0%) | 1 (25%) | 0 (0%) |
| G6. I thoroughly went over the "Red Flags" list with the parents, guiding them through each item. I encouraged them to pinpoint any red flags they believe might arise in their particular situation. | 4 (100%) | 1 (25%) | 1 (25%) | 1 (25%) | 1 (25%) | 0 (0%) |
| Session 14 | | | | | | |
| G1. I provided a brief review of the last home assignment and encouraged the child to collaboratively address any incomplete aspects during today's session (i.e., Best Skill and 6 Senses skill worksheets) | 4 (100%) | 3 (75%) | 1 (25%) | 0 (0%) | 0 (0%) | 0 (0%) |
| G2. I engaged in a detailed conversation with the child regarding the conclusion of treatment, allowing them to express their emotions and thoughts about it. Furthermore, I evaluated any challenges they might foresee or currently encounter regarding this transition. | 4 (100%) | 3 (75%) | 0 (0%) | 1 (25%) | 0 (0%) | 0 (0%) |
| G3. I underscored the child's efforts and achievements, emphasizing that success can be measured in various ways, not solely by one criterion. | 4 (100%) | 3 (75%) | 1 (25%) | 0 (0%) | 0 (0%) | 0 (0%) |
| G4. We talked about any remaining goals the child is striving to achieve and outlined the subsequent steps in the fear hierarchy for their continued exposure. | 4 (100%) | 1 (25%) | 3 (75%) | 0 (0%) | 0 (0%) | 0 (0%) |
| G5. I introduced the child to the idea of a Brave Lifestyle. | 4 (100%) | 1 (25%) | 3 (75%) | 0 (0%) | 0 (0%) | 0 (0%) |
| G6. Using the Red Flags worksheet, I helped the child recognize their own indicators that might signal a need for further support. | 4 (100%) | 1 (25%) | 1 (25%) | 1 (25%) | 1 (25%) | 0 (0%) |
| G7. I presented the child with a graduation certificate and commended their effort throughout the treatment process. | 3 (75%) | 2 (67%) | 0 (0%) | 1 (33%) | 0 (0%) | 0 (0%) |

| ANXIETY TREATMENT PROTOCOL: PILOT 2 | Item completion n (%) | Very easy | Easy | Neutral | Difficult | Very difficult y |
|---|---|---|---|---|---|---|
| Session 1 | | | | | | |
| G1a. I followed the steps of building the therapeutic relationship in order to lay the foundation of our collaboration ("Building the therapeutic relationship-What the therapist needs to consider") | 15 (100%) | 6 (40%) | 8 (53%) | 1 (6.7%) | 0 (0%) | 0 (0%) |
| G2a. I explained the general framework for the application of this treatment. | 15 (100%) | 6 (40%) | 8 (53%) | 1 (6.7%) | 0 (0%) | 0 (0%) |
| G2b. I explained the concept of the negative reinforcement cycle ("Negative reinforcement cycle" in their workbook). | 15 (100%) | 6 (40%) | 6 (40%) | 3 (20%) | 0 (0%) | 0 (0%) |
| G3a.I worked with the parents and child to establish , specific goals for the child's treatment (Parent Goals and Child Goals worksheets. | 15 (100%) | 3 (20%) | 9 (60%) | 2 (13%) | 1 (6.7%) | 0 (0%) |
| G4a. I explained the rationale for the GST model and how its implementation would increase the likelihood of improving the child's anxiety symptoms. | 15 (100%) | 5 (33%) | 6 (40%) | 4 (27%) | 0 (0%) | 0 (0%) |
| G5a. I asked for feedback from the parents and the child. | 15 (100%) | 5 (33%) | 8 (53%) | 2 (13%) | 0 (0%) | 0 (0%) |
| Session 2 | | | | | | |
| G1a. I used and/or involved the child in one of the games described in the manual to reinforce the therapeutic connection (worksheet of the "Everything about me" activity). | 15 (100%) | 9 (60%) | 5 (33%) | 1 (6.7%) | 0 (0%) | 0 (0%) |
| G2a. I stressed the importance of practice and perseverance in activities that are initially difficult but improve over time. | 15 (100%) | 4 (27%) | 7 (47%) | 3 (20%) | 1 (6.7%) | 0 (0%) |
| G3a. I engaged the child in a discussion about their goals for treatment (goals worksheet). | 15 (100%) | 3 (20%) | 8 (53%) | 2 (13%) | 2 (13%) | 0 (0%) |
| Session 3 | | | | | | |
| G1a. I did a brief review of the homework for the homework. | 15 (100%) | 6 (40%) | 7 (47%) | 1 (6.7%) | 1 (6.7%) | 0 (0%) |
| G1b. I discussed with the child about the six basic emotions and their role in our daily experiences. | 15 (100%) | 5 (33%) | 9 (60%) | 1 (6.7%) | 0 (0%) | 0 (0%) |
| G2a. I supported the child to formulate accurate descriptions of his feelings (illustration of the "Wheel of Feelings"). | 15 (100%) | 4 (27%) | 8 (53%) | 2 (13%) | 1 (6.7%) | 0 (0%) |
| G2b. I helped the child to practise assessing the intensity of his/her emotions (worksheet "The Emotions Thermometer"). | 15 (100%) | 3 (20%) | 10 (67%) | 1 (6.7%) | 1 (6.7%) | 0 (0%) |
| G3a. I helped the child to identify specific intense and unhelpful emotions (Emotions Wheel worksheet). | 15 (100%) | 1 (6.7%) | 11 (73%) | 2 (13%) | 1 (6.7%) | 0 (0%) |
| G4a. We discussed the worksheet "Emotions Diary". | 15 (100%) | 3 (20%) | 8 (53%) | 3 (20%) | 1 (6.7%) | 0 (0%) |
| Session 4 | | | | | | |
| G1a. I did a brief review of last week's homework for the house. | 15 (100%) | 6 (40%) | 6 (40%) | 2 (13%) | 1 (6.7%) | 0 (0%) |

| | | | | | | |
|---|---|---|---|---|---|---|
| G1b. I provided psychoeducation about anxiety. | 15 (100%) | 5 (33%) | 7 (47%) | 2 (13%) | 1 (6.7%) | 0 (0%) |
| G2a. I helped the child to understand that the primary goal of this therapy is not to eliminate anxiety but to enhance his skills to manage and cope with it effectively. | 15 (100%) | 4 (27%) | 8 (53%) | 2 (13%) | 1 (6.7%) | 0 (0%) |
| G3a. I helped the child to understand the physical manifestations of anxiety (worksheet "Where I feel anxious"). | 15 (100%) | 5 (33%) | 8 (53%) | 1 (6.7%) | 1 (6.7%) | 0 (0%) |
| G4a. I discussed with the child how instinctive behaviors associated with fighting, fleeing or freezing are triggered. | 14 (93%) | 1 (7.1%) | 10 (71%) | 2 (14%) | 1 (7.1%) | 0 (0%) |
| G5a. I helped the child to understand and apply the concept of externalizing anxiety by separating it from himself. | 15 (100%) | 4 (27%) | 9 (60%) | 2 (13%) | 0 (0%) | 0 (0%) |
| G6a. I described the purpose of creating a collaborative team (clinician, child, family) to collectively combat the child's anxiety. | 15 (100%) | 4 (27%) | 9 (60%) | 1 (6.7%) | 1 (6.7%) | 0 (0%) |
| Session 5 | | | | | | |
| G1a. I discussed with parents the behaviours that inadvertently maintain their child's anxiety (worksheet "Behaviours that maintain anxiety"). | 14 (100%) | 4 (29%) | 4 (29%) | 6 (43%) | 0 (0%) | 0 (0%) |
| G2a. I talked with parents about effective behavioral strategies, such as selective attention and active ignoring ("The power of your attention"). | 14 (100%) | 2 (14%) | 6 (43%) | 5 (36%) | 1 (7.1%) | 0 (0%) |
| G3a. I supported parents in implementing the strategies at home by jointly creating a list of priorities for their child's behaviours. | 14 (100%) | 1 (7.1%) | 8 (57%) | 4 (29%) | 1 (7.1%) | 0 (0%) |
| G4a. I helped parents address potential barriers to implementing the identified behavioral changes at home. | 14 (100%) | 1 (7.1%) | 6 (43%) | 5 (36%) | 2 (14%) | 0 (0%) |
| Session 6 | | | | | | |
| G1a. I did a brief review of last week's homework for the house. | 14 (100%) | 7 (50%) | 5 (36%) | 2 (14%) | 0 (0%) | 0 (0%) |
| G1b. I helped the child to understand the connection between thoughts, feelings and behaviours using the cognitive behavioural triangle (worksheet "Matching game with the GST"). | 14 (100%) | 3 (21%) | 6 (43%) | 5 (36%) | 0 (0%) | 0 (0%) |
| G2a. I discussed with the child that in order to change the course of our thoughts, feelings and behaviors, the first step is to modify our actions. | 14 (100%) | 4 (29%) | 7 (50%) | 2 (14%) | 1 (7.1%) | 0 (0%) |
| G3a. I helped the child to identify and understand behaviours that either reinforce anxiety or, on the contrary, combat it in the long term. | 13 (93%) | 3 (23%) | 8 (62%) | 2 (15%) | 0 (0%) | 0 (0%) |
| G4a. Together with the child, we created a list of behaviours that either enhance anxiety or combat it (worksheet "My behaviours and anxiety"). | 13 (93%) | 2 (15%) | 10 (77%) | 1 (7.7%) | 0 (0%) | 0 (0%) |
| Session 7 | | | | | | |
| G1a. I did a brief review of last week's homework for the house. | 13 (100%) | 5 (38%) | 7 (54%) | 1 (7.7%) | 0 (0%) | 0 (0%) |
| G1b. I showed the child the rationale for the behavioral exposure tasks, (cognitive behavioral triangle worksheet). | 13 (100%) | 3 (23%) | 8 (62%) | 2 (15%) | 0 (0%) | 0 (0%) |

| | | | | | | |
|---|---|---|---|---|---|---|
| G2a. I discussed with the child how consistently practicing actions contrary to what the anxiety dictates, known as exposure, will weaken the power of anxiety and strengthen the child. | 13 (100%) | 2 (15%) | 10 (77%) | 1 (7.7%) | 0 (0%) | 0 (0%) |
| G3a. I helped the child to understand how behaviours can either perpetuate or combat anxiety, (the worksheet "The wave of anxiety"). | 13 (100%) | 2 (15%) | 10 (77%) | 1 (7.7%) | 0 (0%) | 0 (0%) |
| G4a. I helped the child to understand how to assess his anxiety levels (fear thermometer, KYMA-SUDS scale) | 13 (100%) | 3 (23%) | 9 (69%) | 1 (7.7%) | 0 (0%) | 0 (0%) |
| G5a. In collaboration with the child, we created a list of stressful situations, from easy to difficult, in order to plan the strategy of the exposure exercises (worksheet "The fear ladder"). | 13 (100%) | 1 (7.7%) | 5 (38%) | 6 (46%) | 1 (7.7%) | 0 (0%) |
| G6a. I stressed to the child the importance of over-exercising in dealing with stressful situations above and beyond the goals of the report. Report. | 13 (100%) | 2 (15%) | 8 (62%) | 3 (23%) | 0 (0%) | 0 (0%) |
| G7a. I asked the child to work with his parents at home to include additional examples of situations that cause him anxiety, (the "Fear Thermometer" worksheet). | 13 (100%) | 2 (15%) | 8 (62%) | 3 (23%) | 0 (0%) | 0 (0%) |
| Session 8 | | | | | | |
| G1a. I did a brief review of last week's homework for the house. | 13 (100%) | 4 (31%) | 7 (54%) | 2 (15%) | 0 (0%) | 0 (0%) |
| G2a. I helped parents understand the application of the behavioural exposure exercises (worksheets "The wave of anxiety" The ladder of fear"). | 13 (100%) | 3 (23%) | 7 (54%) | 3 (23%) | 0 (0%) | 0 (0%) |
| G3a. We looked again with parents at how they can act as role models and teach bravery to children. | 13 (100%) | 2 (15%) | 7 (54%) | 4 (31%) | 0 (0%) | 0 (0%) |
| G4a. We discussed the reward system that would be appropriate for the family. | 13 (100%) | 2 (15%) | 8 (62%) | 2 (15%) | 1 (7.7%) | 0 (0%) |
| Session 9 | | | | | | |
| G1a. I reviewed the rationale of the report and the hierarchy of fear situations (fear ladder) | 13 (100%) | 2 (15%) | 8 (62%) | 3 (23%) | 0 (0%) | 0 (0%) |
| G2a. We created a fear ladder with the child to work on during the session (worksheet "The fear ladder"). | 13 (100%) | 1 (7.7%) | 9 (69%) | 3 (23%) | 0 (0%) | 0 (0%) |
| G3a. I let the child choose a low-scoring exposure exercise to begin his first in vivo exposure (KYMA -SUDS scale). | 13 (100%) | 2 (15%) | 6 (46%) | 5 (38%) | 0 (0%) | 0 (0%) |
| G4a. We managed any difficulties the child encountered during the exposure exercises (suggested tips from the manual). | 13 (100%) | 2 (15%) | 8 (62%) | 3 (23%) | 0 (0%) | 0 (0%) |
| G4b. Together We designed an exhibition project for the child to do at home (exhibition tracking sheet). | 12 (92%) | 1 (8.3%) | 8 (67%) | 3 (25%) | 0 (0%) | 0 (0%) |
| Session 10 | | | | | | |
| G1a. I did a brief review of last week's homework for the house. | 13 (100%) | 5 (38%) | 7 (54%) | 1 (7.7%) | 0 (0%) | 0 (0%) |
| G2a. I introduced the child to cognitive situational management skills as a way to combat anxiety. | 13 (100%) | 3 (23%) | 8 (62%) | 2 (15%) | 0 (0%) | 0 (0%) |
| G3a. I worked with the child to understand what the thoughts of concern are. | 13 (100%) | 2 (15%) | 7 (54%) | 4 (31%) | 0 (0%) | 0 (0%) |
| G4a. I introduced the child to the concept of thinking traps to prepare him/her for questioning his/her thoughts ("Thinking traps" poster and "Thinking traps" worksheet). | 13 (100%) | 3 (23%) | 6 (46%) | 4 (31%) | 0 (0%) | 0 (0%) |
| G5a. I introduced the child to the idea that stress can cause maladaptive thoughts in children. | 13 (100%) | 3 (23%) | 8 (62%) | 2 (15%) | 0 (0%) | 0 (0%) |

| | | | | | | |
|---|---|---|---|---|---|---|
| G6a. We worked with the child to challenge maladaptive thoughts. | 13 (100%) | 1 (7.7%) | 8 (62%) | 4 (31%) | 0 (0%) | 0 (0%) |
| G7a. We discussed ways in which the child can replace an unhelpful thought with a more realistic one. | 13 (100%) | 2 (15%) | 7 (54%) | 4 (31%) | 0 (0%) | 0 (0%) |
| G8a. I helped the child to become motivated to challenge his anxious thoughts by using supportive statements about himself and adopting examples. | 13 (100%) | 3 (23%) | 7 (54%) | 3 (23%) | 0 (0%) | 0 (0%) |
| G9a. I summarized all of the above under the 4 A's: Recognition, Questioning, Counterattack, Change. | 12 (92%) | 3 (25%) | 6 (50%) | 2 (17%) | 1 (8.3%) | 0 (0%) |
| Session 11 | | | | | | |
| G1a. I discussed with the child if he applied cognitive restructuring what difficulties he encountered. | 13 (100%) | 3 (23%) | 8 (62%) | 2 (15%) | 0 (0%) | 0 (0%) |
| G2a. I reviewed the child's essay exercises. | 13 (100%) | 4 (31%) | 7 (54%) | 1 (7.7%) | 1 (7.7%) | 0 (0%) |
| G3a. We reviewed the hierarchy from session 9 and recalibrated potential exposures to increase the difficulty of the Exposure exercises. | 13 (100%) | 2 (15%) | 8 (62%) | 3 (23%) | 0 (0%) | 0 (0%) |
| G4a. We practiced the report during the session. | 12 (92%) | 3 (25%) | 7 (58%) | 2 (17%) | 0 (0%) | 0 (0%) |
| G5a. We addressed potential difficulties during the report. | 12 (92%) | 2 (17%) | 8 (67%) | 2 (17%) | 0 (0%) | 0 (0%) |
| Session 12 | | | | | | |
| G1a. I did a brief review of last week's homework for the house. | 13 (100%) | 4 (31%) | 8 (62%) | 1 (7.7%) | 0 (0%) | 0 (0%) |
| G2a. I explained to the child the connection between the mind and the body (Coping Cards worksheet). | 13 (100%) | 3 (23%) | 8 (62%) | 2 (15%) | 0 (0%) | 0 (0%) |
| G3a. I had the child practice relaxation skills (deep breathing and progressive muscle relaxation). | 13 (100%) | 3 (23%) | 8 (62%) | 2 (15%) | 0 (0%) | 0 (0%) |
| G4a. There was practice in grounding and mindfulness. | 13 (100%) | 2 (15%) | 10 (77%) | 1 (7.7%) | 0 (0%) | 0 (0%) |
| G5a. I prepared the child for the upcoming end of the treatment and let him express his feelings and thoughts about it. | 13 (100%) | 2 (15%) | 6 (46%) | 5 (38%) | 0 (0%) | 0 (0%) |
| Session 13 | | | | | | |
| G1a. I did a brief review of last week's homework for the house. | 13 (100%) | 4 (31%) | 7 (54%) | 1 (7.7%) | 1 (7.7%) | 0 (0%) |
| G2a. We have encountered difficulties in relation to the aid system. | 12 (92%) | 3 (25%) | 6 (50%) | 1 (8.3%) | 2 (17%) | 0 (0%) |
| G3a. I encouraged parents to adopt a brave lifestyle to act as role models. | 13 (100%) | 3 (23%) | 9 (69%) | 1 (7.7%) | 0 (0%) | 0 (0%) |
| G4a. I prepared the family for the upcoming end of treatment by helping them develop a plan to maintain progress. | 13 (100%) | 1 (7.7%) | 9 (69%) | 2 (15%) | 1 (7.7%) | 0 (0%) |
| G5a. We discussed with parents signs/indications that may indicate that the child's anxiety is at high levels. | 13 (100%) | 2 (15%) | 7 (54%) | 4 (31%) | 0 (0%) | 0 (0%) |
| Session 14 | | | | | | |

| | | | | | | |
|---|---|---|---|---|---|---|
| G1a. I did a quick review of last week'shomework for the house. | 13 (100%) | 5 (38%) | 6 (46%) | 2 (15%) | 0 (0%) | 0 (0%) |
| G2a. I had a detailed discussion with the child about the completion of the treatment, allowing him to express his feelings and thoughts about it. | 13 (100%) | 2 (15%) | 7 (54%) | 3 (23%) | 1 (7.7%) | 0 (0%) |
| G3a. I praised the child's efforts and achievements. | 13 (100%) | 4 (31%) | 8 (62%) | 1 (7.7%) | 0 (0%) | 0 (0%) |
| G4a. I described the next steps in the fear hierarchy for continuing the exposure exercises. | 13 (100%) | 1 (7.7%) | 9 (69%) | 3 (23%) | 0 (0%) | 0 (0%) |
| G5a. I introduced the child to the concept of a brave lifestyle. | 13 (100%) | 5 (38%) | 6 (46%) | 2 (15%) | 0 (0%) | 0 (0%) |
| G6a. I helped the child to recognise his/her own signs that may signal the need for further support (worksheet "Warning bells"). | 13 (100%) | 2 (15%) | 9 (69%) | 2 (15%) | 0 (0%) | 0 (0%) |
| G7a. I gave the child his graduation certificate and praised his effort throughout the treatment process. | 13 (100%) | 8 (62%) | 4 (31%) | 1 (7.7%) | 0 (0%) | 0 (0%) |

Supplementary Table 9: Depression treatment protocol: session-level item completion and difficulty (trainee)

| DEPRESSION TREATMENT PROTOCOL: PILOT 1 | Item completion n (%) | Very easy | Easy | Neutral | Difficult | Very difficult y |
|---|---|---|---|---|---|---|
| Session 1 | | | | | | |
| G1. I welcomed the teen to treatment and set a mutual agenda | 4 (100%) | 2 (50%) | 2 (50%) | 0 (0%) | 0 (0%) | 0 (0%) |
| G2a. I explained the depressive symptoms and how they manifest in teens (psychoeducation) | 4 (100%) | 1 (25%) | 3 (75%) | 0 (0%) | 0 (0%) | 0 (0%) |
| G2b. I explained the basic principles of CBT and what will be covered during the sessions. | 4 (100%) | 1 (25%) | 2 (50%) | 1 (25%) | 0 (0%) | 0 (0%) |
| G3. I engaged the teen in discussion about their goals for treatment and they completed the Goals Worksheet | 4 (100%) | 1 (25%) | 1 (25%) | 2 (50%) | 0 (0%) | 0 (0%) |
| Session 2 | | | | | | |
| G1. I provided a brief review of last week's home assignment and encouraged the teen to collaboratively address any incomplete aspects during today's session (i.e., CBT Triangle Worksheet) | 4 (100%) | 1 (25%) | 2 (50%) | 0 (0%) | 1 (25%) | 0 (0%) |
| G2a. I explained how emotions are linked to bodily sensations, distinguishing between comfortable and uncomfortable feelings. | 4 (100%) | 0 (0%) | 2 (50%) | 1 (25%) | 1 (25%) | 0 (0%) |
| G2b. I helped the teen label and reflect on emotions using the Emotions List worksheet | 4 (100%) | 0 (0%) | 3 (75%) | 0 (0%) | 1 (25%) | 0 (0%) |
| G3. I introduced and taught the teen about the benefits of tracking their mood, emotions, and behaviors. | 4 (100%) | 0 (0%) | 2 (67%) | 1 (33%) | 0 (0%) | 0 (0%) |
| Session 3 | | | | | | |
| G1a. I explained to parents how the symptoms of depression may appear in teens (psychoeducation) | 4 (100%) | 0 (0%) | 2 (50%) | 2 (50%) | 0 (0%) | 0 (0%) |
| G1b. I explained the basic principles of CBT and what will be covered during the sessions. | 4 (100%) | 0 (0%) | 3 (75%) | 0 (0%) | 1 (25%) | 0 (0%) |
| G2. I instructed the parent on techniques for validating their child and facilitated a role-playing exercise to assist them in practicing validation skills during our session. | 4 (100%) | 0 (0%) | 2 (50%) | 1 (25%) | 1 (25%) | 0 (0%) |
| G3. I guided the parent in learning and practicing mindfulness skills. | 4 (100%) | 0 (0%) | 0 (0%) | 1 (33%) | 2 (67%) | 0 (0%) |
| G4. I engaged the parents in a discussion about their goals for treatment using the Parent Goals Worksheet | 4 (100%) | 0 (0%) | 1 (33%) | 1 (33%) | 1 (33%) | 0 (0%) |
| Session 4 | | | | | | |
| G1. I provided a brief review of last week's home assignment and encouraged the teen to collaboratively address any incomplete aspects during today's session (i.e., Mood Tracker Worksheet) | 4 (100%) | 1 (25%) | 1 (25%) | 1 (25%) | 1 (25%) | 0 (0%) |
| G2a. I acquainted the teen with the concept of engaging in mood-boosting activities as a means of enhancing their mood. | 4 (100%) | 1 (25%) | 3 (75%) | 0 (0%) | 0 (0%) | 0 (0%) |
| G2b. I helped the teen identify and select mood boosters using the Mood Boosting Activities List | 4 (100%) | 2 (50%) | 2 (50%) | 0 (0%) | 0 (0%) | 0 (0%) |

| | | | | | | |
|---|---|---|---|---|---|---|
| G3. I introduced the teenager to the significance of taking care of our bodies to enhance our mood, and I discussed methods to achieve this. | 4 (100%) | 2 (67%) | 1 (33%) | 0 (0%) | 0 (0%) | 0 (0%) |
| Session 5 | | | | | | |
| G1. I provided a brief review of last week's home assignment and encouraged the teen to collaboratively address any incomplete aspects during today's session (i.e., Mood Tracker /Mood Booster Worksheet) | 4 (100%) | 0 (0%) | 2 (50%) | 1 (25%) | 1 (25%) | 0 (0%) |
| G2. I introduced the teen to the concept of (negative) automatic thoughts and their impact on mood | 4 (100%) | 0 (0%) | 1 (25%) | 1 (25%) | 2 (50%) | 0 (0%) |
| G3. I introduced the teen to the concept of (negative) core beliefs and their impact on mood | 4 (100%) | 0 (0%) | 0 (0%) | 1 (33%) | 2 (67%) | 0 (0%) |
| G4. I explained the interconnection between thoughts, emotions, and behaviors, illustrating how these components can converge into downward negative spirals. | 4 (100%) | 0 (0%) | 0 (0%) | 1 (33%) | 2 (67%) | 0 (0%) |
| Session 6 | | | | | | |
| G1. I provided a brief review of last week's home assignment and encouraged the teen to collaboratively address any incomplete aspects during today's session (i.e., Mood Tracker/ Mood Booster Worksheet and Automatic Thoughts Worksheet) | 4 (100%) | 0 (0%) | 3 (75%) | 0 (0%) | 1 (25%) | 0 (0%) |
| G2. I helped the teen understand how thinking traps can affect our feelings | 4 (100%) | 0 (0%) | 2 (50%) | 1 (25%) | 1 (25%) | 0 (0%) |
| G3. I assisted the teen in developing awareness and identifying their thinking traps using the Thinking Traps worksheet. | 4 (100%) | 0 (0%) | 1 (33%) | 2 (67%) | 0 (0%) | 0 (0%) |
| Session 7 | | | | | | |
| G1. I provided a brief review of last week's home assignment and encouraged the teen to collaboratively address any incomplete aspects during today's session (i.e., Mood Tracker/Mood Booster Worksheet, Balanced Thinking Chart) | 4 (100%) | 0 (0%) | 3 (75%) | 1 (25%) | 0 (0%) | 0 (0%) |
| G2. I introduced mindful awareness to the teenager, showing how gaining insight into their moment-to-moment emotions empowers intentional choices in their responses. | 4 (100%) | 0 (0%) | 2 (50%) | 2 (50%) | 0 (0%) | 0 (0%) |
| G3. I introduced the Emotion Wave tool to help the teen apply awareness skills, make mood-enhancing choices using the Emotion Wave Worksheet. | 4 (100%) | 0 (0%) | 1 (33%) | 2 (67%) | 0 (0%) | 0 (0%) |
| Session 8 | | | | | | |
| G1. I provided a brief review of last week's home assignment and encouraged the teen to collaboratively address any incomplete aspects during today's session (i.e., Mood Tracker/ Mood Booster Worksheet and Emotion Wave) | 4 (100%) | 0 (0%) | 2 (50%) | 2 (50%) | 0 (0%) | 0 (0%) |
| G2. I guided the teen in recognizing intense emotions and gathering details about their experiences during these moments | 4 (100%) | 0 (0%) | 2 (50%) | 2 (50%) | 0 (0%) | 0 (0%) |
| G3a. I addressed techniques for effectively managing intense emotions utilizing the BEST method. | 4 (100%) | 0 (0%) | 2 (67%) | 1 (33%) | 0 (0%) | 0 (0%) |
| G3b. I addressed techniques for effectively managing intense emotions, including embracing reality and employing sensory soothing. | 4 (100%) | 0 (0%) | 0 (0%) | 3 (100%) | 0 (0%) | 0 (0%) |
| Session 9 | | | | | | |

| | | | | | | |
|---|---|---|---|---|---|---|
| G1. I provided a brief review of last week's home assignment and encouraged the teen to collaboratively address any incomplete aspects during today's session (i.e., Mood Tracker/ Mood Booster Worksheet and and the Self-soothe Kit) | 4 (100%) | 0 (0%) | 4 (100%) | 0 (0%) | 0 (0%) | 0 (0%) |
| G2. I assisted the teenager in understanding barriers to healthy relationships during periods of depression. | 4 (100%) | 0 (0%) | 4 (100%) | 0 (0%) | 0 (0%) | 0 (0%) |
| G3. I helped the teenager identify relationship goals and the behaviors needed to achieve them using the Relationship Goals Worksheet | 4 (100%) | 0 (0%) | 2 (67%) | 1 (33%) | 0 (0%) | 0 (0%) |
| G4. I guided the teenager in choosing mood-boosting social activities using the Social Activities Checklist and discussed challenges in relationships and social interactions. | 4 (100%) | 0 (0%) | 3 (100%) | 0 (0%) | 0 (0%) | 0 (0%) |
| Session 10 | | | | | | |
| G1. I evaluated progress compared to the initial parent session goals, briefly reviewing the skill of validation in communicating with the teen, parent skills practice, and mindfulness skills. | 4 (100%) | 0 (0%) | 3 (75%) | 1 (25%) | 0 (0%) | 0 (0%) |
| G2. I reviewed with the parent the four styles of communication, encouraging them to reflect upon and discuss which one aligns most closely with their own approach. | 4 (100%) | 0 (0%) | 2 (50%) | 2 (50%) | 0 (0%) | 0 (0%) |
| G3a. I asked the parent to identify communication traps with their teenager and explored alternative responses using the Communication Traps Checklist. | 4 (100%) | 1 (25%) | 3 (75%) | 0 (0%) | 0 (0%) | 0 (0%) |
| G3b. I covered a variety of problem-solving strategies and techniques for identifying the most practical and achievable solutions using the Effective Communication Worksheet. | 4 (100%) | 0 (0%) | 3 (100%) | 0 (0%) | 0 (0%) | 0 (0%) |
| G4. I explained the importance of self-validation in eventually communicating effectively with their teen and provided guidance on how to practice it. | 4 (100%) | 0 (0%) | 2 (50%) | 2 (50%) | 0 (0%) | 0 (0%) |
| Session 11 | | | | | | |
| G1. I provided a brief review of last week's home assignment and encouraged the teen to collaboratively address any incomplete aspects during today's session (i.e., Mood Tracker/ Mood Booster Worksheet, the Relationship Goals and Social Activities Checklist worksheets) | 4 (100%) | 0 (0%) | 4 (100%) | 0 (0%) | 0 (0%) | 0 (0%) |
| G2. I reviewed the four styles of communication with the teenager, encouraging them to reflect upon and discuss which one aligns most closely with their own approach. | 4 (100%) | 0 (0%) | 3 (75%) | 1 (25%) | 0 (0%) | 0 (0%) |
| G3a. I assisted the teenager in recognizing barriers to expressing emotions and discussed assertive communication styles using the Communication Record Worksheet. | 4 (100%) | 0 (0%) | 1 (33%) | 1 (33%) | 1 (33%) | 0 (0%) |
| G3b. I introduced the concept of "I feel" statements as a means to practice assertive communication and assigned the I Feel Statements Worksheet for homework. | 4 (100%) | 0 (0%) | 1 (33%) | 2 (67%) | 0 (0%) | 0 (0%) |
| Session 12 | | | | | | |
| G1. I provided a brief review of last week's home assignment and encouraged the teen to collaboratively address any incomplete aspects during today's session (i.e., Mood Tracker/Mood Booster Worksheet, the Communication Record Home Assignment, and the I Feel Statements Home Assignment) | 4 (100%) | 0 (0%) | 2 (50%) | 2 (50%) | 0 (0%) | 0 (0%) |
| G2. I provided psychoeducation on effective problem-solving, followed by a discussion to assess the teen's problem-solving approach, strategies, and motivation for their current style. | 4 (100%) | 0 (0%) | 2 (50%) | 2 (50%) | 0 (0%) | 0 (0%) |

| | | | | | | |
|---|---|---|---|---|---|---|
| G3. We went over practical problem-solving steps, talked about possible challenges, and went through an example using the Problem Solving Worksheet. | 4 (100%) | 0 (0%) | 3 (100%) | 0 (0%) | 0 (0%) | 0 (0%) |
| Session 13 | | | | | | |
| G1. We revisited previously taught skills, talked about their usefulness with examples, and practiced using them (e.g., through problem-solving and role-playing) | 4 (100%) | 1 (25%) | 2 (50%) | 1 (25%) | 0 (0%) | 0 (0%) |
| G2a. I instructed the parent on understanding the impact of their behavior on their child's behavior, emphasizing the basics of behaviorism. | 4 (100%) | 0 (0%) | 1 (25%) | 2 (50%) | 1 (25%) | 0 (0%) |
| G2b. We discussed the three crucial components of behavior change: awareness, motivation, and alternative behaviors using the Teen Behavior Checklist. | 4 (100%) | 0 (0%) | 3 (75%) | 1 (25%) | 0 (0%) | 0 (0%) |
| G3. We talked about treatment progress and the parental role in the maintenance phase, covering skill practice, consistent reinforcement, observing their child's behavior, and planning for challenging conversations using the Skills Quick Reference. | 4 (100%) | 0 (0%) | 3 (100%) | 0 (0%) | 0 (0%) | 0 (0%) |
| Session 14 | | | | | | |
| G1. I provided a brief review of last week's home assignment and encouraged the teen to collaboratively address any incomplete aspects during today's session (i.e., Mood Tracker/Mood Booster Worksheet, and the Self-Compassion Journal) | 4 (100%) | 1 (25%) | 1 (25%) | 2 (50%) | 0 (0%) | 0 (0%) |
| G2a. I prepared the teenager for the end of treatment, reviewed their goals and progress using the Skills Quick Reference worksheet. | 4 (100%) | 0 (0%) | 3 (75%) | 1 (25%) | 0 (0%) | 0 (0%) |
| G2b. I talked to the teenager about their feelings on ending treatment and discussed effective ways to prepare for potential challenges in the maintenance phase using the Red Flags and Planning Ahead worksheet. | 4 (100%) | 0 (0%) | 2 (50%) | 2 (50%) | 0 (0%) | 0 (0%) |
| G3. I engaged the teen in a rehearsal activity to prepare for mood changes, following the steps for rehearsing using the Rehearsing Worksheet. | 4 (100%) | 0 (0%) | 1 (25%) | 3 (75%) | 0 (0%) | 0 (0%) |
| G4. I checked in with the teenager regarding their treatment experience and talked about available resources for them post-treatment, including crisis support and ongoing treatment options. | 4 (100%) | 0 (0%) | 3 (100%) | 0 (0%) | 0 (0%) | 0 (0%) |

| **DEPRESSION TREATMENT PROTOCOL: PILOT 2** | **Item completion n (%)** | **Very easy** | **Easy** | **Neutral** | **Difficult** | **Very difficult y** |
|---|---|---|---|---|---|---|
| Session 1 | | | | | | |
| G1a. I welcomed the teenager to therapy and set a common agenda | 12 (100%) | 3 (25%) | 8 (67%) | 1 (8.3%) | 0 (0%) | 0 (0%) |
| G2a. I explained depressive symptoms and how they manifest in adolescents (psychoeducation) | 12 (100%) | 2 (17%) | 10 (83%) | 0 (0%) | 0 (0%) | 0 (0%) |
| G2b. I explained the basic principles of GST and what we will cover during the sessions | 12 (100%) | 2 (17%) | 7 (58%) | 2 (17%) | 1 (8.3%) | 0 (0%) |
| G3a.I discussed with the adolescent about his goals for his treatment and the adolescent completed the "Goals" worksheet | 12 (100%) | 1 (8.3%) | 6 (50%) | 3 (25%) | 2 (17%) | 0 (0%) |

| | | | | | | |
|---|---|---|---|---|---|---|
| Session 2 | | | | | | |
| G1a. I reviewed the homework for the previous week. | 13 (100%) | 2 (15%) | 8 (62%) | 3 (23%) | 0 (0%) | 0 (0%) |
| G2a. I explained how emotions are linked to the physical sensations, distinguishing between pleasant and unpleasant emotions. | 13 (100%) | 2 (15%) | 8 (62%) | 3 (23%) | 0 (0%) | 0 (0%) |
| G2b. I helped the teenager to name the feelings and reflect on them using the worksheet "Identifying feelings" | 13 (100%) | 2 (15%) | 7 (54%) | 3 (23%) | 1 (7.7%) | 0 (0%) |
| G3a.I explained to the teenager the benefits of monitoring his mood, emotions and behaviour. | 13 (100%) | 1 (7.7%) | 8 (62%) | 3 (23%) | 1 (7.7%) | 0 (0%) |
| Session 3 | | | | | | |
| G1a. I explained to parents how depression symptoms can appear in adolescents (psychoeducation). | 12 (92%) | 1 (8.3%) | 8 (67%) | 3 (25%) | 0 (0%) | 0 (0%) |
| G1b. I explained the basic principles of GST and what we will cover during the sessions. | 12 (92%) | 2 (17%) | 6 (50%) | 4 (33%) | 0 (0%) | 0 (0%) |
| G2a. I guided parents in the application of validation techniques | 12 (92%) | 1 (8.3%) | 6 (50%) | 4 (33%) | 1 (8.3%) | 0 (0%) |
| G3a.I discussed with parents about their goals for treatment using the Parent Goals Worksheet. | 12 (92%) | 1 (8.3%) | 5 (42%) | 4 (33%) | 2 (17%) | 0 (0%) |
| Session 4 | | | | | | |
| G1a. I reviewed the homework for the previous week. | 13 (100%) | 3 (23%) | 7 (54%) | 2 (15%) | 1 (7.7%) | 0 (0%) |
| G2a. I familiarized the adolescent with the concept of participating in mood-enhancing activities as a means of improving his mood. | 13 (100%) | 1 (7.7%) | 8 (62%) | 4 (31%) | 0 (0%) | 0 (0%) |
| G2b. I helped the teenager to identify and choose anything that would improve his mood, using the mood-enhancing activity list. | 13 (100%) | 1 (7.7%) | 8 (62%) | 4 (31%) | 0 (0%) | 0 (0%) |
| G3a. I discussed methods on body care to enhance mood, | 13 (100%) | 2 (15%) | 7 (54%) | 4 (31%) | 0 (0%) | 0 (0%) |
| Session 5 | | | | | | |
| G1a. I reviewed the homework for the previous week. | 13 (100%) | 3 (23%) | 7 (54%) | 3 (23%) | 0 (0%) | 0 (0%) |
| G2a. I introduced the teenager to the concept of (negative) automatic thoughts and their effect on mood | 13 (100%) | 2 (15%) | 7 (54%) | 4 (31%) | 0 (0%) | 0 (0%) |
| G3a. I introduced the adolescent to the concept of (negative) nuclear beliefs and their effect on mood. | 13 (100%) | 2 (15%) | 7 (54%) | 4 (31%) | 0 (0%) | 0 (0%) |
| G4a. I explained the connection between thoughts, feelings and behaviours. | 13 (100%) | 2 (15%) | 8 (62%) | 2 (15%) | 1 (7.7%) | 0 (0%) |
| Session 6 | | | | | | |
| G1a. I reviewed the homework for the previous week. | 13 (100%) | 4 (31%) | 6 (46%) | 3 (23%) | 0 (0%) | 0 (0%) |
| G2a. I helped the teenager understand how the pitfalls of thinking can affect our emotions. | 13 (100%) | 2 (15%) | 6 (46%) | 4 (31%) | 1 (7.7%) | 0 (0%) |
| G3a. I helped the teen identify his thinking traps using the "Thinking Traps" worksheet. | 13 (100%) | 2 (15%) | 6 (46%) | 5 (38%) | 0 (0%) | 0 (0%) |
| Session 7 | | | | | | |
| G1a. I reviewed the homework for the previous week. | 13 (100%) | 4 (31%) | 5 (38%) | 4 (31%) | 0 (0%) | 0 (0%) |
| G2a. I introduced how questioning thoughts can affect emotions. | 13 (100%) | 2 (15%) | 7 (54%) | 4 (31%) | 0 (0%) | 0 (0%) |

| | | | | | | |
|---|---|---|---|---|---|---|
| G2a. I helped the teenager learn how to challenge unhelpful thoughts and think more helpful thoughts. | 13 (100%) | 2 (15%) | 5 (38%) | 4 (31%) | 2 (15%) | 0 (0%) |
| Session 8 | | | | | | |
| G1a. I reviewed the homework for the previous week. | 13 (100%) | 4 (31%) | 6 (46%) | 2 (15%) | 1 (7.7%) | 0 (0%) |
| G2a. I guided the teenager in recognizing intense emotions. | 13 (100%) | 3 (23%) | 6 (46%) | 2 (15%) | 2 (15%) | 0 (0%) |
| G3a. I was involved in techniques for effectively managing intense emotions using the BEST skill. | 12 (92%) | 2 (17%) | 6 (50%) | 3 (25%) | 1 (8.3%) | 0 (0%) |
| G3b. I dealt with techniques for the effective management of intense emotions (e.g. acceptance of reality) | 13 (100%) | 2 (15%) | 7 (54%) | 3 (23%) | 1 (7.7%) | 0 (0%) |
| Session 9 | | | | | | |
| G1a. I reviewed the homework for the previous week. | 13 (100%) | 2 (15%) | 6 (46%) | 5 (38%) | 0 (0%) | 0 (0%) |
| G2a. I helped the teenager to identify his goals for his relationships (worksheet "Goals for relationships") | 13 (100%) | 2 (15%) | 7 (54%) | 4 (31%) | 0 (0%) | 0 (0%) |
| G3a. I helped the teenager to identify the behaviours to achieve the goals he set for his relationships (worksheet "Goals for relationships") | 13 (100%) | 3 (23%) | 6 (46%) | 4 (31%) | 0 (0%) | 0 (0%) |
| G4a. I guided the adolescent in choosing social activities that enhance mood ("Social activity list"). | 13 (100%) | 3 (23%) | 6 (46%) | 4 (31%) | 0 (0%) | 0 (0%) |
| Session 10 | | | | | | |
| G1a. I reviewed the homework for the previous week. | 13 (100%) | 3 (23%) | 6 (46%) | 4 (31%) | 0 (0%) | 0 (0%) |
| G2a. I reviewed the four communication styles with the parent, encouraging them to think about and discuss which one best fits their approach. | 13 (100%) | 2 (15%) | 5 (38%) | 5 (38%) | 1 (7.7%) | 0 (0%) |
| G3a. I asked the parent to explore alternative responses to the communication pitfalls with the adolescent ("Communication pitfall list"). | 13 (100%) | 1 (7.7%) | 8 (62%) | 3 (23%) | 1 (7.7%) | 0 (0%) |
| G3b. I asked the parent to explore alternative responses to the communication pitfalls with the adolescent ("Communication pitfall list"). | 13 (100%) | 1 (7.7%) | 9 (69%) | 3 (23%) | 0 (0%) | 0 (0%) |
| G4a. I explained the importance of self-affirmation for effective communication with the adolescent and provided instructions on how to implement it. | 13 (100%) | 1 (7.7%) | 7 (54%) | 4 (31%) | 1 (7.7%) | 0 (0%) |
| Session 11 | | | | | | |
| G1a. I reviewed the homework for the previous week. | 13 (100%) | 3 (23%) | 8 (62%) | 2 (15%) | 0 (0%) | 0 (0%) |
| G2a. I reviewed the four communication styles with the teenager, encouraging them to think about and discuss which one best fits their approach. | 13 (100%) | 3 (23%) | 7 (54%) | 3 (23%) | 0 (0%) | 0 (0%) |
| G3a. I helped the adolescent identify barriers to expressing feelings (Communication Recording worksheet). | 13 (100%) | 3 (23%) | 7 (54%) | 3 (23%) | 0 (0%) | 0 (0%) |
| G3b. I introduced the concept of "I feel" statements as a means of practicing assertive communication. | 13 (100%) | 3 (23%) | 7 (54%) | 3 (23%) | 0 (0%) | 0 (0%) |
| Session 12 | | | | | | |
| G1a. I reviewed the homework for the previous week. | 13 (100%) | 4 (31%) | 7 (54%) | 2 (15%) | 0 (0%) | 0 (0%) |
| G2a. I provided psychoeducation on effective problem solving. | 13 (100%) | 3 (23%) | 8 (62%) | 2 (15%) | 0 (0%) | 0 (0%) |
| G3a. We looked at the practical steps of problem solving (worksheet "Problem solving"). | 13 (100%) | 3 (23%) | 6 (46%) | 3 (23%) | 1 (7.7%) | 0 (0%) |

| | | | | | | |
|---|---|---|---|---|---|---|
| Session 13 | | | | | | |
| G1a. I reviewed the homework for the previous week. | 12 (92%) | 3 (25%) | 5 (42%) | 3 (25%) | 1 (8.3%) | 0 (0%) |
| G2a. I guided the parent in understanding the effect of their behaviour on the child's behaviour. | 12 (92%) | 3 (25%) | 2 (17%) | 5 (42%) | 2 (17%) | 0 (0%) |
| G2b. We discussed the three key components of behaviour change: awareness, motivation and alternative behaviours ("Adolescent Behaviour List") | 12 (92%) | 3 (25%) | 5 (42%) | 3 (25%) | 1 (8.3%) | 0 (0%) |
| G3a. We talked about the progress of treatment and the role of parents in the maintenance phase ("Brief review of skills"). | 12 (92%) | 3 (25%) | 5 (42%) | 3 (25%) | 1 (8.3%) | 0 (0%) |
| Session 14 | | | | | | |
| G1a. I reviewed the homework for the previous week. | 13 (100%) | 4 (31%) | 6 (46%) | 3 (23%) | 0 (0%) | 0 (0%) |
| G2a. I reviewed the adolescent's goals and progress (skills brief review worksheet). | 13 (100%) | 3 (23%) | 6 (46%) | 4 (31%) | 0 (0%) | 0 (0%) |
| G2b. I discussed with the teenager effective ways to prepare for possible challenges in the retention phase (worksheet "Warning bells and planning for the future"). | 13 (100%) | 3 (23%) | 7 (54%) | 3 (23%) | 0 (0%) | 0 (0%) |
| G3a. I helped the teenager to prepare for possible future mood changes (worksheet "Rehearsal: Planning for difficult situations"). | 13 (100%) | 3 (23%) | 5 (38%) | 5 (38%) | 0 (0%) | 0 (0%) |
| G4a. I checked with the adolescent about his treatment experience and talked about the resources available to him after treatment. | 13 (100%) | 3 (23%) | 6 (46%) | 4 (31%) | 0 (0%) | 0 (0%) |

Supplementary Table 10: BPT treatment protocol: session-level item completion and difficulty (trainee)

| BPT TREATMENT PROTOCOL: PILOT 1 | Item completion n (%) | Very easy | Easy | Neutral | Difficult | Very difficult y |
|---|---|---|---|---|---|---|
| Session 1 | | | | | | |
| G1. I initiated introductions with parents, employing conversational inquiries to foster a connection. I reflected on their experiences and provided validation where appropriate, demonstrating positive regard. | 7 (100%) | 0 (0%) | 6 (86%) | 1 (14%) | 0 (0%) | 0 (0%) |
| G2. I explained the scope of this treatment (i.e., including duration, frequency, family participation, and overall goals for interventions) utilizing the “Outline of Therapy” and “Recipe for Improving Behavior” worksheets | 7 (100%) | 1 (14%) | 4 (57%) | 2 (29%) | 0 (0%) | 0 (0%) |
| G3. I encouraged parents' reflection and sharing with curiosity and empathy, refraining from actively trying to correct or alter their expressed thoughts/feelings or from giving any judgment-laden feedback. | 7 (100%) | 1 (14%) | 2 (29%) | 4 (57%) | 0 (0%) | 0 (0%) |
| G4. I engaged in discussions and posed questions to parents related to various aspects of parenthood, including their core values, using the "Knowing Ourselves Session 1" worksheet. | 7 (100%) | 0 (0%) | 3 (43%) | 3 (43%) | 1 (14%) | 0 (0%) |
| G5a. I introduced parents to behavioral principles regarding reinforcement, praise, positive attention, and differential attention utilizing “The Power of Praise and Positive Attention” worksheet | 6 (86%) | 0 (0%) | 2 (33%) | 3 (50%) | 1 (17%) | 0 (0%) |
| G5b. I collaborated with parents to formulate a plan for initiating praise of appropriate behaviors and reducing negative attention for inappropriate behaviors at home in the upcoming week, utilizing the "Recipe – Ingredient 1" and "Home Practice – Week 1" worksheets. | 6 (86%) | 0 (0%) | 4 (67%) | 0 (0%) | 2 (33%) | 0 (0%) |
| G6. I praised parents for their efforts during the session, reinforcing the positive steps taken to support their child's development, and I concluded by inviting their feedback | 7 (100%) | 0 (0%) | 2 (29%) | 4 (57%) | 1 (14%) | 0 (0%) |
| Session 2 | | | | | | |
| G1. I welcomed the parents upon their return to therapy, providing a brief summary of today's session structure and goals, as outlined on the first page of the manual for this session | 7 (100%) | 3 (43%) | 4 (57%) | 0 (0%) | 0 (0%) | 0 (0%) |
| G2. I reviewed with the parents the home assignment from last week, praising their efforts, and together, we collaboratively brainstormed solutions to address any challenges in implementing new skills (i.e., use of praise, positive attention) | 7 (100%) | 1 (14%) | 5 (71%) | 1 (14%) | 0 (0%) | 0 (0%) |
| G3. I encouraged parents' reflection and sharing with curiosity and empathy, refraining from actively trying to correct or alter their expressed thoughts/feelings or from giving any judgment-laden feedback, utilizing the “Knowing Ourselves Session 2” worksheet | 6 (86%) | 0 (0%) | 4 (67%) | 2 (33%) | 0 (0%) | 0 (0%) |
| G4. I discussed with the parents how thoughts, feelings, and behaviors are interconnected and mutually influence each other, explaining how these dynamics impact their interactions with their child utilizing the “Cognitive Triad” worksheet | 7 (100%) | 0 (0%) | 7 (100%) | 0 (0%) | 0 (0%) | 0 (0%) |

| | | | | | | |
|---|---|---|---|---|---|---|
| G5. I worked together with the parents to solidify the plan for sustaining the home practice of praising and providing positive attention for appropriate behaviors in the coming week, utilizing the "Recipe – Ingredient 2" and "Home Practice Goals – Week 2" worksheets. | 7 (100%) | 0 (0%) | 5 (71%) | 2 (29%) | 0 (0%) | 0 (0%) |
| G6. I praised parents for their efforts in the session, forewarned them about potential early discouragement, emphasized the gradual process of change, and provided assurance that the long-term commitment would ultimately benefit their child. | 7 (100%) | 0 (0%) | 5 (71%) | 1 (14%) | 1 (14%) | 0 (0%) |
| Session 3 | | | | | | |
| G1. I welcomed the parents upon their return to therapy, providing a brief summary of today's session structure and goals, as outlined on the first page of the manual for this session | 7 (100%) | 1 (14%) | 5 (71%) | 1 (14%) | 0 (0%) | 0 (0%) |
| G2. I reviewed with the parents the home assignment from last week, praising their efforts, and together, we collaboratively brainstormed solutions to address any challenges in implementing new skills (i.e., continued use of praise and positive attention, practicing self-awareness of thoughts/feelings/behaviors) | 7 (100%) | 0 (0%) | 6 (86%) | 0 (0%) | 1 (14%) | 0 (0%) |
| G3. I engaged in a discussion with the parents about different parenting styles, sharing insights from research on the effectiveness of the Authoritative parenting style, using the "Parenting Styles" worksheet. | 7 (100%) | 0 (0%) | 6 (86%) | 1 (14%) | 0 (0%) | 0 (0%) |
| G4. I engaged in a conversation with the parents regarding the pivotal role they play in modeling behaviors for their children, utilizing the "Recipe – Ingredient 3" Worksheet. | 7 (100%) | 0 (0%) | 6 (86%) | 0 (0%) | 1 (14%) | 0 (0%) |
| G5. I collaborated with the parents to create a plan for modeling and teaching desired behaviors at home, and we reinforced the strategy for maintaining the home practice of praising and offering positive attention for appropriate behaviors in the upcoming week, utilizing the "Model / Teach" and "Home Practice Goals – Week 3" worksheets. | 7 (100%) | 0 (0%) | 6 (86%) | 0 (0%) | 1 (14%) | 0 (0%) |
| Session 4 | | | | | | |
| G1. I reviewed with the parents the home assignment from the previous week, praising their efforts, and we jointly brainstormed solutions to tackle challenges in implementing new skills (i.e., sustained use of praise and positive attention, cultivating self-awareness, and employing modeling/teaching techniques for their child) | 7 (100%) | 0 (0%) | 6 (86%) | 1 (14%) | 0 (0%) | 0 (0%) |
| G2. I engaged in a discussion with the parents regarding the typical functions of behavior and potential underlying causes and contributors to various behaviors in children, utilizing the "Understand Behaviors" worksheet. | 7 (100%) | 0 (0%) | 4 (57%) | 3 (43%) | 0 (0%) | 0 (0%) |
| G3. I discussed with the parents the benefits of positive behavior plans in fostering behavior change, utilizing the "Recipe - Ingredient 4" worksheet. | 7 (100%) | 0 (0%) | 2 (29%) | 5 (71%) | 0 (0%) | 0 (0%) |
| G4. I worked with the parents to create a written positive behavior plan for home practice, concentrating on a specific behavior identified for change during this session. | 7 (100%) | 0 (0%) | 2 (29%) | 3 (43%) | 2 (29%) | 0 (0%) |
| Session 5 | | | | | | |

| | | | | | | |
|---|---|---|---|---|---|---|
| G1. I welcomed the child in therapy, offered an age-appropriate explanation of my role and our work with the family, outlined the plan for the day, and took the opportunity to learn more about them while modeling praise and positive attention to the parents during our discussion. | 7 (100%) | 2 (29%) | 5 (71%) | 0 (0%) | 0 (0%) | 0 (0%) |
| G2a. I provided the parents and child with an opportunity to freely engage in play together, aiming to gather insights into parent-child relationships and their interactive styles, utilizing the "Observing Parent-Child Playtime" worksheet. | 7 (100%) | 1 (14%) | 4 (57%) | 1 (14%) | 1 (14%) | 0 (0%) |
| G2b. I offered friendly and authentic positive comments about the family's play, and I participated in a brief play session with the child, allowing the child to lead, while making positive observations. | 7 (100%) | 0 (0%) | 5 (71%) | 0 (0%) | 2 (29%) | 0 (0%) |
| G3a. I conveyed enthusiasm for any rewards the child has earned at home while implementing the positive behavioral plan, recognizing the child's efforts to achieve behavior goals and acknowledging the parents for observing and appreciating the child's accomplishments. | 7 (100%) | 0 (0%) | 5 (71%) | 1 (14%) | 1 (14%) | 0 (0%) |
| G3b. I collaborated with the family to address any obstacles in implementation at home, addressing specific concerns privately with parents, as needed, to avoid negative discussions in front of the child. I encouraged both parents and child to persist in their efforts with the positive behavior plan in the coming week. | 7 (100%) | 0 (0%) | 6 (86%) | 0 (0%) | 1 (14%) | 0 (0%) |
| Session 6 | | | | | | |
| G1. I checked how the behavioral plan progressed. If necessary, adjustments were made to the behavioral plan. | 7 (100%) | 0 (0%) | 6 (86%) | 1 (14%) | 0 (0%) | 0 (0%) |
| G2a. I explained the importance of regular, enjoyable quality time with their child, emphasizing its role in teaching skills that foster positive parent-child interactions using the Recipe – Ingredient 5 worksheet. | 7 (100%) | 0 (0%) | 5 (71%) | 2 (29%) | 0 (0%) | 0 (0%) |
| G2b. I prepared the parents to engage in consistent quality time with their child at home using the Creating Nourishing Relationship Time worksheet. | 7 (100%) | 0 (0%) | 4 (57%) | 2 (29%) | 1 (14%) | 0 (0%) |
| G3. I began guiding parents in addressing their and their children's emotional needs and developing emotion regulation skills referring to the Recipe – Ingredient 6 worksheet. | 7 (100%) | 0 (0%) | 6 (86%) | 1 (14%) | 0 (0%) | 0 (0%) |
| G4. I urged parents to continue implementing the positive behavioral plan and referred the parents to the Home Practice Goals – Week 6 worksheet to track how often each parent applied the quality time and to write down any reactions. | 7 (100%) | 0 (0%) | 5 (71%) | 1 (14%) | 1 (14%) | 0 (0%) |
| Session 7 | | | | | | |
| G1a. During a playtime session at the office, I used the Nourishing Skills Checklist and Playtime Notes worksheets to observe how parents implemented positive interaction skills while noting the child’s behavior. | 7 (100%) | 0 (0%) | 6 (86%) | 0 (0%) | 1 (14%) | 0 (0%) |
| G1b. My observations allowed me to provide feedback and guidance to the parents regarding their skill utilization. | 7 (100%) | 0 (0%) | 4 (57%) | 2 (29%) | 1 (14%) | 0 (0%) |
| G2. In a child mini-playtime session, I used the Nourishing Skills worksheet to promote positive play and interactions between myself and the child. | 6 (86%) | 0 (0%) | 4 (67%) | 2 (33%) | 0 (0%) | 0 (0%) |

| | | | | | | |
|---|---|---|---|---|---|---|
| G3. I guided the family in a psychoeducational activity about the nature of emotions, gradations of emotion, and emotion regulation strategies using copies of the relevant worksheets (i.e., Understanding Feeling, Taking Care of Myself and My Feeling, and How Am I Feeling). | 7 (100%) | 0 (0%) | 5 (71%) | 0 (0%) | 1 (14%) | 1 (14%) |
| G4. I inquired how the positive behavior plan has been going at home and discussed any adjustments to the extent appropriate with the child present. | 7 (100%) | 1 (14%) | 4 (57%) | 0 (0%) | 2 (29%) | 0 (0%) |
| Session 8 | | | | | | |
| G1. I checked whether the parents consistently spent quality time with their child and how the behavioral plan progressed. If necessary, adjustments were made to the behavioral plan. | 7 (100%) | 0 (0%) | 4 (57%) | 1 (14%) | 1 (14%) | 1 (14%) |
| G2. I engaged in a discussion with the parents regarding their perspectives on the therapy's progression for both themselves and their child, evaluating both successes and challenges. | 7 (100%) | 0 (0%) | 7 (100%) | 0 (0%) | 0 (0%) | 0 (0%) |
| G3. I supported parents in thinking about their own wellbeing and any current stressors and/or needs that are affecting them referring to Recipe - Ingredient 7 worksheet about Parent Wellbeing. | 7 (100%) | 0 (0%) | 5 (71%) | 1 (14%) | 1 (14%) | 0 (0%) |
| Session 9 | | | | | | |
| G1. I checked whether the parents consistently spent quality time with their child and how the behavioral plan progressed. If necessary, adjustments were made to the behavioral plan. | 7 (100%) | 0 (0%) | 5 (71%) | 1 (14%) | 1 (14%) | 0 (0%) |
| G2a. I guided the parents through the principles of delivering clear and effective instructions to children using the Recipe - Ingredient 8 worksheet. | 6 (86%) | 0 (0%) | 5 (83%) | 0 (0%) | 1 (17%) | 0 (0%) |
| G2b. I engaged the parents in a practice activity using the Clear Directions Practice Activity cards to practice creating clear directions or selecting an appropriate alternative to giving directions. | 6 (86%) | 0 (0%) | 5 (83%) | 0 (0%) | 1 (17%) | 0 (0%) |
| G3. I shared information about the importance of providing children with safety and security using the Recipe - Ingredient 9 worksheet. | 6 (86%) | 0 (0%) | 5 (83%) | 1 (17%) | 0 (0%) | 0 (0%) |
| Session 10 | | | | | | |
| G1. I checked whether the parents consistently spent quality time with their child and how the behavioral plan progressed. If necessary, adjustments were made to the behavioral plan. | 7 (100%) | 1 (14%) | 5 (71%) | 0 (0%) | 1 (14%) | 0 (0%) |
| G2. I helped the parents understand how both parents and children can get caught in a behavioral cycle and how parents can break this pattern using the Understanding the Coercive Cycle worksheet. | 7 (100%) | 0 (0%) | 5 (71%) | 2 (29%) | 0 (0%) | 0 (0%) |
| G3a. We discussed the importance of setting limits and establishing consequences suitable for the child's age when addressing misbehavior. | 6 (86%) | 0 (0%) | 4 (67%) | 2 (33%) | 0 (0%) | 0 (0%) |
| G3b. We identified concerning behaviors and made a plan for addressing the target behavior by utilizing the Recipe - Ingredient 10 worksheet to create a consequence strategy. | 4 (57%) | 0 (0%) | 4 (100%) | 0 (0%) | 0 (0%) | 0 (0%) |
| G4. I asked the parents to focus this upcoming week on teaching the desired behaviors to their child. We discussed the approach, giving extra praise for these behaviors and designating who would lead when presenting the consequence plan to the child on the next session. | 5 (71%) | 0 (0%) | 4 (80%) | 1 (20%) | 0 (0%) | 0 (0%) |
| Session 11 | | | | | | |

| | | | | | | |
|---|---|---|---|---|---|---|
| G1. I checked whether the parents consistently spent quality time with their child and how the behavioral plan progressed, providing praise for the success or remaining neutral if the child did not meet the goals. | 7 (100%) | 0 (0%) | 6 (86%) | 0 (0%) | 1 (14%) | 0 (0%) |
| G2. I helped the parents introduce the consequence plan to the child during this session and answer the child's questions. | 1 (14%) | 0 (0%) | 1 (100%) | 0 (0%) | 0 (0%) | 0 (0%) |
| G3. I encouraged parents and the child to enjoy quality time together by playing with suitable toys during a brief playtime office session. | 7 (100%) | 0 (0%) | 7 (100%) | 0 (0%) | 0 (0%) | 0 (0%) |
| G4. I instructed parents to initiate the consequence plan and provided them with the Home Practice Goals worksheet. | 1 (14%) | 0 (0%) | 1 (100%) | 0 (0%) | 0 (0%) | 0 (0%) |
| Session 12 | | | | | | |
| G1. I checked whether the parents consistently spent quality time with their child and how the behavioral plan progressed. If necessary, adjustments were made to the behavioral plan. | 7 (100%) | 1 (14%) | 6 (86%) | 0 (0%) | 0 (0%) | 0 (0%) |
| G2a. I asked the parents to complete the Self-Assessment worksheet and briefly discussed the skills that are going well for them and areas for improvement. | 6 (86%) | 2 (33%) | 4 (67%) | 0 (0%) | 0 (0%) | 0 (0%) |
| G2b. We evaluated the implementation of the consequence plan, making adjustments as needed and planning for the coming week. | 2 (29%) | 0 (0%) | 2 (100%) | 0 (0%) | 0 (0%) | 0 (0%) |
| G3. If time allowed after adjusting the consequence plan, I prompted the parents to review the Stress and Coping worksheet at home. During the session, we briefly discussed their emotional needs. | 6 (86%) | 1 (17%) | 4 (67%) | 1 (17%) | 0 (0%) | 0 (0%) |
| Session 13 | | | | | | |
| G1. I checked whether the parents consistently spent quality time with their child and how the behavioral and consequence plans progressed. If necessary, adjustments were made to these plans. | 7 (100%) | 1 (14%) | 5 (71%) | 1 (14%) | 0 (0%) | 0 (0%) |
| G2. We briefly recapped the skills learned for improving and managing child behavior using the Recipe Card worksheet. | 7 (100%) | 1 (14%) | 6 (86%) | 0 (0%) | 0 (0%) | 0 (0%) |
| G3. I prepared the parents for the upcoming end of therapy, anticipating future challenges and ways to respond while providing the option of future additional family meetings. | 7 (100%) | 1 (14%) | 4 (57%) | 1 (14%) | 1 (14%) | 0 (0%) |
| Session 14 | | | | | | |
| G1. I checked whether the parents consistently spent quality time with their child and how the behavioral and consequence plans progressed. If necessary, adjustments were made to these plans. | 7 (100%) | 1 (14%) | 5 (71%) | 1 (14%) | 0 (0%) | 0 (0%) |
| G2. We talked about the work each family member has done during the course of therapy, eliciting their perceptions of progress made and remaining hopes and goals for the future. | 7 (100%) | 1 (14%) | 5 (71%) | 1 (14%) | 0 (0%) | 0 (0%) |
| G3. I engaged the family in an activity that encourages sharing emotions with each other by playing the “Family Feelings Hot Potato” game. | 6 (86%) | 1 (17%) | 3 (50%) | 1 (17%) | 1 (17%) | 0 (0%) |
| G4. I gave the child the certificate and goodbye note to mark their work during therapy and recognize the end of this course of therapy. | 7 (100%) | 3 (43%) | 3 (43%) | 1 (14%) | 0 (0%) | 0 (0%) |

| | | | | | | |
|---|---|---|---|---|---|---|
| G5. I asked the parents to continue with the consequence plan and provided them with Week 15 Home Practice Goals to document their progress. I encouraged them to communicate clear directions to their child and spend quality time together. | 6 (86%) | 1 (17%) | 3 (50%) | 2 (33%) | 0 (0%) | 0 (0%) |
| Session 15 | | | | | | |
| G1. I checked whether the parents consistently spent quality time with their child and how the behavioral and consequence plans progressed. If necessary, adjustments were made to these plans. | 6 (86%) | 1 (17%) | 3 (50%) | 2 (33%) | 0 (0%) | 0 (0%) |
| G2. I ensured that parents had a clear understanding of how to proceed once therapy concluded. | 7 (100%) | 1 (14%) | 5 (71%) | 1 (14%) | 0 (0%) | 0 (0%) |
| G3. I gave parents the opportunity to reflect on what they have learned and what has changed over the course of therapy. | 7 (100%) | 1 (14%) | 5 (71%) | 1 (14%) | 0 (0%) | 0 (0%) |
| G4. I gave each parent a certificate and a goodbye note to mark their work during therapy and concretely recognized the end of the course of therapy. | 7 (100%) | 1 (14%) | 5 (71%) | 1 (14%) | 0 (0%) | 0 (0%) |
| **BPT TREATMENT PROTOCOL: PILOT 2** | **Item completion n (%)** | **Very easy** | **Easy** | **Neutral** | **Difficult** | **Very difficult y** |
| Session 1 | | | | | | |
| G1a. I started to get to know the parents, with the aim of creating a better connection. | 26 (100%) | 11 (42%) | 12 (46%) | 3 (12%) | 0 (0%) | 0 (0%) |
| G2a. I explained the context of this treatment (e.g. duration, frequency) ("Structure of the treatment" and "Prescription for improving behaviour"). | 26 (100%) | 9 (35%) | 14 (54%) | 3 (12%) | 0 (0%) | 0 (0%) |
| G3a.I encouraged reflection and exchange of views with parents. | 26 (100%) | 11 (42%) | 13 (50%) | 1 (3.8%) | 1 (3.8%) | 0 (0%) |
| G4a.Discussed with parents about parenting (worksheet "Knowing ourselves - session 1"). | 26 (100%) | 5 (19%) | 18 (69%) | 3 (12%) | 0 (0%) | 0 (0%) |
| G5a. I introduced parents to the principles of reinforcing positive behaviour ("The power of praise and positive attention" worksheet). | 26 (100%) | 9 (35%) | 14 (54%) | 2 (7.7%) | 1 (3.8%) | 0 (0%) |
| G5b. I worked with the parents in formulating a plan to reinforce positive behaviour ("Prescription - 1st ingredient" and "Home practice - 1st week"). | 25 (96%) | 7 (28%) | 14 (56%) | 2 (8.0%) | 2 (8.0%) | 0 (0%) |
| G6a. I praised the parents for their efforts during the session. | 26 (100%) | 10 (38%) | 12 (46%) | 3 (12%) | 1 (3.8%) | 0 (0%) |
| Session 2 | | | | | | |
| G1a. I welcomed parents to the therapy, providing a brief summary of the structure and objectives of today's session | 25 (100%) | 15 (60%) | 8 (32%) | 2 (8.0%) | 0 (0%) | 0 (0%) |
| G2a. I reviewed last week's homework with the parents. | 25 (100%) | 7 (28%) | 15 (60%) | 3 (12%) | 0 (0%) | 0 (0%) |
| G3a. I encouraged reflection and exchange with parents with curiosity and empathy ("Knowing ourselves - 2nd session"). | 25 (100%) | 11 (44%) | 13 (52%) | 1 (4.0%) | 0 (0%) | 0 (0%) |

| | | | | | | |
|---|---|---|---|---|---|---|
| G4a. I discussed with the parents how thoughts, feelings and behaviours are connected and influence each other (Cognitive Triad worksheet). | 25 (100%) | 8 (32%) | 14 (56%) | 2 (8.0%) | 0 (0%) | 1 (4.0%) |
| G5a. I worked with parents to solidify the plan for reinforcing positive behavior ("Prescription - 2nd ingredient" and "Practice goals for home - week 2"). | 24 (96%) | 7 (29%) | 14 (58%) | 3 (13%) | 0 (0%) | 0 (0%) |
| G6a. I praised the parents for their efforts in the session, stressing the importance of gradual change. | 25 (100%) | 8 (32%) | 13 (52%) | 3 (12%) | 1 (4.0%) | 0 (0%) |
| Session 3 | | | | | | |
| G1a. I welcomed parents to the therapy, providing a brief summary of the structure and objectives of today's session | 25 (100%) | 13 (52%) | 11 (44%) | 1 (4.0%) | 0 (0%) | 0 (0%) |
| G2a. I reviewed last week's homework with the parents. | 25 (100%) | 9 (36%) | 13 (52%) | 3 (12%) | 0 (0%) | 0 (0%) |
| G3a. I discussed with parents about different parenting styles (worksheet "Parenting styles"). | 25 (100%) | 10 (40%) | 13 (52%) | 2 (8.0%) | 0 (0%) | 0 (0%) |
| G4a. I discussed with parents about their role in shaping behaviours for their children (worksheet "Recipe - 3rd ingredient"). | 25 (100%) | 6 (24%) | 13 (52%) | 4 (16%) | 2 (8.0%) | 0 (0%) |
| G5a. I worked with parents on a plan about model imitation and teaching desired behaviors at home (worksheets "Model Imitation/Training" and "Practice Objectives for Home - Week 3"). | 23 (92%) | 9 (39%) | 11 (48%) | 2 (8.7%) | 1 (4.3%) | 0 (0%) |
| Session 4 | | | | | | |
| G1a. I welcomed parents to the therapy, providing a brief summary of the structure and objectives of today's session | 25 (100%) | 15 (60%) | 9 (36%) | 1 (4.0%) | 0 (0%) | 0 (0%) |
| G2a. I discussed with parents the usual functions of behaviour and the underlying causes of their child's behaviour (worksheet "Understanding behaviour"). | 25 (100%) | 7 (28%) | 12 (48%) | 5 (20%) | 1 (4.0%) | 0 (0%) |
| G3a. Discussed with parents the benefits of positive behaviour plans towards behaviour change (worksheet "Prescription - 4th component"). | 25 (100%) | 5 (20%) | 13 (52%) | 6 (24%) | 1 (4.0%) | 0 (0%) |
| G4a. I worked with the parents to create a written positive behavior modeling plan to practice at home, focusing on a specific behavior that was identified as needing to change, during this session. | 24 (96%) | 6 (25%) | 8 (33%) | 8 (33%) | 2 (8.3%) | 0 (0%) |
| Session 5 | | | | | | |
| G1a. I welcomed the child to therapy, gave an age-appropriate explanation of my role and our work with the family. | 23 (100%) | 11 (48%) | 9 (39%) | 2 (8.7%) | 1 (4.3%) | 0 (0%) |
| G2a. I gave the parents and the child the opportunity to play freely together, with the aim of gathering information about the parent-child relationship (worksheet "Observation of parent-child play"). | 23 (100%) | 8 (35%) | 13 (57%) | 1 (4.3%) | 1 (4.3%) | 0 (0%) |
| G2b. I provided friendly and genuinely positive feedback on the family game. | 23 (100%) | 8 (35%) | 15 (65%) | 0 (0%) | 0 (0%) | 0 (0%) |
| G3a. I reinforced the child's efforts to achieve the goals during the implementation of the positive behaviour plan, | 20 (87%) | 4 (20%) | 13 (65%) | 3 (15%) | 0 (0%) | 0 (0%) |
| G3b. I worked with the family to address any barriers to implementing the practice at home. | 22 (96%) | 3 (14%) | 11 (50%) | 8 (36%) | 0 (0%) | 0 (0%) |
| Session 6 | | | | | | |

| | | | | | | |
|---|---|---|---|---|---|---|
| G1a. I checked the evolution of the behavior plan. | 24 (100%) | 6 (25%) | 12 (50%) | 3 (13%) | 3 (13%) | 0 (0%) |
| G2a. I explained to parents the importance of regular, enjoyable quality time with their child (worksheet "Recipe - 5th ingredient"). | 24 (100%) | 7 (29%) | 16 (67%) | 1 (4.2%) | 0 (0%) | 0 (0%) |
| G2b. I prepared parents to regularly spend quality time with their child at home ("Creating relationship time" worksheet). | 24 (100%) | 7 (29%) | 15 (63%) | 2 (8.3%) | 0 (0%) | 0 (0%) |
| G3a. I discussed emotional regulation skills with parents (worksheet "Recipe - 6th ingredient"). | 24 (100%) | 8 (33%) | 11 (46%) | 4 (17%) | 0 (0%) | 1 (4.2%) |
| G4a. I encouraged the parents to continue to implement the positive behaviour plan (worksheet "Home practice goals - week 6"). | 23 (96%) | 7 (30%) | 13 (57%) | 2 (8.7%) | 1 (4.3%) | 0 (0%) |
| Session 7 | | | | | | |
| G1a. I observed how parents were applying positive interaction skills in the play session ("Caring Skills List" and "Play Notes" worksheets). | 23 (100%) | 9 (39%) | 13 (57%) | 1 (4.3%) | 0 (0%) | 0 (0%) |
| G1b. My observations allowed me to provide feedback and guidance to parents on the use of their skills. | 22 (96%) | 6 (27%) | 12 (55%) | 4 (18%) | 0 (0%) | 0 (0%) |
| G2a. In the short play session, I promoted positive play by enhancing the therapeutic relationship between me and the child (worksheet "Notes on caring skills") | 23 (100%) | 7 (30%) | 13 (57%) | 2 (8.7%) | 1 (4.3%) | 0 (0%) |
| G3a. I discussed with the family the strategies for regulating emotions ("Understanding emotions", "Caring for myself & my emotions" and "How I feel"). | 22 (96%) | 6 (27%) | 10 (45%) | 4 (18%) | 2 (9.1%) | 0 (0%) |
| G4a. I asked how the plan for shaping positive behaviour at home is progressing. | 22 (96%) | 9 (41%) | 8 (36%) | 4 (18%) | 1 (4.5%) | 0 (0%) |
| Session 8 | | | | | | |
| G1a. I checked whether the parents were consistently spending quality time with their child and the development of the behaviour plan. | 23 (100%) | 10 (43%) | 9 (39%) | 2 (8.7%) | 2 (8.7%) | 0 (0%) |
| G2a. I talked to the parents about their views on the progress of the treatment for both themselves and their child. | 22 (96%) | 8 (36%) | 11 (50%) | 2 (9.1%) | 1 (4.5%) | 0 (0%) |
| G3a. I helped parents to think around current stressful situations and/or needs that affect them (worksheet "Recipe - 7th ingredient" for parental well-being). | 23 (100%) | 6 (26%) | 14 (61%) | 2 (8.7%) | 1 (4.3%) | 0 (0%) |
| Session 9 | | | | | | |
| G1a. I checked whether the parents regularly spent quality time with their child. | 23 (100%) | 9 (39%) | 10 (43%) | 4 (17%) | 0 (0%) | 0 (0%) |
| G2a. I briefed parents on the key principles involved in providing clear and effective instructions to children, using the "Recipe - 8th ingredient" worksheet. | 23 (100%) | 6 (26%) | 13 (57%) | 3 (13%) | 0 (0%) | 1 (4.3%) |
| G2b. I participated with parents in an activity to practice formulating clear instructions ("Activity cards with clear instructions" ). | 22 (96%) | 5 (23%) | 12 (55%) | 3 (14%) | 2 (9.1%) | 0 (0%) |
| G3a. I shared information about the importance of providing safety to children using the "Recipe - 9th ingredient" worksheet. | 22 (96%) | 4 (18%) | 13 (59%) | 4 (18%) | 1 (4.5%) | 0 (0%) |
| Session 10 | | | | | | |

| | | | | | | |
|---|---|---|---|---|---|---|
| G1a. I checked whether parents were consistently spending quality time with their child and the development of the behaviour plan. | 23 (100%) | 12 (52%) | 7 (30%) | 4 (17%) | 0 (0%) | 0 (0%) |
| G2a. I helped parents understand how they can break the pattern of the cycle of behaviour (worksheet "Understanding the cycle of coercion"). | 22 (96%) | 8 (36%) | 9 (41%) | 5 (23%) | 0 (0%) | 0 (0%) |
| G3a. We discussed the importance of setting limits and consequences appropriate to the child's age. | 22 (96%) | 6 (27%) | 10 (45%) | 6 (27%) | 0 (0%) | 0 (0%) |
| G3b. We made a plan to address a specific unwanted behaviour (worksheet "Prescription - 10th ingredient"). | 17 (74%) | 6 (35%) | 8 (47%) | 2 (12%) | 1 (5.9%) | 0 (0%) |
| G4a. I asked the parents to focus in the coming week on teaching their child desirable behaviors. | 22 (96%) | 7 (32%) | 14 (64%) | 1 (4.5%) | 0 (0%) | 0 (0%) |
| Session 11 | | | | | | |
| G1a. I checked whether parents were consistently spending quality time with their child and the development of the behaviour plan, | 23 (100%) | 9 (39%) | 12 (52%) | 2 (8.7%) | 0 (0%) | 0 (0%) |
| G2a. I helped the parents present the consequence plan to the child during this session. | 15 (65%) | 5 (33%) | 6 (40%) | 3 (20%) | 1 (6.7%) | 0 (0%) |
| G3a. I encouraged parents and child to spend quality time in the short play session. | 21 (91%) | 7 (33%) | 10 (48%) | 1 (4.8%) | 3 (14%) | 0 (0%) |
| G4a. I instructed the parents to start the consequence plan and gave them the worksheet for the practice objectives for the homework. | 14 (61%) | 5 (36%) | 6 (43%) | 2 (14%) | 1 (7.1%) | 0 (0%) |
| Session 12 | | | | | | |
| G1a. I checked whether parents were consistently spending quality time with their child and the development of the behaviour plan. | 22 (96%) | 4 (18%) | 16 (73%) | 2 (9.1%) | 0 (0%) | 0 (0%) |
| G2a. I asked the parents to fill in the self-evaluation sheet | 19 (83%) | 3 (16%) | 10 (53%) | 5 (26%) | 1 (5.3%) | 0 (0%) |
| G2b. We evaluated the implementation of the impact plan and made the necessary adjustments. | 15 (65%) | 2 (13%) | 6 (40%) | 5 (33%) | 2 (13%) | 0 (0%) |
| G3a. I encouraged parents to review the "Stress and its management" worksheet at home. | 23 (100%) | 4 (17%) | 14 (61%) | 5 (22%) | 0 (0%) | 0 (0%) |
| Session 13 | | | | | | |
| G1a. I checked whether parents were consistently spending quality time with their child and the development of behavioral and consequence plans. | 22 (96%) | 7 (32%) | 12 (55%) | 2 (9.1%) | 1 (4.5%) | 0 (0%) |
| G2a. We briefly summarised the skills that parents were taught to manage their child's behaviour (recipe card worksheet). | 23 (100%) | 7 (30%) | 15 (65%) | 1 (4.3%) | 0 (0%) | 0 (0%) |
| G3a. I prepared the parents for the end of treatment, anticipating future difficulties and ways of coping. | 23 (100%) | 3 (13%) | 15 (65%) | 4 (17%) | 1 (4.3%) | 0 (0%) |
| Session 14 | | | | | | |
| G1a. I checked whether parents were consistently spending quality time with their child and the development of behavioral and consequence plans. | 23 (100%) | 10 (43%) | 11 (48%) | 2 (8.7%) | 0 (0%) | 0 (0%) |
| G2a. We talked about the work that each family member has done during treatment. | 23 (100%) | 10 (43%) | 9 (39%) | 3 (13%) | 1 (4.3%) | 0 (0%) |

| | | | | | | |
|---|---|---|---|---|---|---|
| G3a. I engaged the family in an activity that encourages the expression of feelings between them by playing the game "Family Feelings - Hot Potato". | 23 (100%) | 6 (26%) | 12 (52%) | 3 (13%) | 2 (8.7%) | 0 (0%) |
| G4a. I gave the child his graduation certificate and farewell note. | 23 (100%) | 11 (48%) | 10 (43%) | 1 (4.3%) | 1 (4.3%) | 0 (0%) |
| G5a. I encouraged parents to give clear instructions to their child and to spend quality time together. | 23 (100%) | 7 (30%) | 15 (65%) | 1 (4.3%) | 0 (0%) | 0 (0%) |
| Session 15 | | | | | | |
| G1a. I checked whether parents were consistently spending quality time with their child and the development of behavioral and consequence plans. | 23 (100%) | 12 (52%) | 11 (48%) | 0 (0%) | 0 (0%) | 0 (0%) |
| G2a. I made sure that the parents clearly understood how to proceed after the treatment was completed. | 23 (100%) | 9 (39%) | 12 (52%) | 2 (8.7%) | 0 (0%) | 0 (0%) |
| G3a. I gave parents the opportunity to reflect on what they learned and what changed during the treatment. | 23 (100%) | 9 (39%) | 13 (57%) | 1 (4.3%) | 0 (0%) | 0 (0%) |
| G4a. I gave each parent a certificate and a goodbye note. | 22 (96%) | 11 (50%) | 10 (45%) | 0 (0%) | 1 (4.5%) | 0 (0%) |

### Supplementary Figure 1 - National survey with professionals: clinical challenges

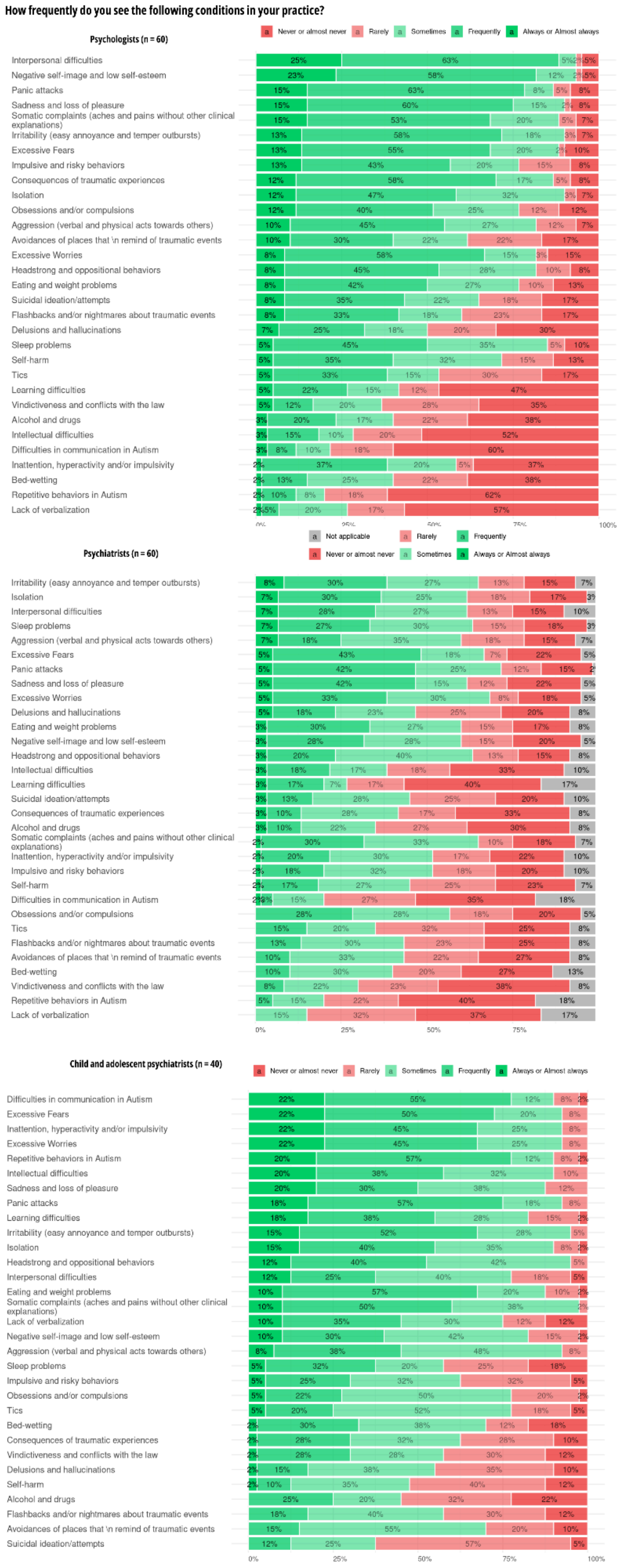

### Supplementary Figure 2 - National survey with professionals: background training

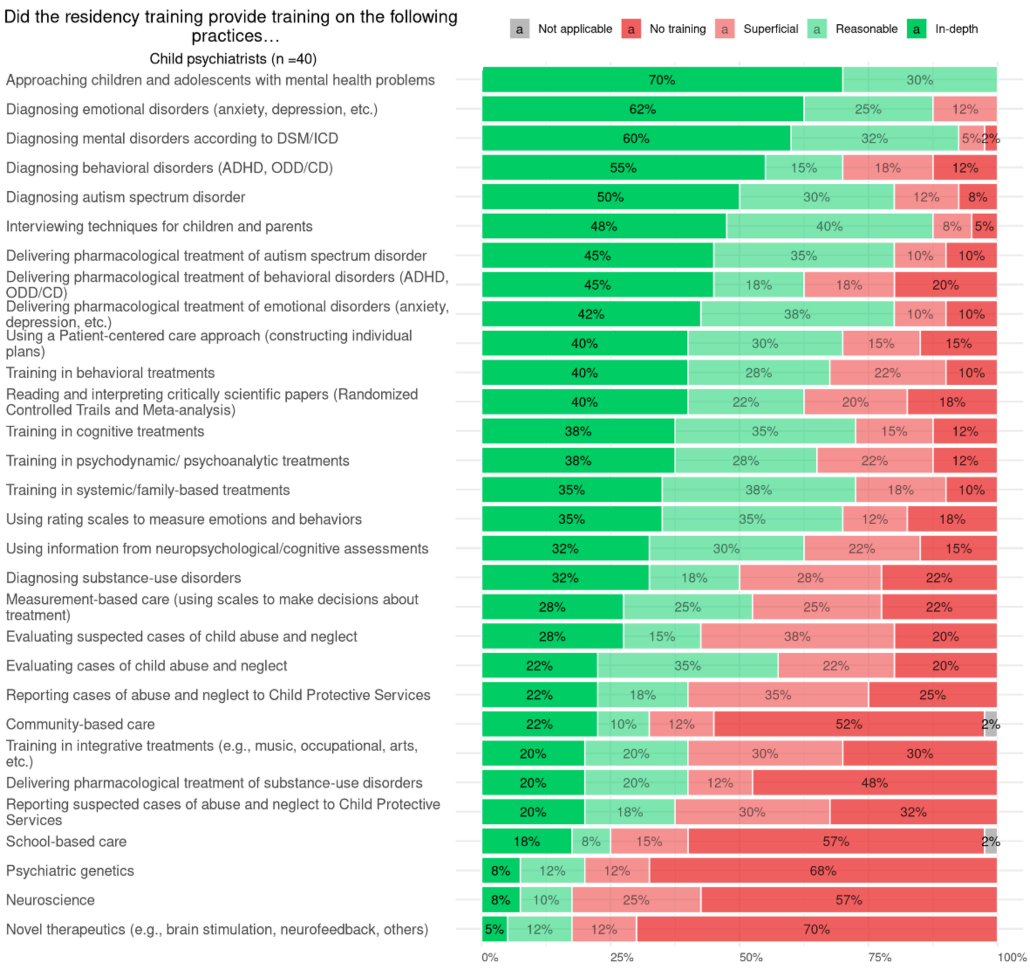


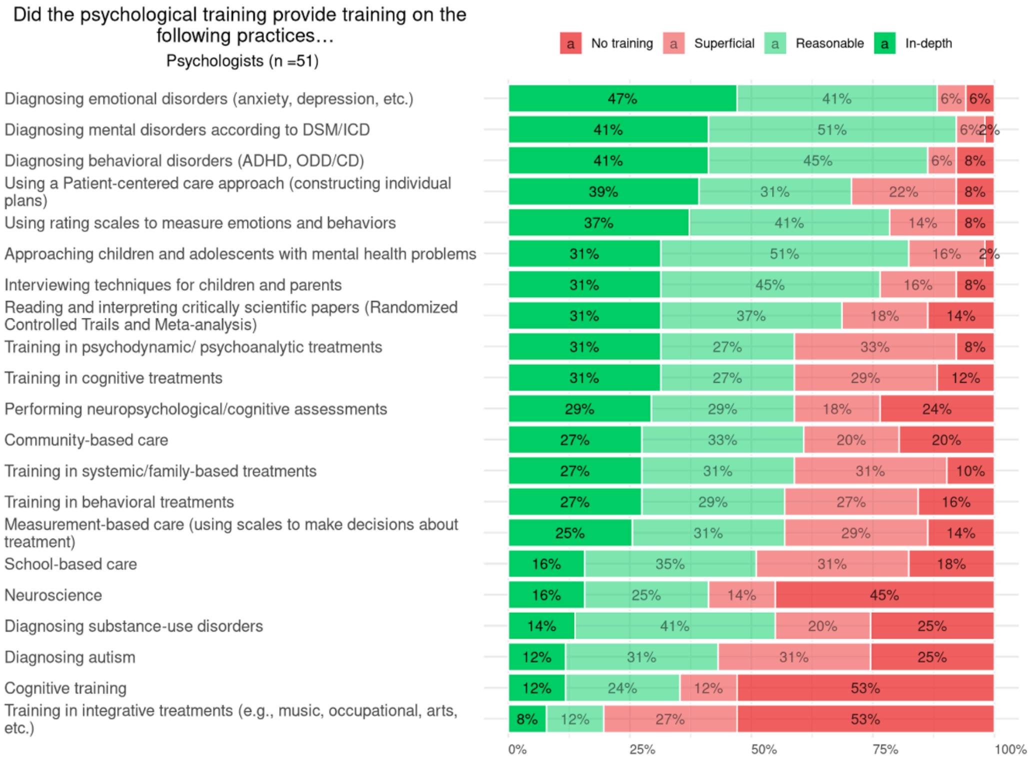

### Supplementary Figure 3 - National survey assessing needs with professionals: training interests

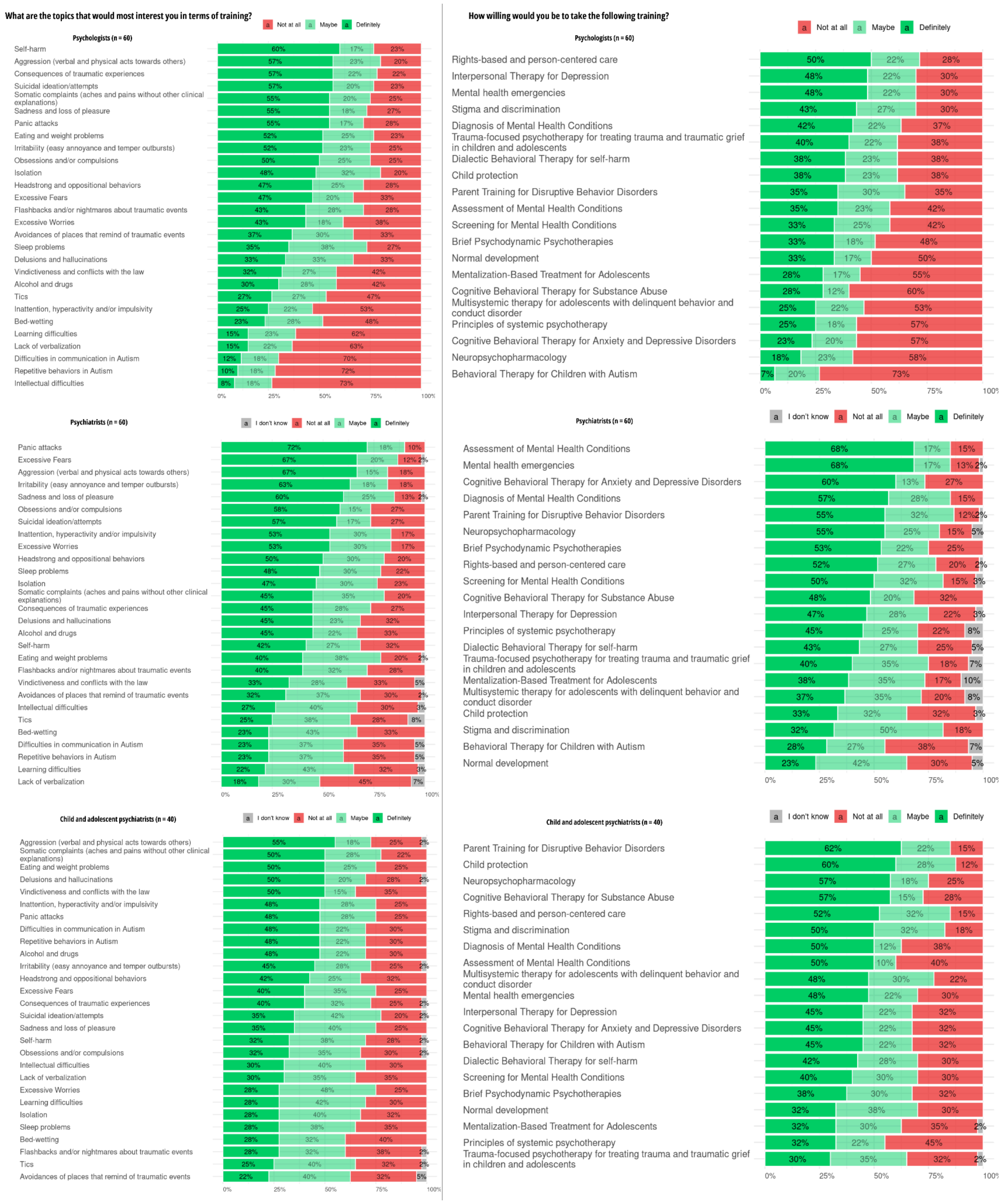

Supplementary Figure 4. Clinical scores across protocol sessions

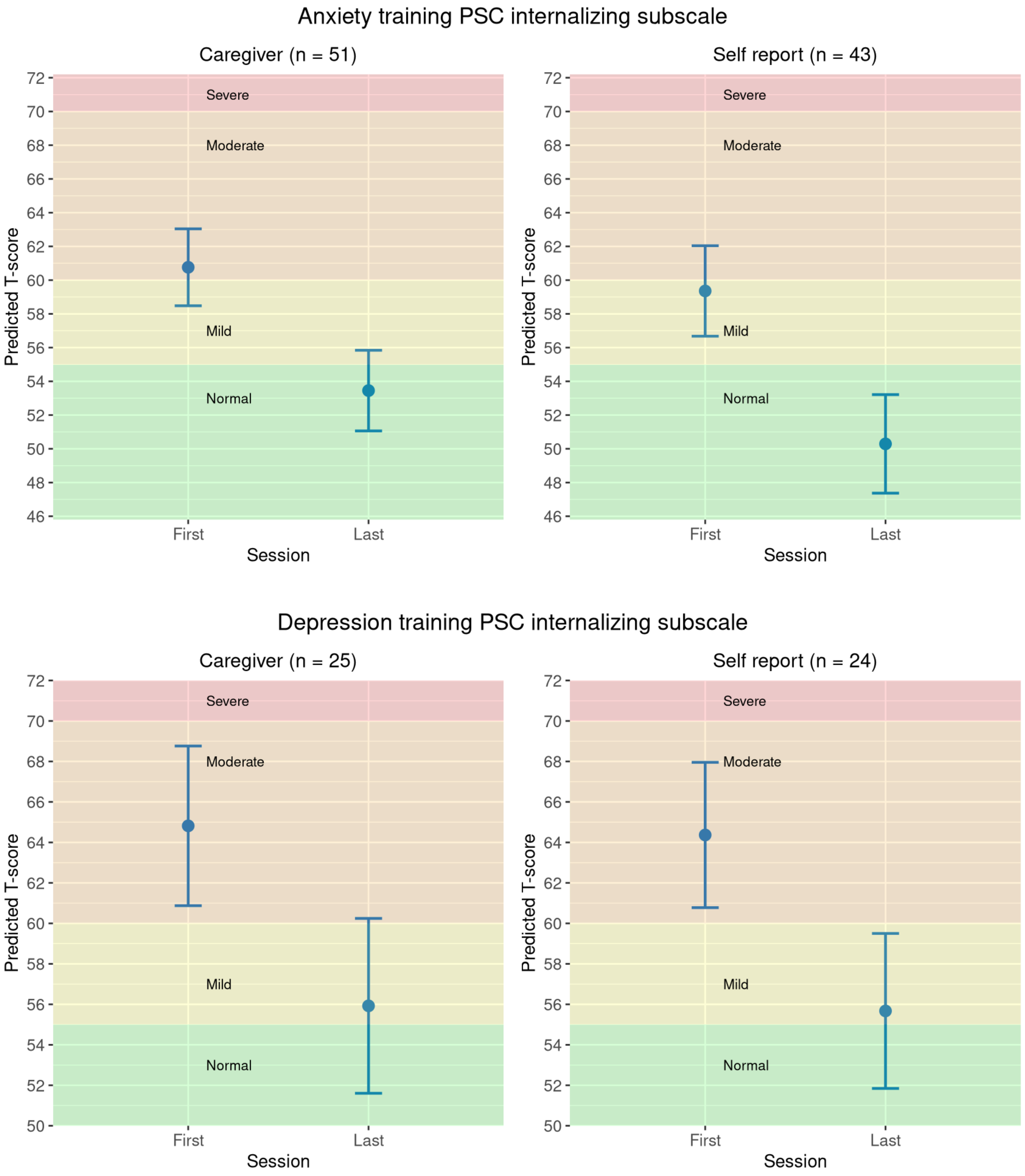

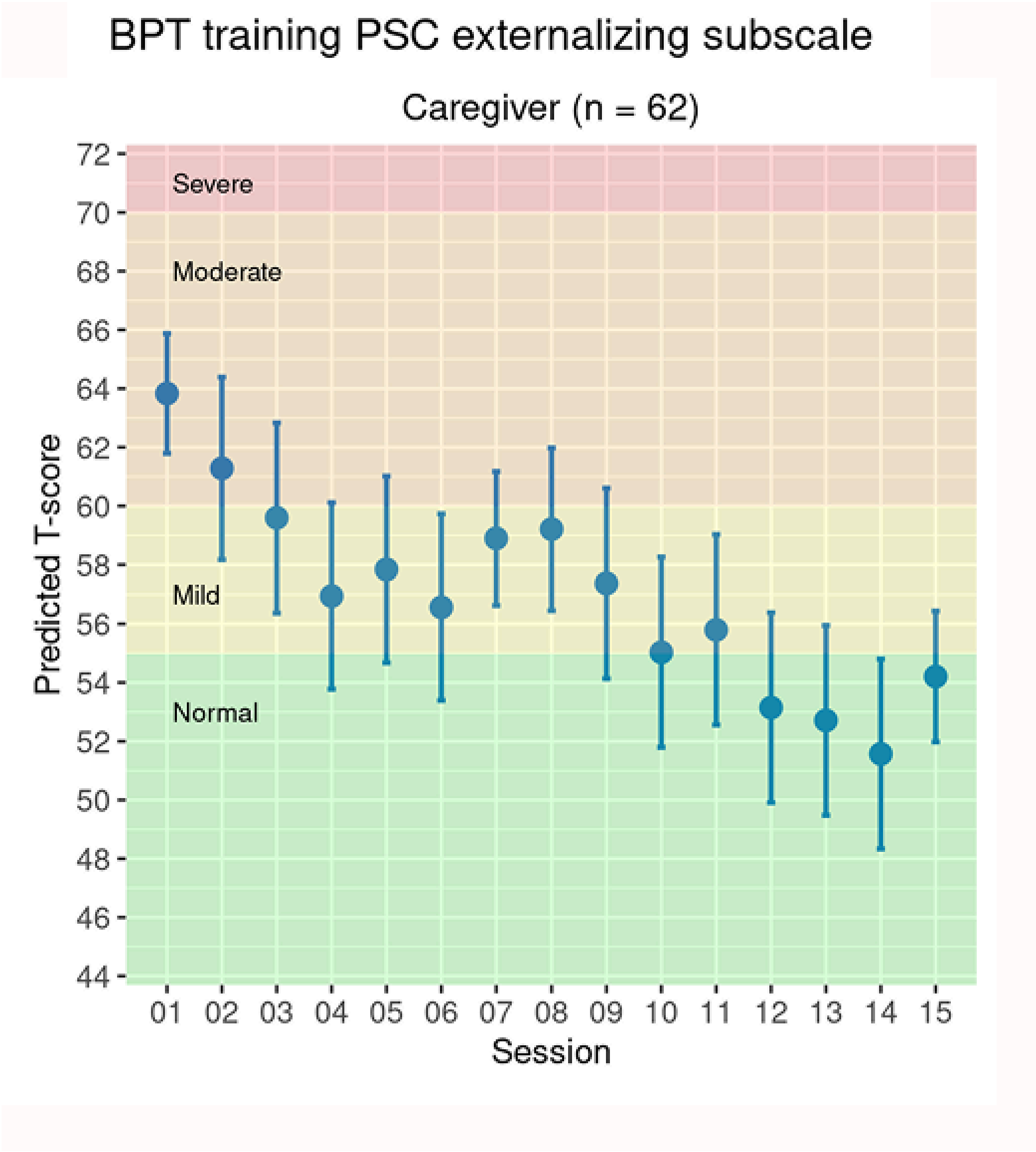

BPT training PSC externalizing subscale
Caregiver (n = 62)
Severe
Moderate
Mild
Normal
Predicted T-score
72
70
68
66
64
62
60
58
56
54
52
50
48
46
44
01 02 03 04 05 06 07 08 09 10 11 12 13 14 15
Session

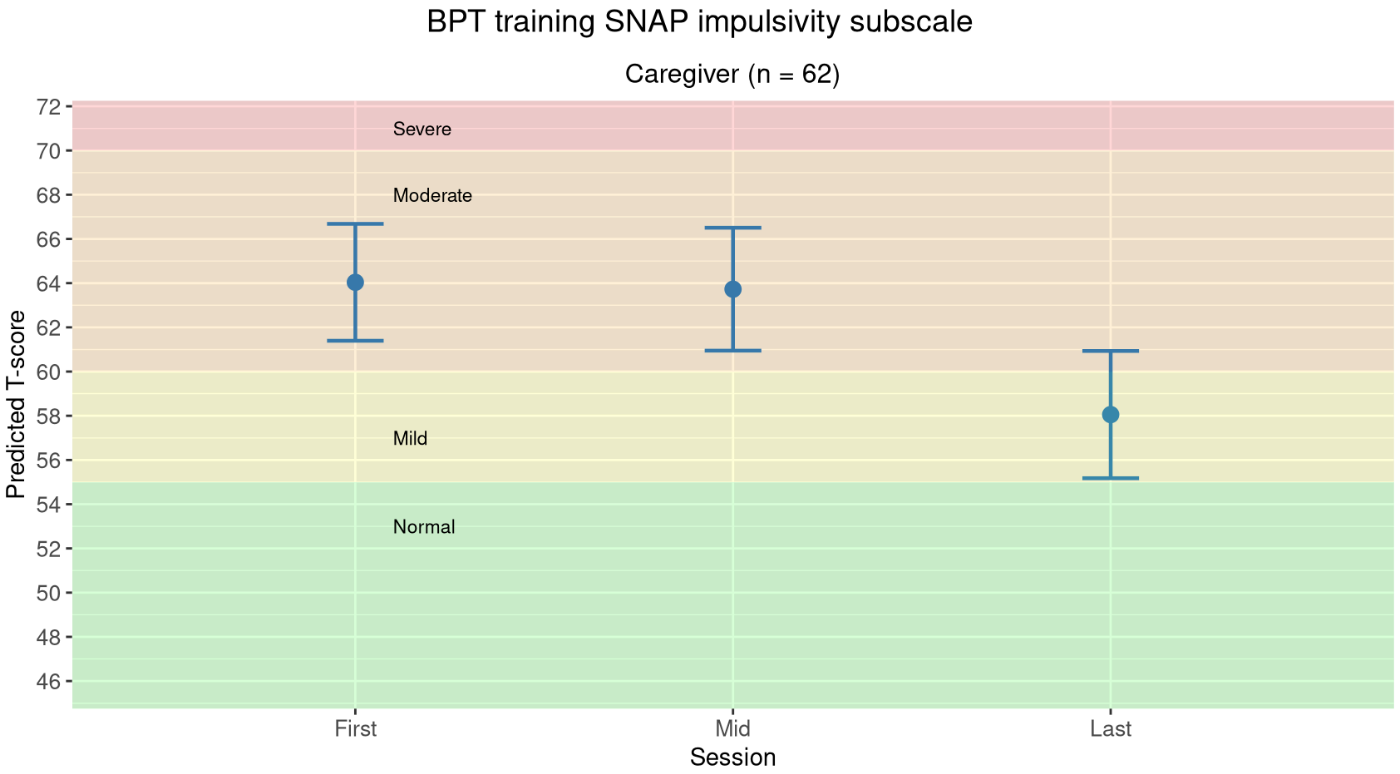
BPT training SNAP impulsivity subscale
Caregiver (n = 62)
Predicted T-score
72
70
68
66
64
62
60
58
56
54
52
50
48
46
Severe
Moderate
Mild
Normal
First
Mid
Last
Session

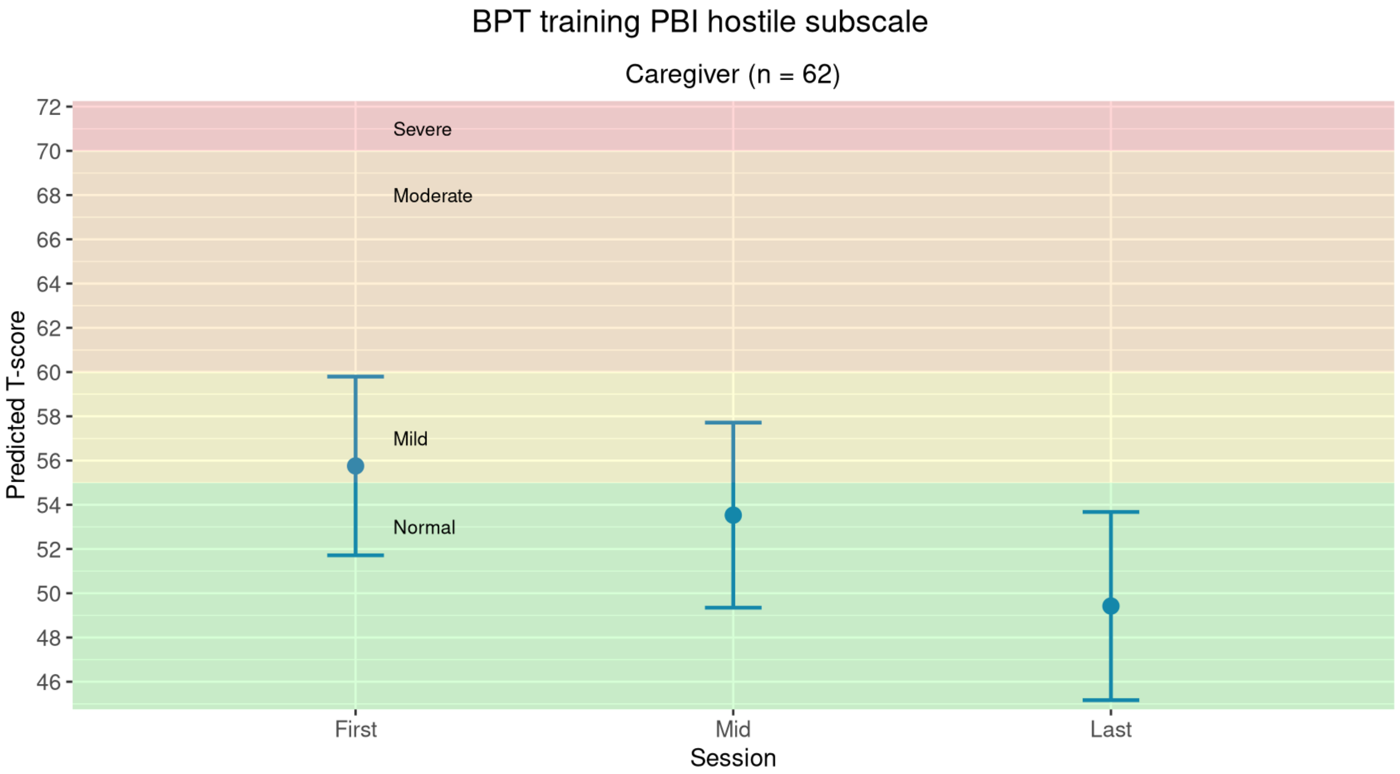
BPT training PBI hostile subscale
Caregiver (n = 62)
Predicted T-score
72
70
68
66
64
62
60
58
56
54
52
50
48
46
Severe
Moderate
Mild
Normal
First
Mid
Last
Session

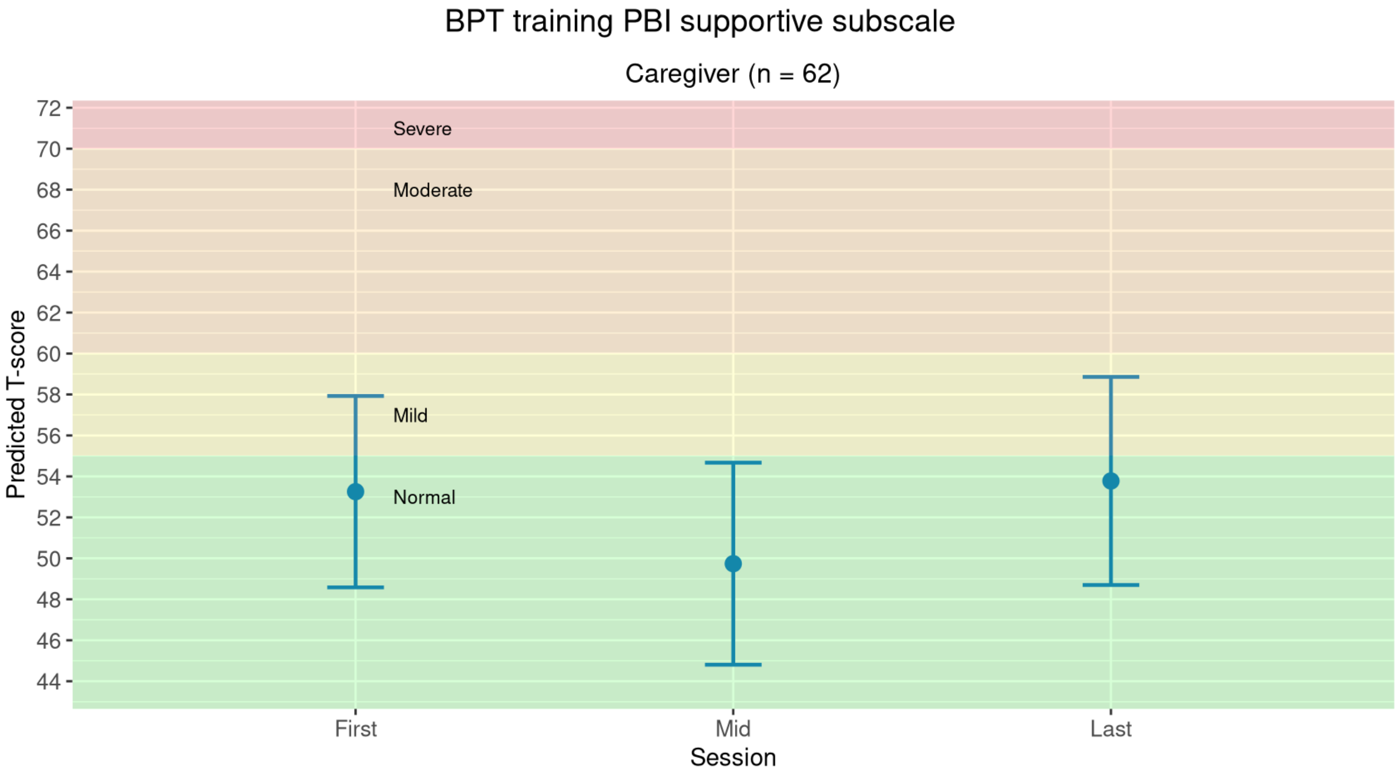
BPT training PBI supportive subscale
Caregiver (n = 62)
Predicted T-score
72
70
68
66
64
62
60
58
56
54
52
50
48
46
44
Severe
Moderate
Mild
Normal
First
Mid
Last
Session

Supplementary Figure 5. Estimated percentage change on clinical scores across protocol sessions

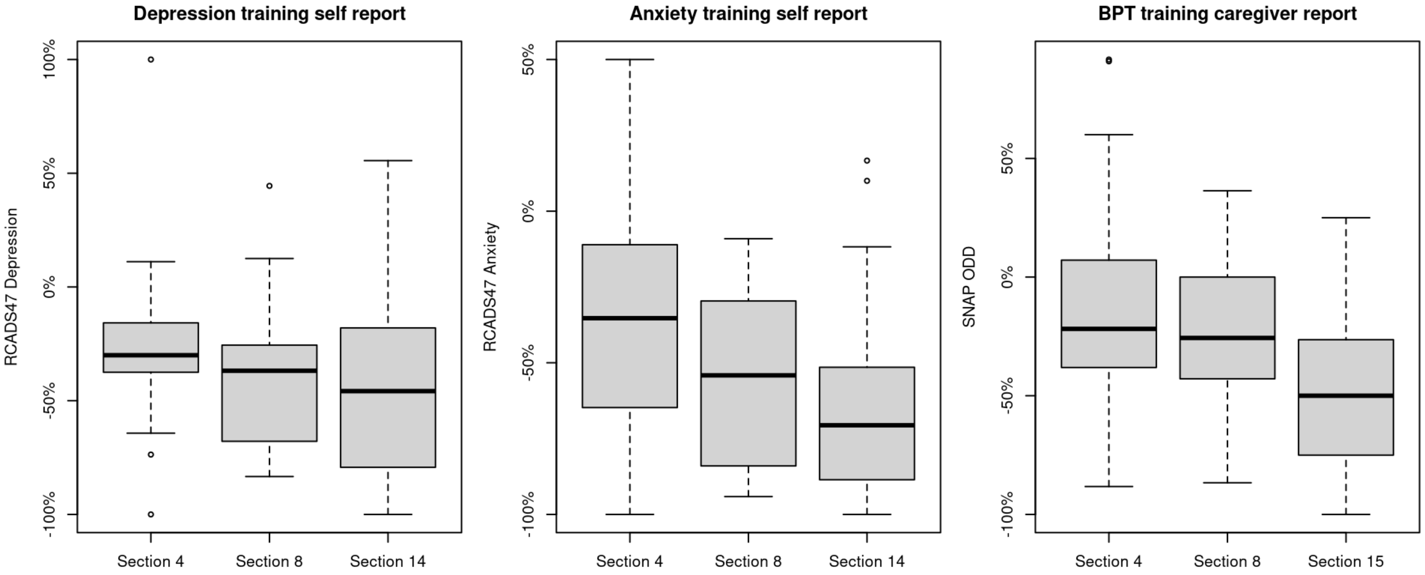


**Notes**: Box spans the interquartile range (25th to 75th percentile) with median line at the 50th percentile. Whiskers represent the range of observed values excluding outliers (shown as individual points).

### Supplementary Figure 6 - Pilot implementation: satisfaction rates

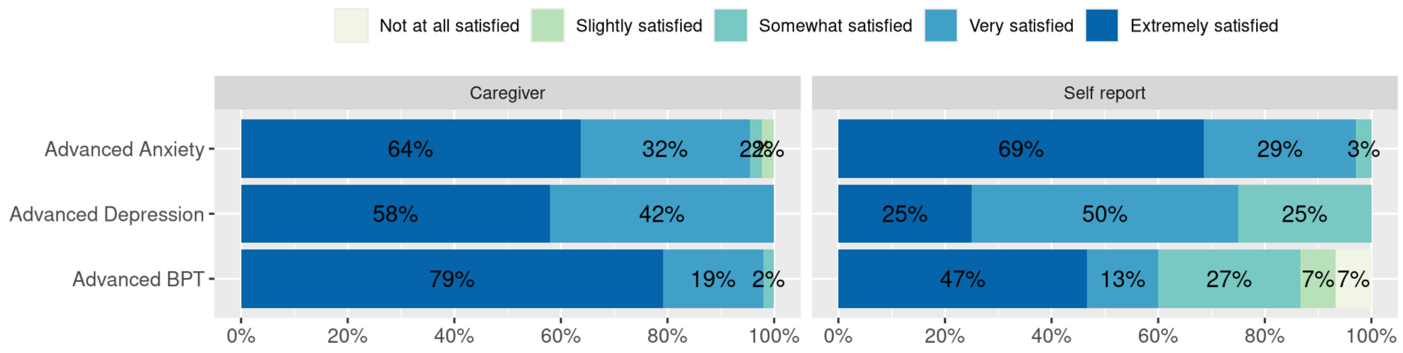

Supplementary Figure 7 - Completion rate of manual items

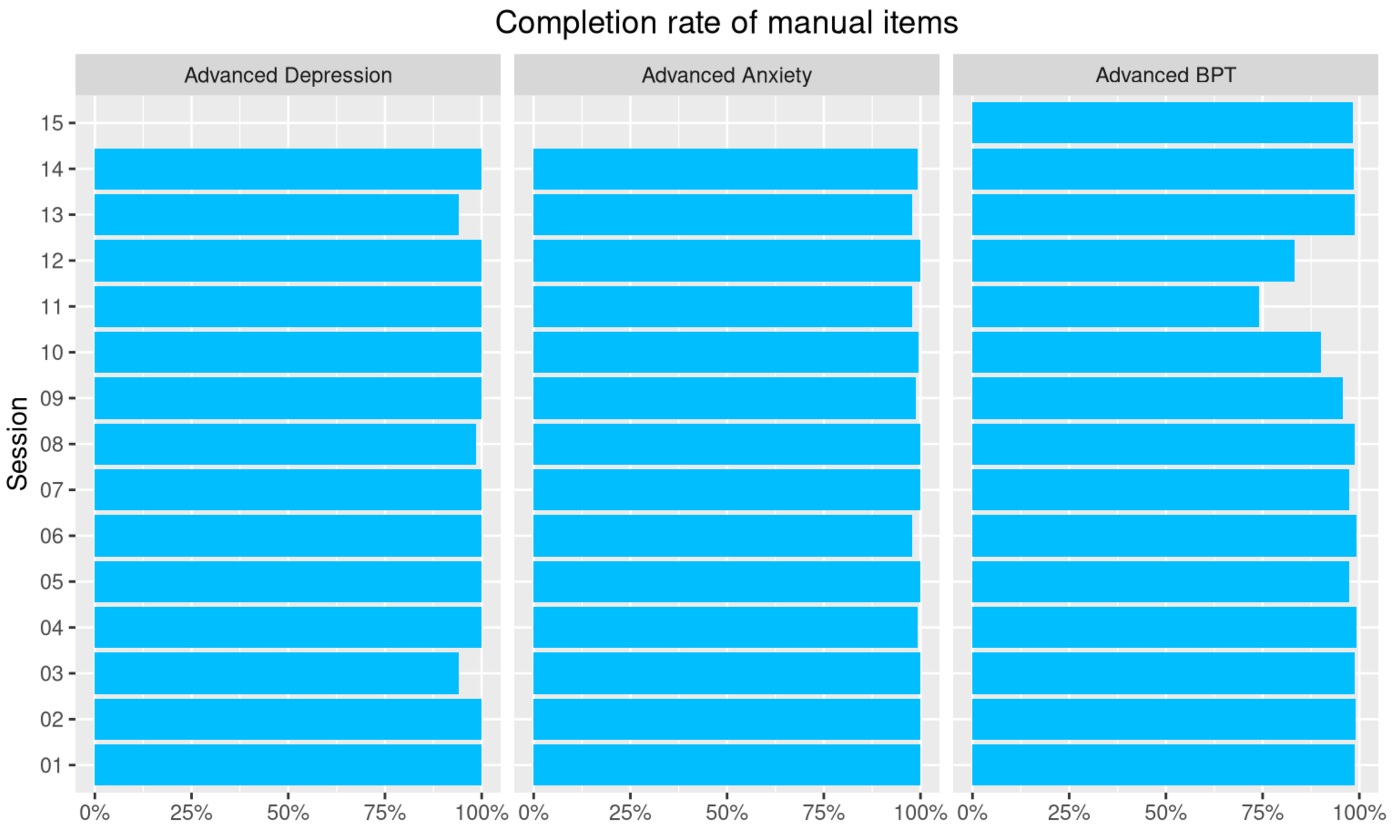
